# Distinct longitudinal brain white matter microstructure changes and associated polygenic risk of common psychiatric disorders and Alzheimer’s disease in the UK Biobank

**DOI:** 10.1101/2023.10.19.23297257

**Authors:** Max Korbmacher, Dennis van der Meer, Dani Beck, Daniel E. Askeland-Gjerde, Eli Eikefjord, Arvid Lundervold, Ole A. Andreassen, Lars T. Westlye, Ivan I. Maximov

**Affiliations:** Department of Health and Functioning, Western Norway University of Applied Sciences, Bergen, Norway; NORMENT Centre for Psychosis Research, Division of Mental Health and Addiction, University of Oslo and Oslo University Hospital, Oslo, Norway; Mohn Medical Imaging and Visualization Centre (MMIV), Bergen, Norway; Faculty of Health, Medicine and Life Sciences, Maastricht University, Maastricht, Netherlands; Department of Psychiatric Research, Diakonhjemmet Hospital, Oslo, Norway; Department of Psychology, University of Oslo, Oslo, Norway; Department of Radiology, Haukeland University Hospital, Bergen, Norway; Department of Biomedicine, University of Bergen, Bergen, Norway; KG Jebsen Centre for Neurodevelopmental Disorders, University of Oslo, Oslo, Norway

**Keywords:** Ageing, White Matter, Microstructure, Brain Ageing, Polygenic Risk, Mag-netic Resonance Imaging, Diffusion MRI

## Abstract

During the course of adulthood and ageing, white matter (WM) structure and organisation are characterised by slow degradation processes such as demyelination and shrinkage. An acceleration of such ageing process has been linked to the development of a range of diseases. Thus, an accurate description of healthy brain maturation, in particular, in terms of WM features, provides a cornerstone in the understanding of ageing. We use longitudinal diffusion magnetic resonance imaging to provide an overview of WM changes at different spatial and temporal scales in the UK Biobank (UKB) (N=2,678; age_*scan*1_=62.38±7.23 years; age_*scan*2_=64.81±7.1 years). To examine the genetic overlap between WM structure and common clinical conditions, we tested the associations between WM structure and polygenic risk scores (PGRS) for the most common neurodegenerative disorder, Alzheimer’s disease, and common psychiatric disorders (uniand bipolar depression, anxiety, obsessive-compulsive, autism, schizophrenia, attention-deficit-hyperactivity) in longitudinal (N=2,329) and crosssectional UKB validation data (N=31,056). Global and regional single and multi-compartment fractional anisotropy, intra-axonal water fraction, and kurtosis metrics decreased 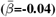, whereas diffusivity metrics, and free water increased with age 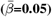, with the annual rate of WM change (ARoC) accelerating at higher ages for both global 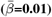 and regional WM metrics 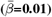. Voxel-level trends indicated decreasing anisotropy, and variable spatial patterns for other diffusion metrics, suggesting differential changes in frontal compared to other brain regions. Although effect sizes were small 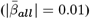, ARoC in middle cerebral peduncle WM had the strongest association with PGRS, especially for Alzheimer’s: 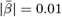. PGRS were more strongly related to ARoC than cross-sectional measures (*d*_*scan*1_=0.03, *d*_*scan*2_=0.03, *d*_*validation*_=0.03). Our findings indicate spatially distributed WM changes across the brain, as well as distributed associations of PGRS with WM. Importantly, brain longitudinal changes reflected the genetic risk for disorder development better than the utilised cross-sectional measures, with regional differences giving more specific insights into gene-brain change associations than global averages.

## Introduction

White matter microstructure (WMM) changes significantly throughout the lifespan. Recent large-scale studies suggest a strong association between WMM and age during both healthy and diseased ageing (1–7). Previous findings demonstrated general trends of tissue anisotropy increases and water diffusivity decreases throughout childhood (8–10), and, as reversal dynamics of these trends, throughout adulthood (3, 11). Importantly, abnormal white matter (WM) development has been associated with the development of neurocognitive skills and mental health symptoms in childhood and adolescence (12) and brain disorders later in life (13, 14). A person’s genetically determined propensity to develop a certain disorder can also be summarised by polygenic risk scores (PGRS) (15–23). Combining brain imaging and genetics opens the opportunity to associate the elevated genetic risk for disorders with specific brain features (24) in terms of scalar imaging metrics (25, 26). This allows to identify which brain regions might be more prone to disease developments based on the genetic makeup, and provide additional biological detail to the observed WM changes. An advantage of using WMM for such associations is the level of detail provided by biophysical models, such as intra- and extra axonal diffusion processes (27–31). WM-specific changes are associated with various common psychiatric disorders (13, 14), and precede, for example, the symptom onset in Alzheimer’s Disease (AD) (32). This outlines WMM as an important aspect for further investigation in disease formation and outcomes.

As the temporal aspect matters when investigating tissue changes, longitudinal designs are required (6). Hence, longitudinal changes in WM are informative when examining PGRS-tissue associations as they allow to connect PGRS with actual WM change and not only cross-sectional “snapshots” of brain structure at a given moment of an individual’s life.

In order to estimate ageing effects on WM alterations, studies commonly focus on diffusion tensor imaging (DTI) (33). While DTI is useful for mesostructural characterisations, DTI is also limited in addressing certain common phenomena in WM such as crossing fibre bundles, non-Gaussian diffusion, and differences between intra- and extra-axonal water compartments (34). Recent achievements in advanced diffusion magnetic resonance imaging (dMRI) techniques offer a spectrum of biophysical models (27–31) addressing these issues, for example, by differentiating between intra and extra-axonal space (27, 29, 31), or by capturing non-Gaussian diffusion (27, 28). In turn, there are few longitudinal studies observing WMM changes (10, 35, 36), and even fewer also utilising advanced diffusion approaches beyond DTI (6, 10). In order to fill this gap, we assess the metrics of a series of dMRI approaches in a large longitudinal mid-to-late life adult sample provided by the UKB (37) and identify the spatio-temporal patterns of ageing-related WMM changes on a global, regional, and voxel-level scale. To further investigate potential genetic underpinnings of these WMM changes, we estimate PGRS informed by previous genome-wide association studies for AD, the most common neurodegenerative disorder, and common psychiatric disorders, including Major Depressive Disorder (MDD), Bipolar Disorder (BIP), Anxiety Disorder (ANX), Autism Spectrum Disorder (ASD), Schizophrenia (SCZ), Attention Deficit Hyperactivity Disorder (ADHD), and Obsessive-Compulsive Disorder (OCD). PGRS capture an individual’s genetic propensity for a trait by aggregating the estimated effects of risk variants across the entire genome. Together with brain structure and brain structural changes, PGRS may be informative for the development of disease. For example, these gene-brain associations can help identify concrete spatial patterns for different diseases (38, 39). For further inference on the generalisability of the associations between PGRS and WM in the longitudinal sample with available PGRS (after exclusions: N = 2,329), we estimated the same associations for independent participants from the cross-sectional portion of the UKB (after exclusions: N = 31,056) (37). Based on previous findings (3, 40), we expected near-linear age associations of diffusion metrics, generally outlining lower fractional anisotropy, intra-axonal water fraction, and kurtosis at higher ages, but higher diffusivity, extra-axonal free water fraction. Moreover, we expected to observe PGRS-WMM associations for AD globally and in most age-sensitive regions.

## Methods

### A. Sample characteristics

We obtained UKB data (37), including the longitudinal dMRI data of N = 4,871 participants at two time points. Participant data were excluded when consent had been withdrawn, and when dMRI data were not meeting quality control (QC) standards using the YTTRIUM method (41) (see also Supplement 1). Additionally, we excluded participants which were diagnosed with any mental and behavioural disorder (ICD-10 category F), disease of the nervous system (ICD-10 category G), and disease of the circulatory system (ICD-10 category I). Remaining data sets after exclusions were applied were N = 2,678 participants (52.99% females). At baseline, participants were on average 62.26±7.19 years old (range: 46.12-80.30 years) and at time point two, mean age was 64.70±7.07 years (range: 49.33-82.59 years), indicating an average age difference as Δ*age* =2.44±0.73 years (range: 1.12-6.90 years). The data were collected at three sites: (1) in Cheadle (57.36%), (2) Newcastle (37.04%), and (3) Reading (5.60%). PGRS data were available for N = 2,329 of these longitudinal data sets and for N = 31,056 cross-sectional validation data sets (after exclusions).

### B. MRI acquisition and post-processing

UKB MRI data acquisition procedures are described elsewhere (37, 42, 43).

After obtaining access to the raw dMRI data, we preprocessed it using an optimised pipeline (41). The pipeline includes corrections for noise (44), Gibbs ringing (45), susceptibility-induced and motion distortions, and eddy currents artefacts (46). Isotropic 1 mm^3^ Gaussian smoothing was carried out using FSL’s (47, 48) *fslmaths*. Employing the multi-shell data, Diffusion Tensor Imaging (DTI), Diffusion Kurtosis Imaging (DKI) (28) and White Matter Tract Integrity (WMTI) (27) metrics were estimated using Matlab 2017b code (https://github.com/NYU-DiffusionMRI/DESIGNER). Spherical mean technique SMT (30), and multi-compartment spherical mean technique (mcSMT) (29) metrics were estimated using original code (https://github.com/ekaden/smt) (29, 30). Estimates from the Bayesian Rotational Invariant Approach (BRIA) were evaluated by the original Matlab code (https://bitbucket.org/reisert/baydiff/src/master/) (31).

In total, we obtained 26 WM metrics from six diffusion approaches (DTI, DKI, WMTI, SMT, mcSMT, BRIA; see for overview in Supplement 2). In order to normalise all metrics, we used Tract-based Spatial Statistics (TBSS) (49), as part of FSL (47, 48). In brief, initially all brain-extracted (50) fractional anisotropy (FA) images were aligned to MNI space using non-linear transformation (FNIRT) (48). Following, the mean FA image and related mean FA skeleton were derived. Each diffusion scalar map was projected onto the mean FA skeleton using TBSS. To provide a quantitative description of diffusion metrics at a region level, we used the John Hopkins University (JHU) atlas (51), and obtained 30 hemisphere-specific white matter (WM) regions of interest (ROIs) based on a probabilistic WM atlas (JHU) (52) for each of the 26 metrics. Altogether, 1,794 diffusion features were derived per individual [26 metrics *×* (48 ROIs + 20 tracts + 1 global mean value)].

### C. Polygenic Risk Scores

We estimated PGRS for each participant with available genomic data, using PRSice2 (53) with default settings. As input for the PGRS, we used summary statistics from recent genome-wide association studies of ASD (15), MDD (19), SCZ (22), ADHD (16), BIP (21), OCD (17), ANX (18), and AD (20). We used a minor allele frequency of 0.05, as most commonly used threshold across PRS studies of psychiatric disorders.

While psychiatric disorders were *p*-values thresholded at *α* = 0.05 (15–23), recommendations for AD (*α* = 1.07^−4^ (54)) lead to the application of a lower threshold of *α* = 0.0001, with the goal of optimising signal to noise in comparison to previously used *α* = 0.001 (55). The goal with the estimation of the PGRS was to relate cross-sectional WMM metrics and WMM changes to disease-related genetic profiles to examine to which degree these genetic risk profiles can explain WMM changes in midlife to senescence.

### Statistical Analyses

All statistical analyses were carried out using R version 4.2.0 (www.r-project.org), and FSL version 6.0.1 (48).

First, we assessed unadjusted time point differences by using paired samples *t*-tests on a set of cognitive measures, and each of the global and regional scalar diffusion metrics *F* (such as FA from DTI) and present Cohen’s *d* indicating the effect size:

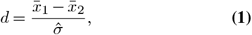

describing the difference between means 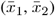 over an estimate of the sample standard deviation of the data 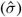.

We then used linear mixed effects regression models (LMER) adjusting *F* for age, sex, the sex-age interaction (sex *×* age), time point (*TP*) and scanner site (*Site*). *ID* was treated as random intercept (*RRI*), and *y* the y-intercept:

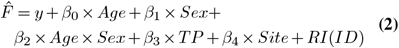

For comparison of age-relationships between time points, we utilised simple linear and generalised additive models (GAMs) on each single time point (treating the data as crosssectional), with the linear models taking the following form:

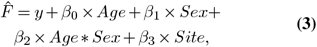

and the GAMs this form (including a spline function of age *S*(*Age*) to model non-linear associations):

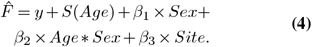

We also estimated the annual rate of change (*ARoC*) of each global feature by taking the difference in WMM features *F* between time points over the time passed between time points (scan1&2) indicated by the Δ*age* = *age*_*scan*2_ − *age*_*scan*1_:

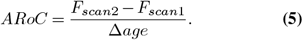

Global *ARoC* was corrected for sex, sex *×* age, and scanner site (as *RI*):

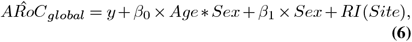

for which then age-correlations were estimated. For regional features, we used simple linear models to support model convergence:

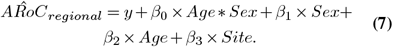

For the voxel-level analysis, we estimated one-sample *t*-tests on the contrast between each time point’s maps within the FA skeleton accounting for age, Δage, scanner site and sex using FSL randomise with 10,000 permutations (H_0_: Difference = 0).

Finally, we assess the associations between *ARoC* and *PGRS* for global and regional WMM metrics adjusting for age, age*×*sex, and site using simple linear models:

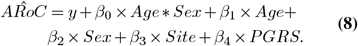

For voxel-level analyses, we used TBSS randomise with permutation-based statistics (running 10,000 permutations) (**?**). Mean maps and between-time-point contrast maps served for the computation of one-sample t-tests for each of the observed metric, while accounting for age, sex and site (i.e., random intercept models):

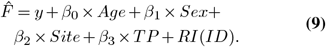

*P*-values were adjusted for multiple comparison using Bonferroni-correction(56) for global and region-averaged metrics, and family-wise error (FWE)-corrections were used for voxelwise inferential statistics (using Threshold-Free Cluster Enhancement(57)). We report conditional variance explained in the main text 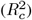, which refers to variance explained by fixed factors.

## Results

### Cognitive changes

Our analysis suggested no significant time point differences in cognitive measures for the inter-scan interval of Δ =2.44±0.73 (Supplement 16).

### Global WMM changes

The globally averaged WMM metrics differed between time points (Cohen’s 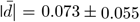, *d*_*min*_ = −0.248, *d*_*max*_ = 0.159), with the exception of BRIA - micro ADC, all SMT metrics, and SMTmc - diffusion coefficient (Supplement 3-4).

Congruently, LMER (Eq. 2) outline the effect of time point, in addition to age and sex (Supplement 5a-b). While the effects of time point (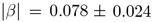, *β*_*min*_ = −0.107, *β*_*max*_ = 0.113), age (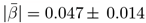, *β*_*min*_ = −0.063, *β*_*max*_ = 0.067), and sex *×* age (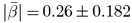, *β*_*min*_ = −0.619, *β*_*max*_ = 0.197) were significant (*p* < .001; Supplement 6-7), and modelling diffusion metrics well 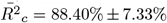 (Supplement 8), only two diffusion metrics showed significant sensitivity to sex (BRIA micro FA: *β* = −0.642, *p* = 0.034, DKI radial kurtosis: *β* = −0.785, *p* = 0.022, both lower in males)), and presented large variability (Supplement 5c-d, Supplement 9-10). Moreover, WMM metrics’ age-associations were better modelled as non-linear 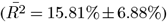 than linear associations 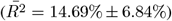, considering variance explained (Fig. 1). Yet, this difference in model fit was non-significant (*p* = 0.557). We show decreases in FA, and kurtosis (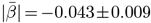, *β*_*min*_ = −0.063, *β*_*max*_ = −0.035). At the same time, water diffusivity and extra-axonal water fraction, including free water (mainly cerebro-spinal fluid) increase (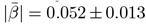, *β*_*min*_ = 0.025, *β*_*max*_ = 0.067). Notably, the intra-axonal water fraction, estimated by different diffusion approaches, exhibit the same behaviour independent of the diffusion approach (lower at higher ages). Finally, we present accelerations of the annual rate of change (ARoC) at higher ages for most global WMM metrics (Fig. 2). Large age-group differences were observed at Cohen’s 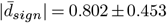, |*d*_*min*_| = 0.015, |*d*_*max*_| = 1.920 (when including also non-significant findings: 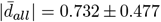, |*d*_*min*_| = 0.010, |*d*_*max*_| = 1.92 (see Supplement 18 for details on test statistics, including effect size estimates). Decelerating ARoC were observed for BRIA-DAX intra and extra, and DKI-AK, and relatively stable ARoC for SMT - longitudinal coefficient. See also Supplement 19 for ARoC-age-trends indicating accelerated ageing at 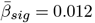, Supplement 20 for uncorrected age-stratification, Supplement 21 for corrected sex-stratification, respectively.

**Fig. 1.**
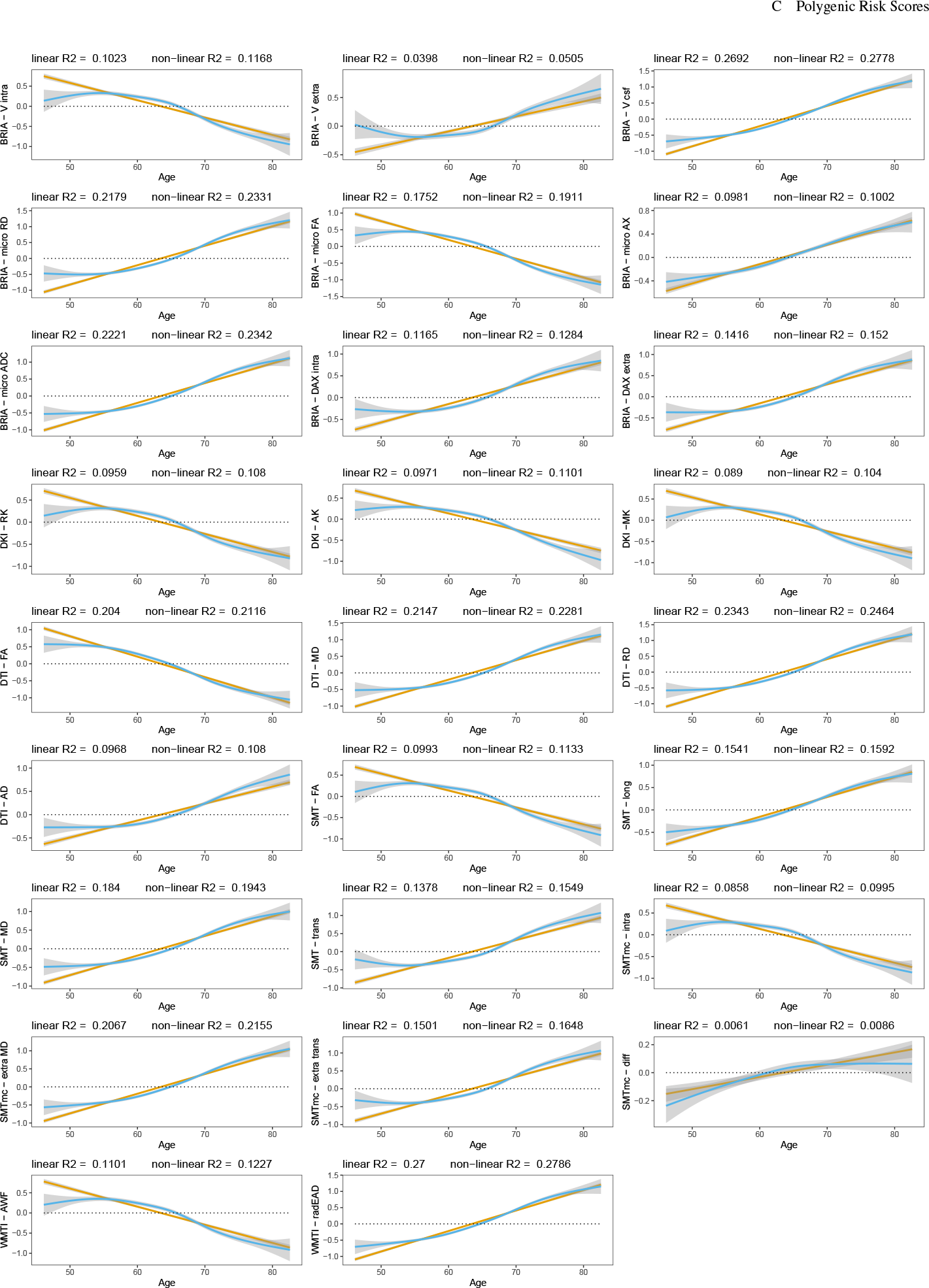
Global WMM ageing trajectories. WMM values were standardised, mean-centred, and adjusted for covariates of no interest using LMERs. Second, age-WMM relationships were described by linear and non-linear functions.

**Fig. 2.**
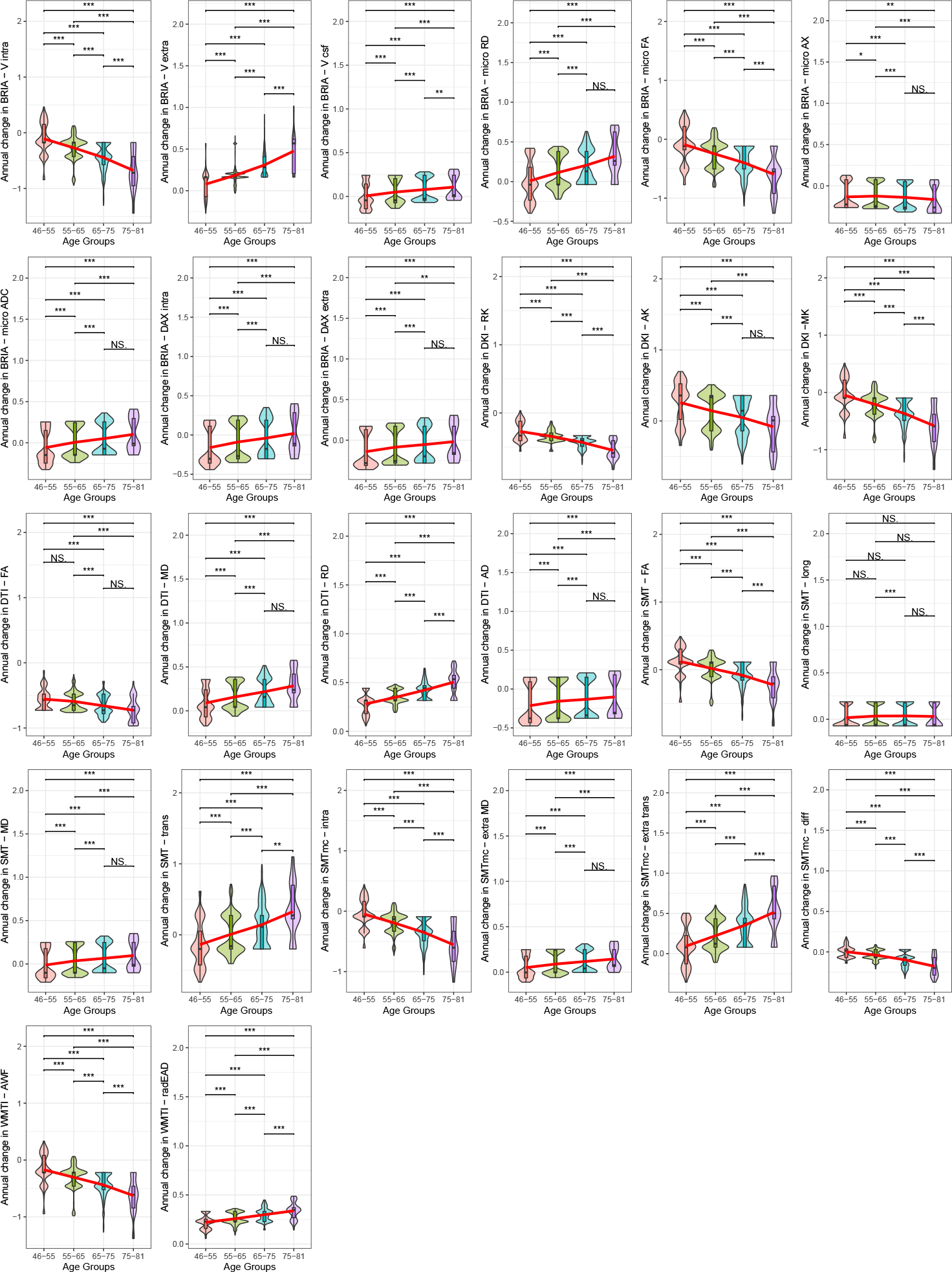
Age-stratified annual WMM change. WMM were corrected for age, sex, age*×*sex and site, and standardized for comparability (without mean-centering). We present *p*-values for Wilcoxon tests, which were significant at the Bonferroni-corrected * *α* < 0.05/(26*6) = 3.21 *×*10^−4^, ** *α* < 0.01/(26*6) = 6.41*×*10^−5^, *** *α* < 0.001/(26*6) = 6.41*×*10^−6^, NS = non-significant. The red lines were added as a visual help to identify trends of accelerated or decelerated annual change.

### Regional white matter microstructure changes

Estimating paired-samples tests indicate that 47.57% of white matter features decreased (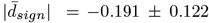, |*d*_*min*_| = −0.825, |*d*_*max*_| = −0.030), 32.30% increased (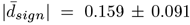, |*d*_*min*_|= 0.283, |*d*_*max*_| = 0.516) and 20.13% did not change between time points. Most extreme *p*-values among these unadjusted time-point differences were observed in Fornix (DTI-RD: 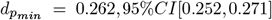 and the body of the corpus callosum (DTI-FA: 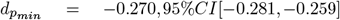, and largest effect sizes of Cohen’s |*d*| *>* 0.5 found in the middle cerebellar peduncle (BRIA microAX: *d*_*max*_ = −0.825, 95%*CI*[−0.875, −0.775]; Fig. 3a).

**Fig. 3.**
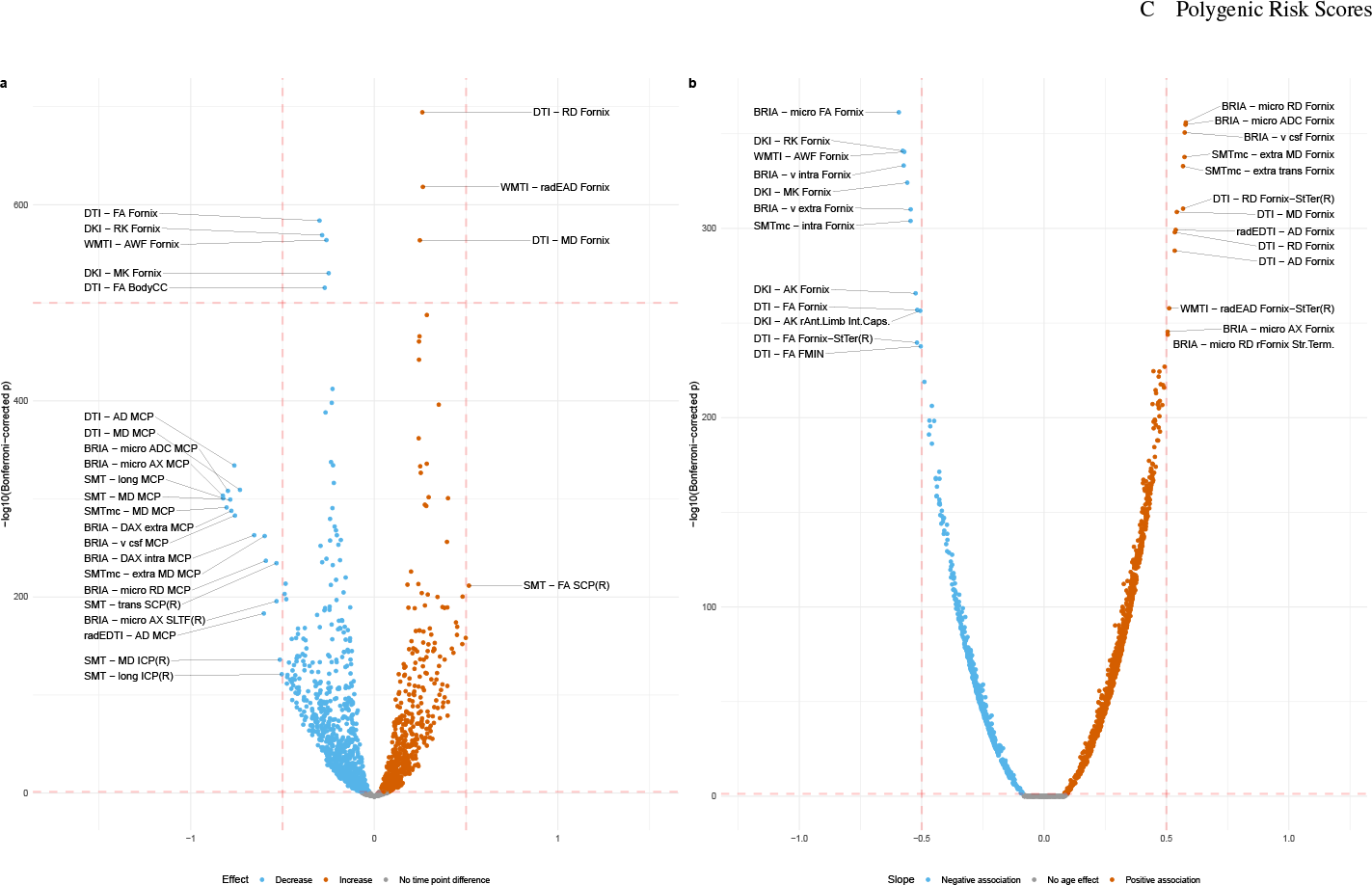
Regional white matter microstructure changes between time points and age-associations. Panel (a) presents unadjusted effect sizes (Cohen’s *d*) versus Bonferroni-adjusted -log10 *p*-values. Labelling was done using a medium effect size threshold of Cohen’s |*d*| *>* 0.5 (also marked with vertical lines) as well as extreme Bonferroni-adjusted *p*-values of −10*log*(*p*) *>* 500. Panel (b) presents adjusted WMM associations with age. Age-WMM were adjusted for sex, sex*×*age, scanner site and time point (Eq. 2). The plot presents standardised slopes (*β*) versus Bonferroni-adjusted -log10 *p*-values. Labelling was done using a large association of |*β*| *>* 0.5 (also marked with vertical lines). BodyCC = body of the corpus callosum, SCP(R) = right superior cerebellar peduncle. MCP = middle cerebellar peduncle, ICP = inferior cerebellar peduncle. SLTF(R) = right superior longitudinal temporal fasciculus. StTer(R) = right stria terminalis. Dotted lines were inserted as visual aid: the lower horizontal dotted line represent the significance level of *p* = .05 and the upper horizontal line −10*log*(*p*) = 500. The vertical lines represented labelling borders based on a medium effect size of Cohen’s |*d*| *>* 0.5 (panel a) and large associations of |*β*| *>* 0.5 (panel b). Tables with test statistics are available at https://github.com/MaxKorbmacher/Long_Diffusion/.

LMER (Eq. 2), outlined strongest age-associations in fornix WMM (|*d*| *>* 0.5; Fig. 3b, see Supplement 13 for distribution of *β*-values). We also identify various regions sensitive to sex differences and smaller sex age-interaction effects, with repeated occurrence of significant fornix, cerebral peduncle, corticospinal tract, and medial lemniscus differences (Supplement 11,12). Across regions, time point was a non-significant fixed effect (*β <* 1.5*×* 10^−5^, *p >* .05). For comparability across regions, we used |*ARoCs*| in association with age, adjusting for *age × sex*, sex, and site effects, showing accelerated ageing (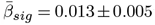; Supplement 17).

### Voxelwise white matter microstructure changes

When investigating voxelwise changes of WMM between time points (adjusted for age, sex, and scanner site), we find two major patterns examining both increases and decreases in WMM: 1) a global decrease in fractional anisotropy metrics (with the exception of SMT-FA showing distributed changes, including frontal increase and posterior decrease), and 2) an overall decrease in radial, axial and mean diffusivity metrics in orbitofrontal, occipital and brain-stem and cerebellum and increase in superior frontal areas (Table 1). For an overview of the voxelwise WMM maps at corrected *α* = 0.05 see Supplement 25.

**Table 1.**
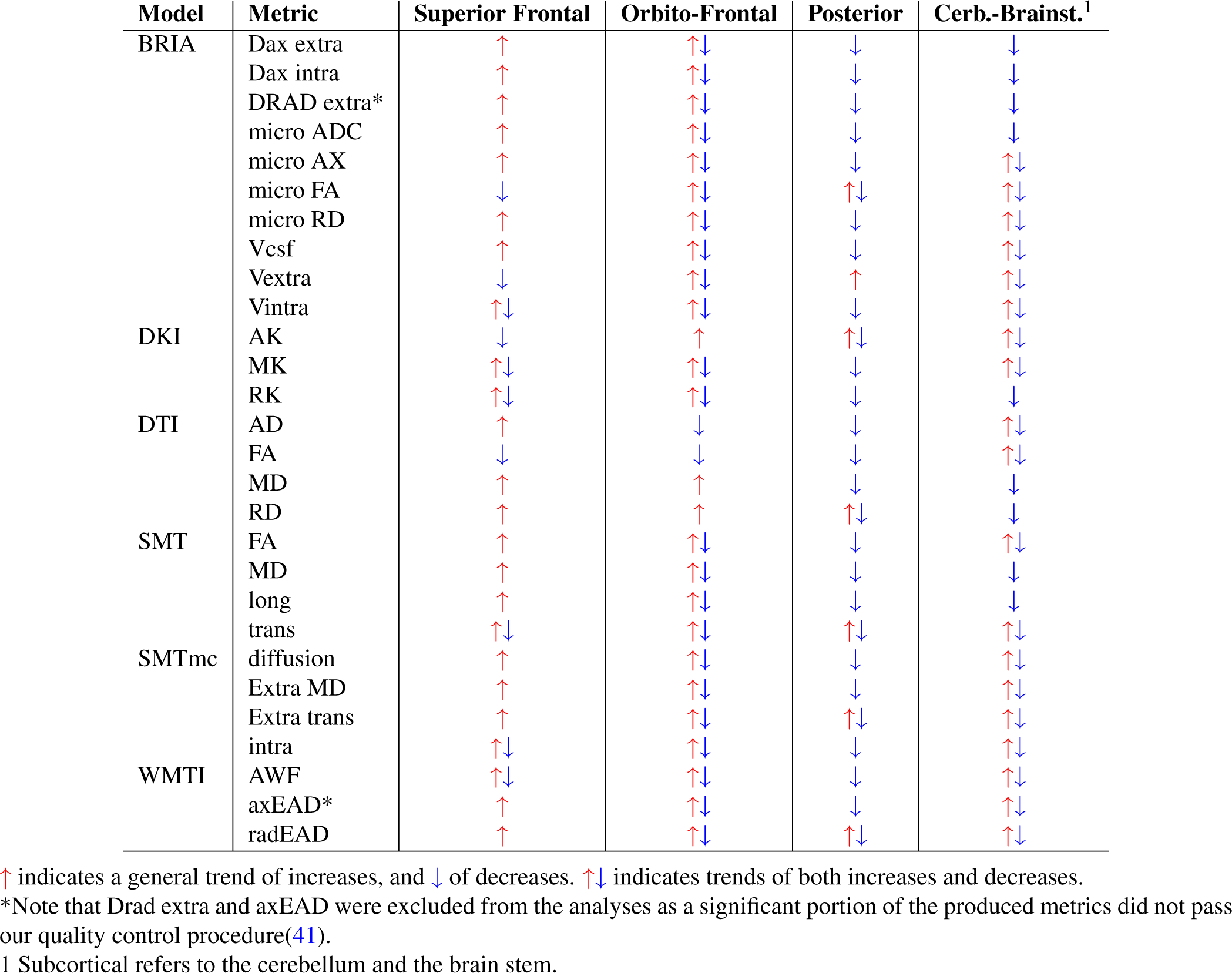
Trends in WMM changes.

### PGRS Associations

Although annual change in the cerebral peduncle showed the strongest associations with PGRS of AD and global WMM with ADHD, both global 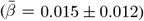 and regional 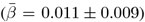 WMM-PGRS associations were non-significant after adjusting the *α*-level for multiple comparisons (Fig. 4a-b). Nevertheless, a highly brain-region-specific pattern of associations between WMM ARoC and PGRS was observed (Fig. 4b) with the medial cerebral peduncle (Fig. 4c) showing the strongest consistent associations with PGRS and specifically associations with AD (|*β*_*max*_| = 0.053, 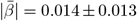), and MDD (|*β*_*max*_| = 0.051, 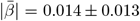; Fig. 4b-c, see for all peduncle-PGRS associations Supplement 22, indicating strongest ARoC-PGRS 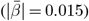 associations but weaker non-replicating associations for cross-sectional assessments 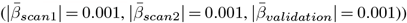). Fornix, the most age-sensitive region related strongest to ANX (|*β*_*max*_| = 0.011, 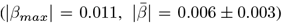), and OCD PGRS (|*β*_*max*_| = 0.008, 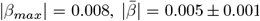; Supplement 23).

**Fig. 4.**
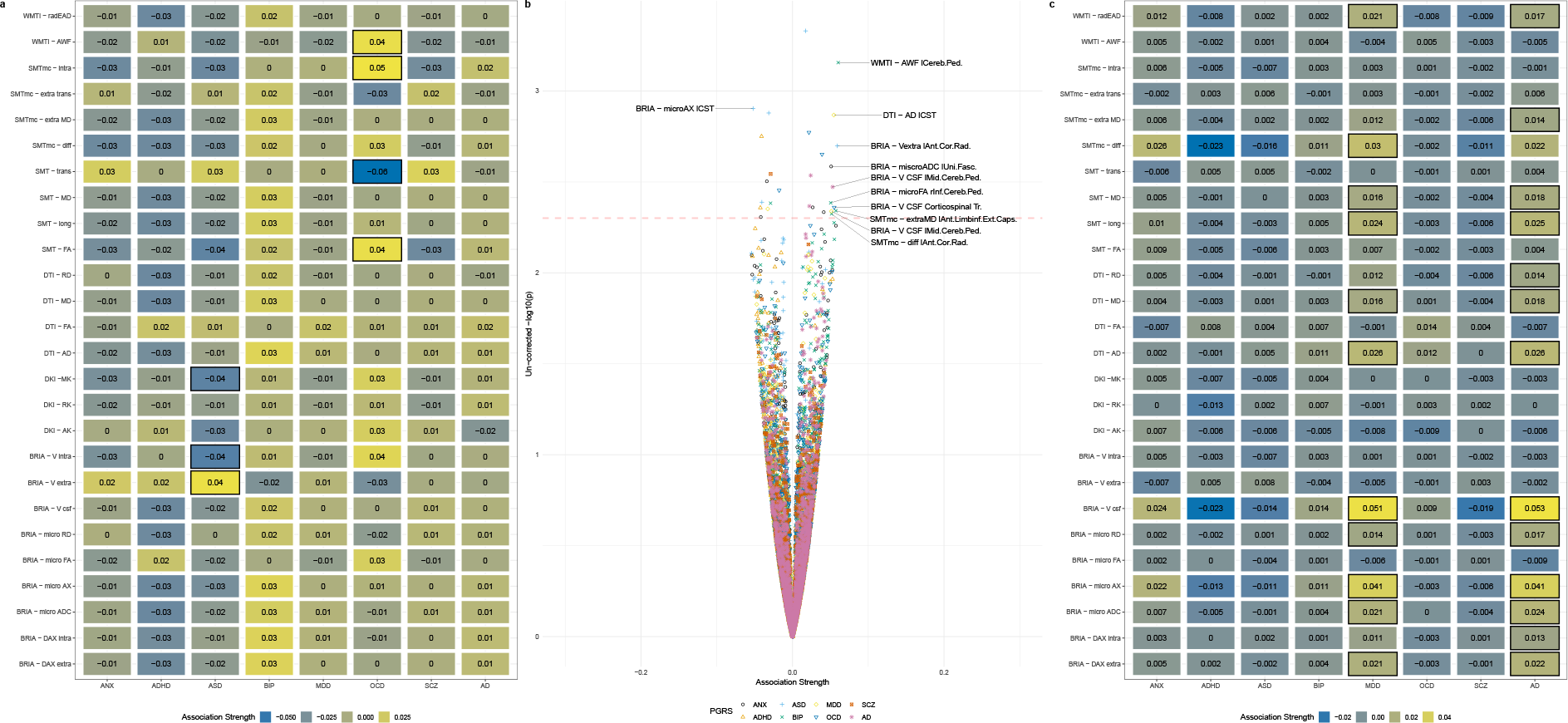
Associations of PGRS with the rate of white matter change. Panel (a) presents the global associations between PGRS and WMM change. Colours indicate the strength of association (standardised *β*-coefficients). Panel (b) presents the regional associations between PGRS and WMM change. The dotted line indicates an uncorrected *α <* 0.001. Labels presenting the respective regions and metrics are supplied above this *α*-threshold, as well as at |*β*| *>* 0.05. Panel (c) presents the regional associations exclusively for the medial cerebral peduncle, the region where the strongest ARoC-PGRS associations were observed. Boxes in panels (a) and (c) indicate the statistical significance at an uncorrected *α <* 0.05. *Note:* All associations were adjusted for age, sex, the *age × sex* interaction, and site. None of the presented associations survived the adjustment of the *α*-level for multiple comparisons.

Similarly, observing each time point separately, both global and regional WMM were non-significant after adjusting for multiple comparisons. However, WMM-PGRS associations were highly similar at the two time points in the longitudinal sample, yet highlighted BIP and SCZ associations (considering *p*_*uncorrected*_ *<* 0.05, Fig. 5a-b), did however not replicate in an independent cross-sectional validation sample of *N* = 31, 056 UKB participants (Fig. 5c). Noteworthy, WMM-PGRS associations were strongest for AD 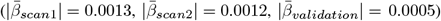 and ANX 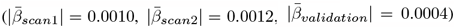 in the longitudinal sample, but strongest for ADHD 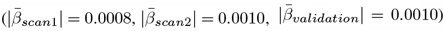, and ASD in the cross-sectional validation sample 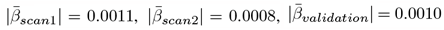, Fig. 5d-f).

**Fig. 5.**
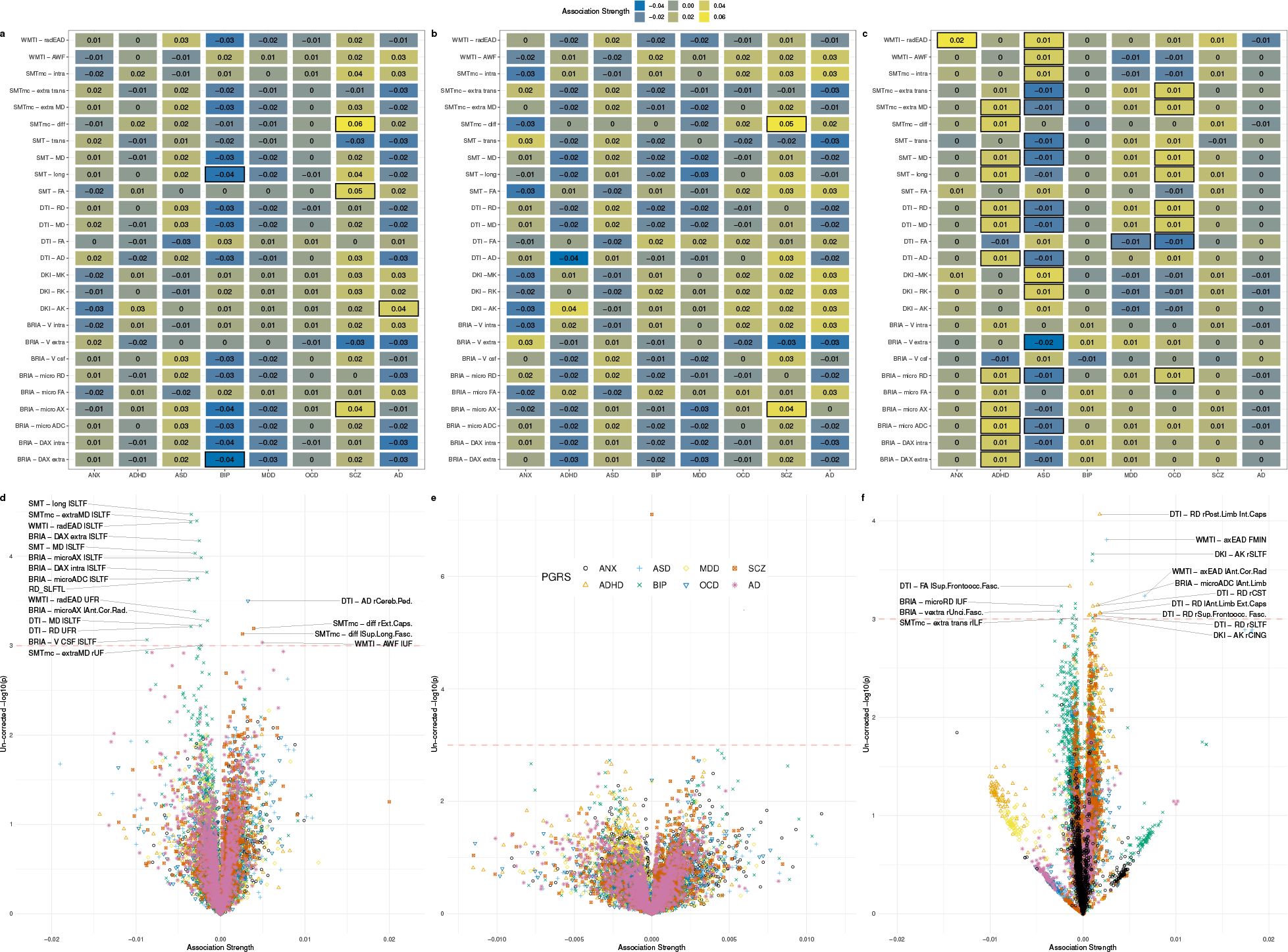
Cross-sectional associations between PGRS and WMM in longitudinal and cross-sectional validation data. Panel (a) shows PGRS-associations of globally averaged WMM for time point one in the longitudinal sample *N* = 2, 329, and panel (b) for time point two, respectively. Panel (c) shows the global WMM-PGRS associations for the cross-sectional validation sample *N* = 31, 056. Boxes indicate significance at an uncorrected *α <* 0.05. For simplicity, standardised regression coefficients with |*β*|*<* 0.005 were rounded down to *β* = 0. Panel (d) shows PGRS-associations of regionally averaged WMM for time point one in the longitudinal sample, and panel (e) for time point two. Panel (f) presents the regional associations for the cross-sectional validation sample. The dotted line in panels (d-f) indicates an uncorrected *α <* 0.001. Labels presenting the respective regions and metrics are supplied above this *α* threshold, and |*β*| *>* 0.001. *Note:* All associations were adjusted for age, sex, *age × sex* as fixed effects, and site as random effect, and none of the associations survived the adjustment of the *α*-level for multiple comparisons.

Overall, ARoC-PGRS association 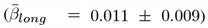 were significantly stronger than cross-sectional effects 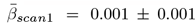, *d* = 0.028, 95%*CI*[0.004, 0.051], *p <* .001, 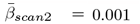, *d* = 0.001 *±* 0.001, *d* = 0.026, 95%*CI*[0.003, 0.049], *p <* 0.031, 95%*CI*[0.008, 0.054], *p <* .001 (see Supplement 24 for distribution of effects).

## Discussion

We investigated WMM changes using longitudinal UKB diffusion data using a series of diffusion approaches and their associations with PGRS. The comparison between two time points, with an average inter-scan interval of 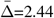 years, was performed at different spatial scales in order to localise the strongest ageing effect and its correlations with other covariates. Unadjusted time point differences in global WMM metrics (Supplement 3) resembled closely the covariate-adjusted time point differences, as well as age effects (Supplement 5a-b). These ageing effects largely confirmed previous findings showing a global decrease of fractional anisotropy and the increase in axial, radial and mean diffusivity for both intra- and extra-axonal space(3, 6, 40, 58– 60), whereas kurtosis metrics decrease with age(3, 6). Noteworthy, we provided evidence for the intra-axonal water fraction estimated by different diffusion approaches demonstrating age-related decrease, accompanied by opposite trends for the extra-axonal water fraction and the CSF water fraction (also known as *free water* in other diffusion approaches).

We observed accelerated change in global and regional WMM at higher ages. Observed inconsistencies in age dependencies of the annual change in axonal diffusivity stand in contrast to a previous longitudinal study analysing DTI data on the voxel-level(36). Such difference in the localisation of the increased acceleration and in findings on axial diffusivity might be driven by methodological variations: here, we focused on global and regional averages of annual WMM change. Yet, in contrast to the mentioned study(36), another longitudinal investigation did also not detect accelerated axial diffusivity changes(61). However, our study provides larger statistical power than previous longitudinal investigations on the rate of change in WMM (e.g.,(36, 61)), and is hence bench-marking global and regional increases in the rate of WM change at a higher age.

Region-wise investigations allowed for more differentiated results and presented larger ageing effects compared to global WMM metrics (Fig. 3, Supplement 13). Region-level assessments further underlined the strong effect of age on WMM(3, 6, 40, 60, 61). Differences between unadjusted and adjusted associations gave more detail to dependencies of regional WMM age associations on sex and scanner site. Unadjusted associations outlined the cerebellar peduncle, and the superior longitudinal temporal fasciculus as regions with largest age-association, and corpus callosum and fornix as statistically most significantly age-associated regions. After corrections, the fornix was identified as the most significant and strongest age-associated region, which is congruent with previous cross-sectional findings on limbic WMM age associations(3, 4, 40, 62). These WMM age associations also followed the described global pattern of fractional anisotropy metrics decreasing at higher ages, and axial, radial, and mean diffusivity metrics increasing during ageing.

The observed regional fornix changes were moreover clearly delineated in the voxel-level analysis. Additionally, a more general pattern was observed: radial, axial and mean diffusivity metrics increased in superior frontal areas but decreased in more posterior and inferior areas (Table 1, Supplement 25). While previous findings show consistent DTI AD, RD and MD increases and FA decreases across the brain for global and tract measures throughout ageing(60, 63), or particularly in the brain stem(36), no study has yet outlined such differential, non-homogeneous spatial patterns of WMM changes across the brain. Only one other study(64) revealed a similar pattern for DTI when examining cross-sectional AD, RD, and MD tract-age-associations in the UKB, and recent reviews which highlight the frontal lobes as most susceptible to white matter deterioration(65, 66). Potentially, our findings provide additional evidence for the “last-in-first-out” retrogenesis hypothesis, which states that brain areas that develop slowest (such as the prefrontal cortex) are more vulnerable to negative ageing effects, such as degeneration(67). On the other hand, the observed WMM changes might simply map onto frontal GM areas which are most affected by normal ageing processes instead of areas with higher evolutionary expansion(67). Yet, as the age-dependence of frontal WM seems to be partly explained by cognitive ability(68), and cerebrovascular factors(69), frontal WM might be particularly interesting for further clinical dMRI examinations.

In accordance with our previous findings(3, 40), we found that the fornix is highly sensitive to ageing-related changes. The fornix – a C-shaped bundle of nerve-fibres that acts as a major output tract of the hippocampus – is a brain region implicated in various neurological and psychiatric disorders, such as, mild cognitive decline(70), impairment(71, 72), Parkinson’s disease(73), arguably Alzheimer’s disease (compare(74–78), and bipolar disorder(79). Moreover, the genetic architecture of fornix WMM is related to various neurological and psychiatric disorders(80). Bridging such genetics and imaging findings indicates that there are genetic underpinnings for the accelerated ageing of the fornix and other regions, which might explain pathology development. While our findings render the fornix as a promising marker of ageing, future studies need to explore this region as potential therapeutic target.

We identified general patterns of WM changes when applying diffusion approaches at voxelwise scale; namely, 1) global fractional anisotropy metric decreases, and 2) superior axial, radial and mean diffusivity metric increases in superior frontal brain regions, but 3) decreases in posterior regions, the brain stem and the cerebellum. Fractional anisotropy (DTI-FA and BRIA-microFA) decreases suggest different potential biological processes such as a seizing myelination or cell death across the brain (with microFA adding information on the fibre orientation coherence(81), Table 1, Supplement 25) or axonal degradation(82, 83) in combination with intra-axonal water fraction metric. The other diffusion metrics contain information on axial, radial and mean diffusivity and coherently suggested WMM degeneration with increasing age in superior frontal lobes and potential cell-swelling in posterior and subcortical regions where diffusivity decreases. This is congruent with multi-compartment metrics from BRIA, SMT, and WMTI (see Supplement 25). Potentially, the decreases in diffusivity metrics in posterior and subcortical regions depict compensatory mechanisms, accounting for the frontal lobe deterioration. Additional examination of diffusion properties (e.g., using tractography) leveraging both single and multi-shell dMRI might provide further insight into the differential developments of white matter across the adult lifespan.

Finally, this study presents a comprehensive overview of WM association with PGRS of common psychiatric disorders and Alzheimer’s Disease. Moreover, we differentiated between global and regional WM metrics at each time point and the annual rate of change, which led to different association patterns. However, the associations of both WM and annual WM-change with PGRS were non-significant when correcting for multiple comparisons. Previous results demonstrate small associations between WM and PGRS in the UKB for MDD(39), SCZ, BIP(84), and AD(38), as well as in the Adolescent Brain and Cognitive Development Study(24). For the further highlighting of key associations, we considered uncorrected *p*-values, examining the PGRS-WM associations as suggestive for potential associations of WM and genetic risk.

Surprisingly, the global annual rate of WMM change associated only with ADHD PGRS. WMM at each time point provided a more nuanced association pattern including ANX, OCD, and AD, similar to previous findings on AD PGRS-WM associations(38). Additionally, similarities between ANX and OCD PGRS associations with WMM might originate from large symptom overlap between these diagnoses(85). This association pattern was however not replicated in an independent cross-sectional portion of the UKB which instead outlined associations with ASD and BIP. Hence, whether these differential association patterns speak to sample-specific gene-brain relationships, or are simply noise due to a lack of statistical power (as a function of small effect sizes) requires follow-up with larger samples, more time points, and larger inter-scan-intervals.

Observing relationships of PGRS with both WMM and its annual rate of change in single regions highlight the brain stem, cerebral peduncle, and the limbic system as potential PGRS-association targets. Importantly, PGRS associations were orders of magnitude stronger for WMM rate of change than for cross-sectional metrics, which underlines the importance of examining the genetic underpinnings of WM in longitudinal data. Notably, areas outlined as most age-sensitive (the fornix and cerebral peduncle) were also the strongest related in their annual change to ANX, ADHD, OCD, and SCZ PGRS. Yet, more longitudinal research is needed to validate the presented findings. Genetic overlaps between fornix WMM and the listed disorders(80), as well as the involvement of the cerebellum, which is connected with the cortex via the cerebral peduncles, in various psychiatric disorders, gives additional insight into the role of genetic makeup for WMM development(86). Furthermore, the cerebral peduncles were particularly associated with AD, ADHD, and OCD PGRS, but also with ANX, BIP, MDD, and SCZ PGRS. This spatially specific pattern of PGRS associations emphasises the usefulness of regional investigations, due to highly spatially distributed influence of genetics. While the small effect sizes limit inferences on WMM-PGRS associations, the highlighted associations of PGRS and WMM *change* are worth further investigation.

There are several limitations to be mentioned in the context of this study. First, the age range was limited to individuals older than 40 years, allowing only for generalisations across mid-to-late adulthood. Future studies should consider large samples to cover the whole lifespan, particularly when the objective is to investigate WM ageing or to investigate generalisable associations of WM with genotypes and phenotypes. Second, the inter-scan interval was relatively short 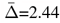 years). Longer inter-scan intervals might reveal clearer information on accelerated WMM ageing processes at different ages and their genetic underpinnings. Longer intervals are also useful to examine the relationship of cognitive decline and WMM. Third, we used a relatively homogeneous, non-diverse sample including nearly exclusively white UK citizens, which limits the generalisability beyond white Northern Europeans and US Americans in their midlife to older age. Additionally, although the sample size was larger than in previous longitudinal WMM investigations, power was still limited to find PGRS associations (presenting small effects). The volunteer-based sampling of the UKB participants might additionally have introduced bias, reducing generalisability to the UK population(87). Yet, the imaging sample of the UKB shows an additional positive health bias (better physical and mental health) over the rest of the UKB sample(88), rendering this sub-sample as even less representative of the UK population. However, this might not necessarily be a disadvantage considering the objective of this study, which was to map WM change in healthy mid-to-late life adults and associate polygenic risk with the observed WM changes. Fourth, conservative corrections of the *α*-level using Bonferroni corrections might have led to false negatives, especially for the small observed effects of PGRS on the annual change in regional WMM where many associations were explored. Finally, the fornix, the key region of the presented regional WMM changes, is a region susceptible of free water contamination due to its closeness to the cerebrospinal fluid(2) and has previously been suspected of partial volume effects(62). Hence, findings on this region need to be interpreted carefully.

To conclude, our findings provide insight about short-term WM changes indicating degradation processes indicated by lower FA, kurtosis, intra-axonal water fraction, and higher diffusivity, free water, and extra axonal water fraction. These changes are associated with the de-myelination and structural disintegration across the adult brain as a consequence of ageing without strong or detectable cognitive decline. Demyelination and WM features degradation primarily affect frontal brain regions, whereas posterior regions as well as brain stem and cerebellum show opposing trends. Further investigations should focus on fornix WMM *changes* throughout the lifespan to investigate health and disease outcomes, and the role in such of the genetic architecture of the cerebellar peduncle in such white matter changes and disease development.

## Data Availability

Data are available from the UK Biobank (https://www.ukbiobank.ac.uk/).

## AUTHOR CONTRIBUTIONS

Max Korbmacher: Study design, Software, Quality control, Formal analysis, Visualizations, Project administration, Writing—original draft, Writing—review & editing. Dennis van der Meer: Software, Writing – review & editing. Eli Eikefjord: Funding acquisition. Dani Beck: Writing—review & editing. Daniel E. Askeland-Gjerde: Writing—review & editing. Ole A. Andreassen: Writing—review & editing, Funding acquisition. Lars T. Westlye: Writing—review & editing, Funding acquisition. Ivan I. Maximov: Supervision, Study design, Software, Data preprocessing and quality control, Writing—review & editing, Funding acquisition.

## ACKNOWLEDGEMENTS

This study has been conducted using UKB data under Application 27412. UKB has received ethics approval from the National Health Service National Research Ethics Service (ref 11/NW/0382). The work was performed on the Service for Sensitive Data (TSD) platform, owned by the University of Oslo, operated and developed by the TSD service group at the University of Oslo IT-Department (USIT). Computations were performed using resources provided by UNINETT Sigma2 – the National Infrastructure for High Performance Computing and Data Storage in Norway. Finally, we want to thank all UKB participants and facilitators who made this research possible.

This research was funded by the Research Council of Norway (#223273, #300767); the South-Eastern Norway Regional Health Authority (#2022080, #2019101); and the European Union’s Horizon2020 Research and Innovation Programme (#847776, #802998).

## CONFLICTS OF INTEREST

OAA has received a speaker’s honorarium from Lundbeck and is a consultant to Coretechs.ai.

## DATA AND CODE AVAILABILITY

All raw data are available from the UKB5 (www.ukbiobank.ac.uk). Analysis code and extended supplementary files will be made available at the time of publication at https://github.com/MaxKorbmacher/Long_Diffusion.

## Supplementary Note 1 The utilized white matter quality control pipeline

In brief, YTTRIUM (41) converts dMRI scalar metrics into 2D format, using a structural similarity (89, 90) extension of each scalar map to their mean image in order to create a 2D distribution of image and diffusion parameters. These quality assessments are based on a 2-step clustering algorithm applied to identify subjects located out of the main distribution. Additionally, rows including impossible values, such as diffusion coefficients *d*: 0 *< d <* 4 *μ*m^2^ *·* ms, kurtosis values *K* 0 *< K <* 3, and FA values 0 *< FA <* 1 were excluded. QC of the mean skeleton values rendered *N* = 622 datasets of the BRIA metric extra-axonal radial diffusivity (DRADextra) as such impossible values, and examining regional and tract averages *N* = 643 of the WMTI metric axial extra-axonal diffusivity (axEAD). Due to the relatively large share of these outliers on the total sample, we excluded these metrics from the analyses.

## Supplementary Note 2 Description of white matter features by diffusion approaches

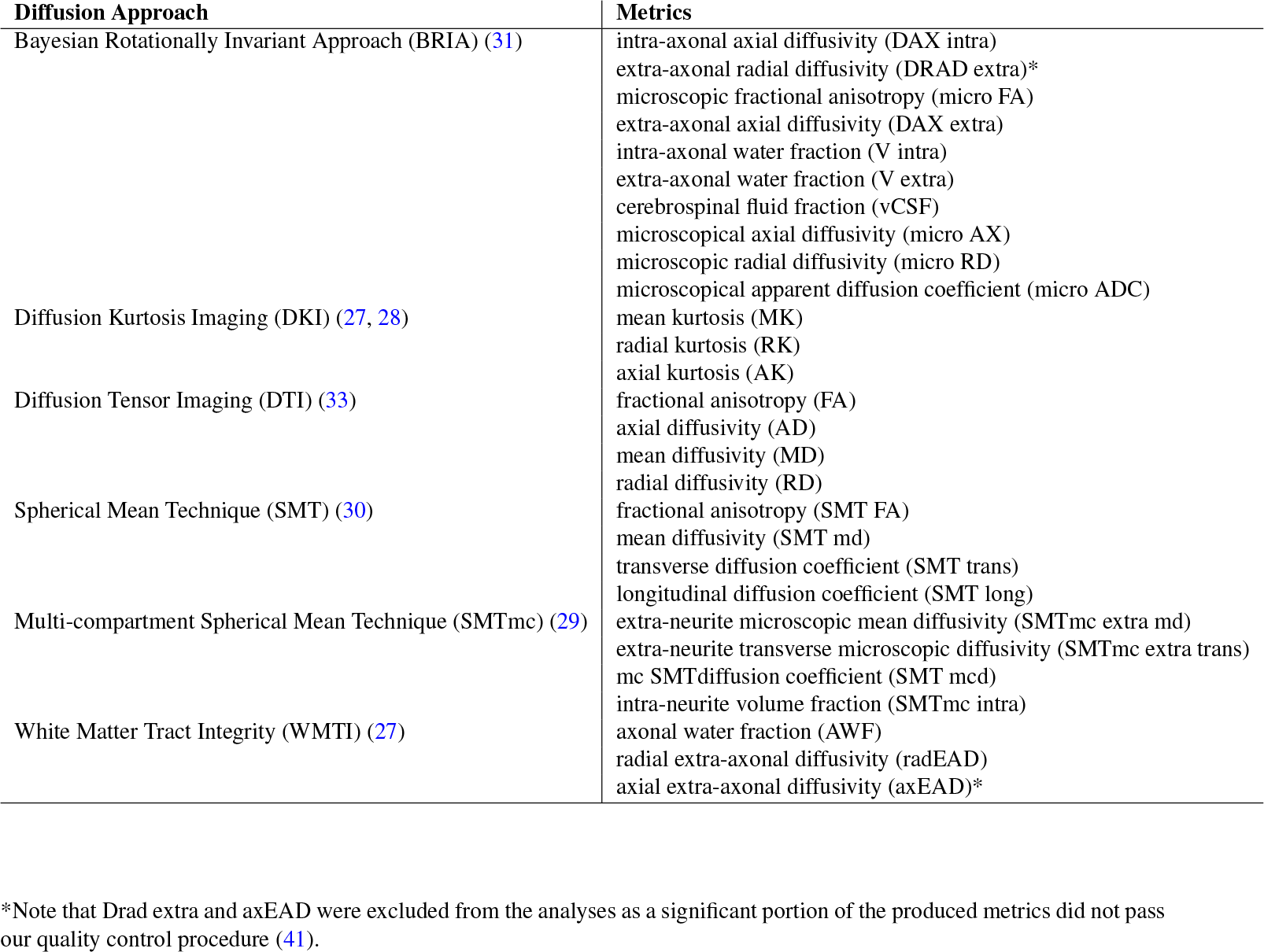

## Supplementary Note 3 Effect sizes for unadjusted microstructure changes between time points

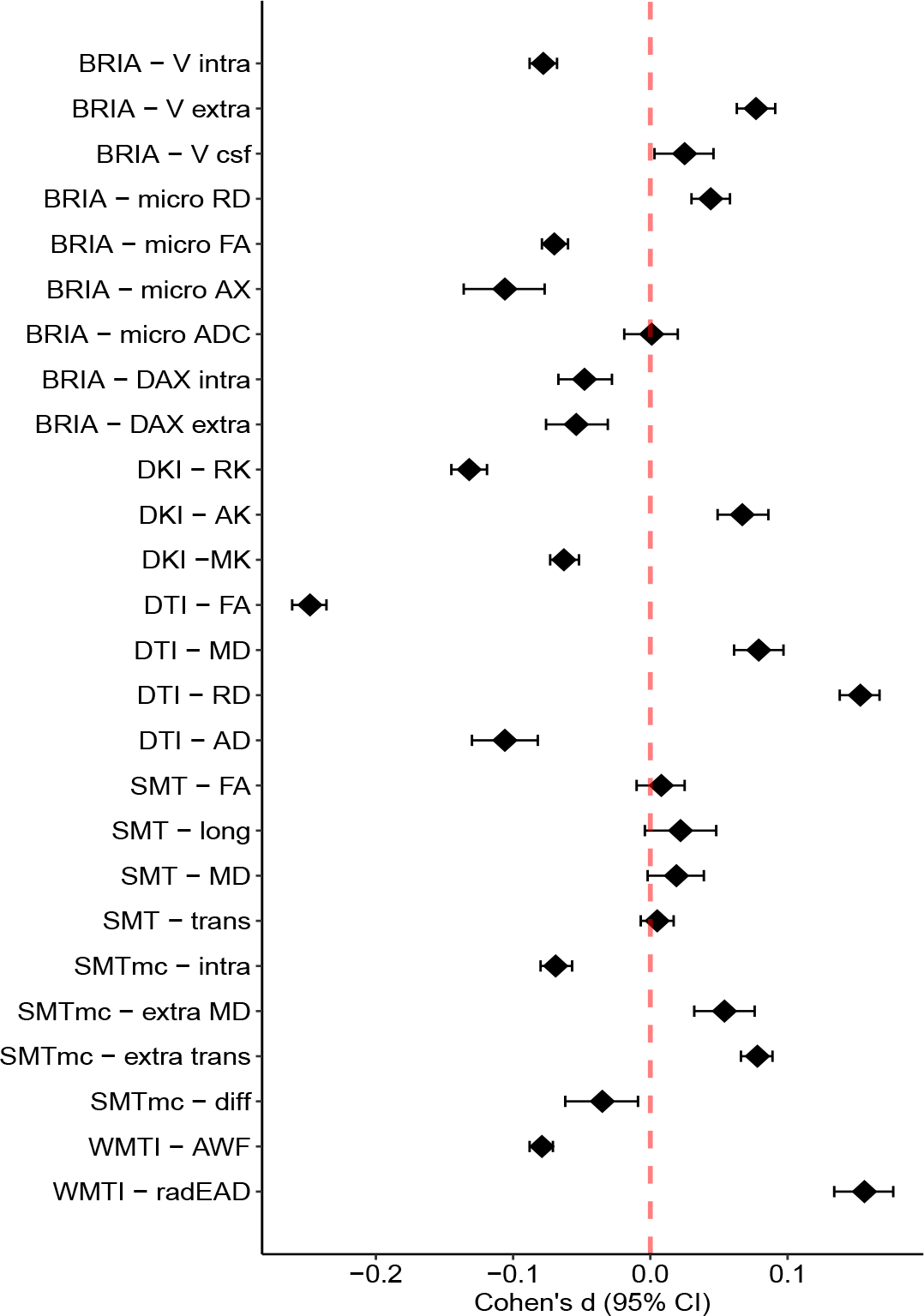

## Supplementary Note 4 Test statistics for time point differences for global diffusion metrics

The table presents mean (M), standard deviation (SD) for each time point (TP), T-statistic (T), uncorrected p-value (p), Bonferroni-corrected p-value *p*_*Bonf*_, and Cohen’s d with 95% CI as effect size.

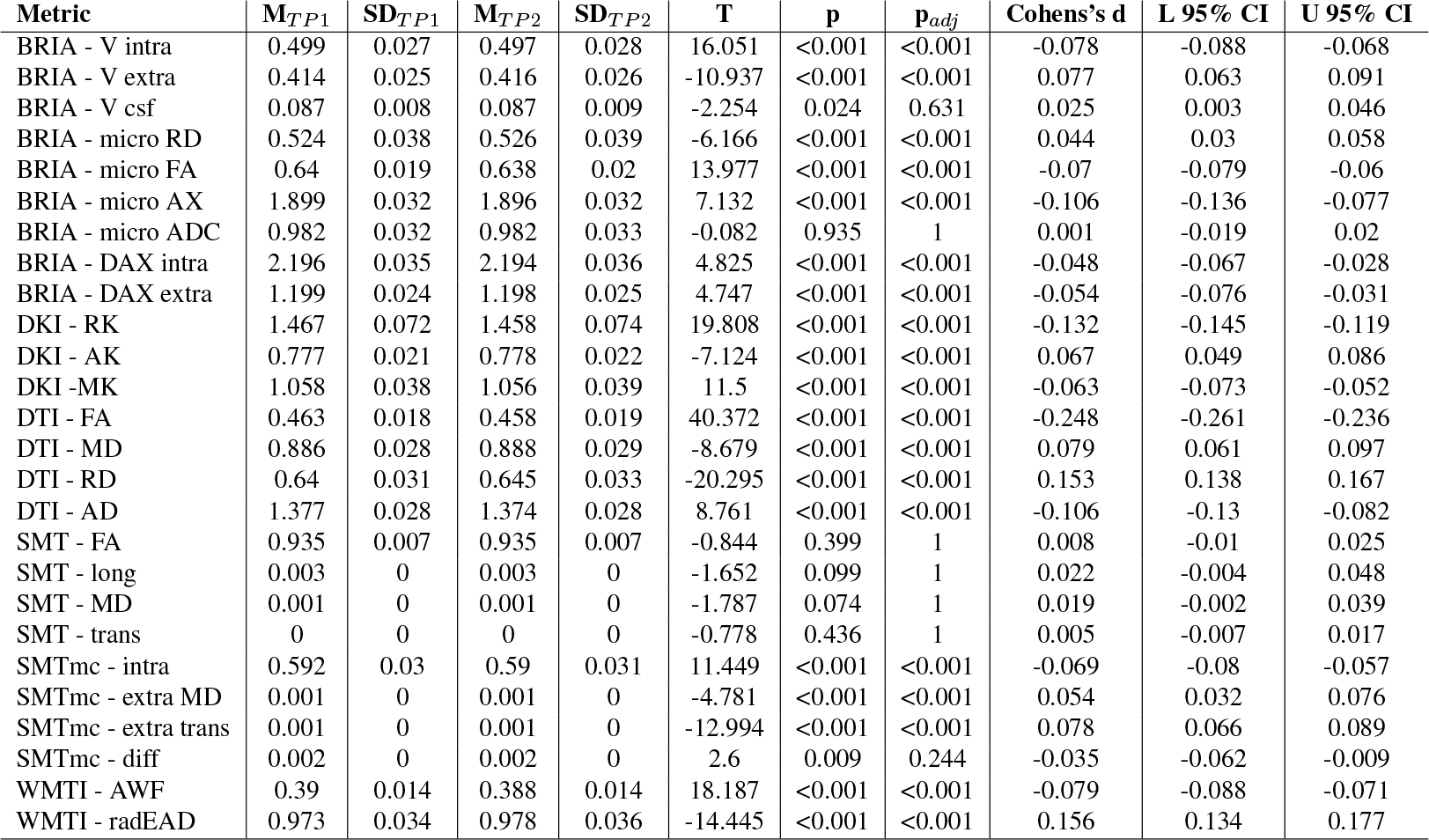

## Supplementary Note 5 Effects of age, time point, sex and the age-sex interaction on global diffusion metrics in LMER

Standardized *β* values *±* Standard Error indicating the fixed effects of (a) age, (b) time point, (c) sex, and (d) sex*×*age on global diffusion metrics. The colour indicates the metric’s corresponding diffusion approach.

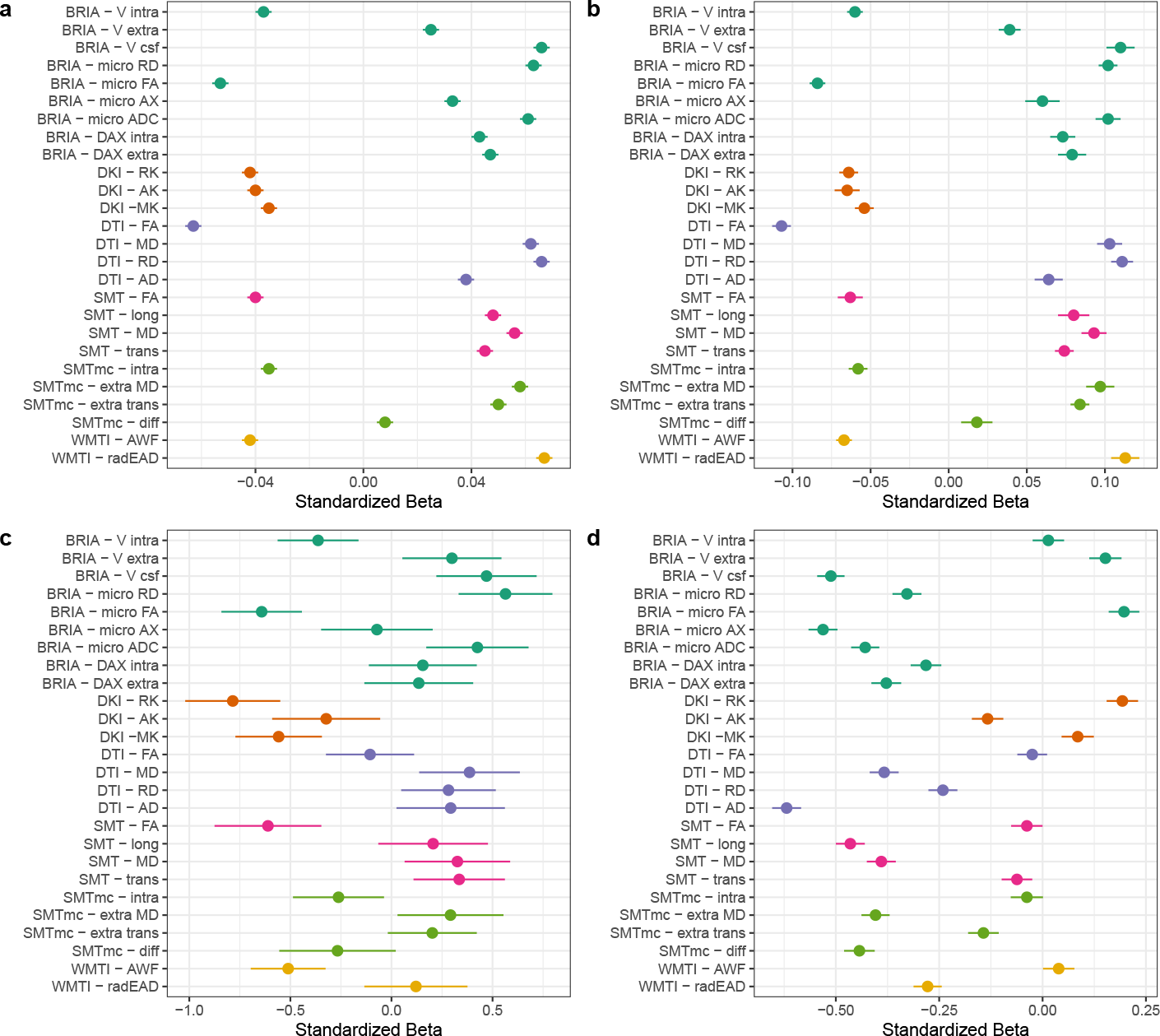

## Supplementary Note 6 Test statistics for the effect of age on global diffusion metrics in LMER

*β*_*std*_ refers to the standardized slopes and *SE* to the standard error.

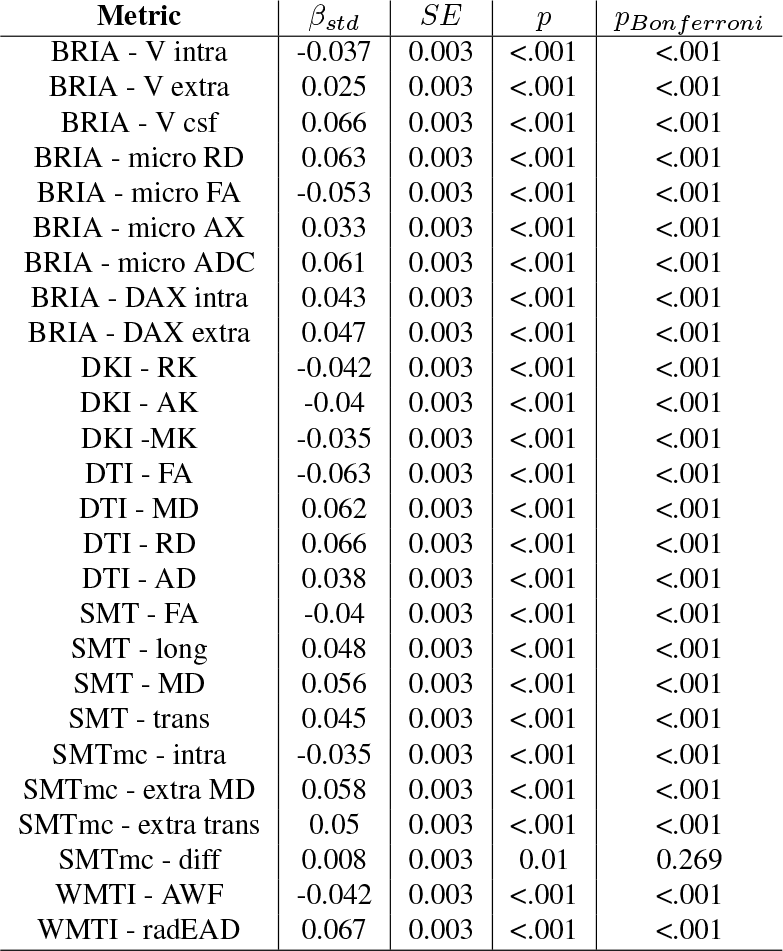

## Supplementary Note 7 The effect of time point on global diffusion metrics in mixed linear models

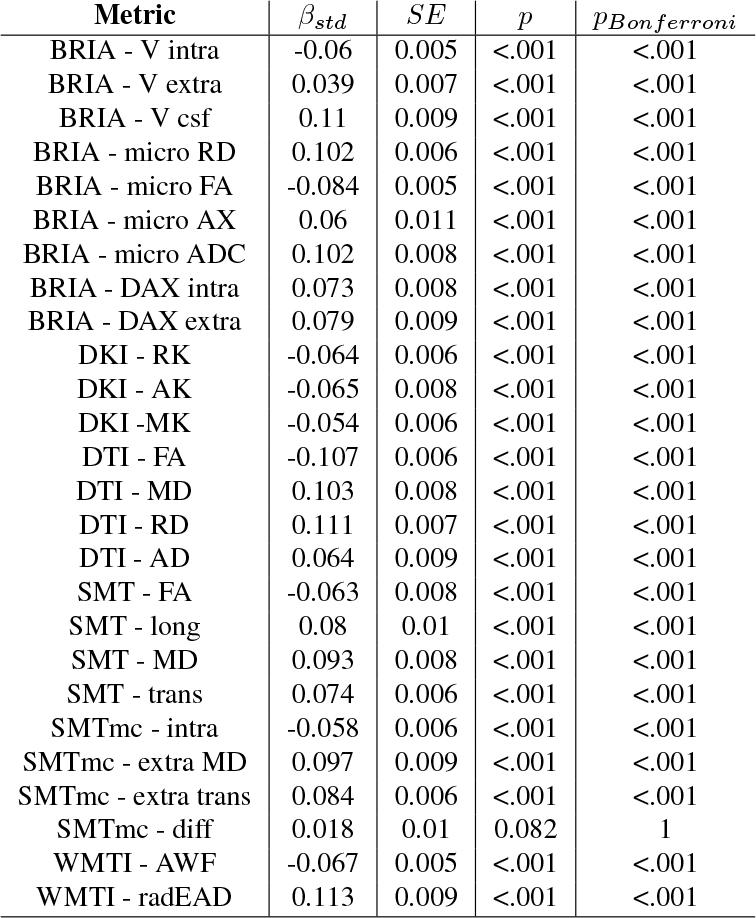

## Supplementary Note 8 Mixed linear model diagnostics

The following table shows the model diagnostics when predicting diffusion metrics from age, sex, age *×* sex, and time point, while accounting for the participants nested in scanner site as random intercept.

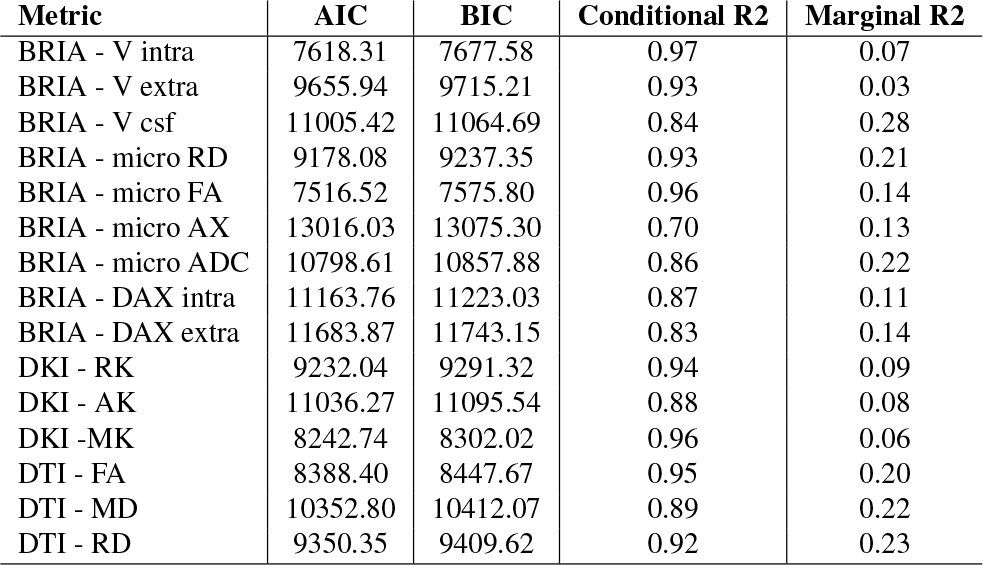

## Supplementary Note 9 The effect of sex on global diffusion metrics in LMER

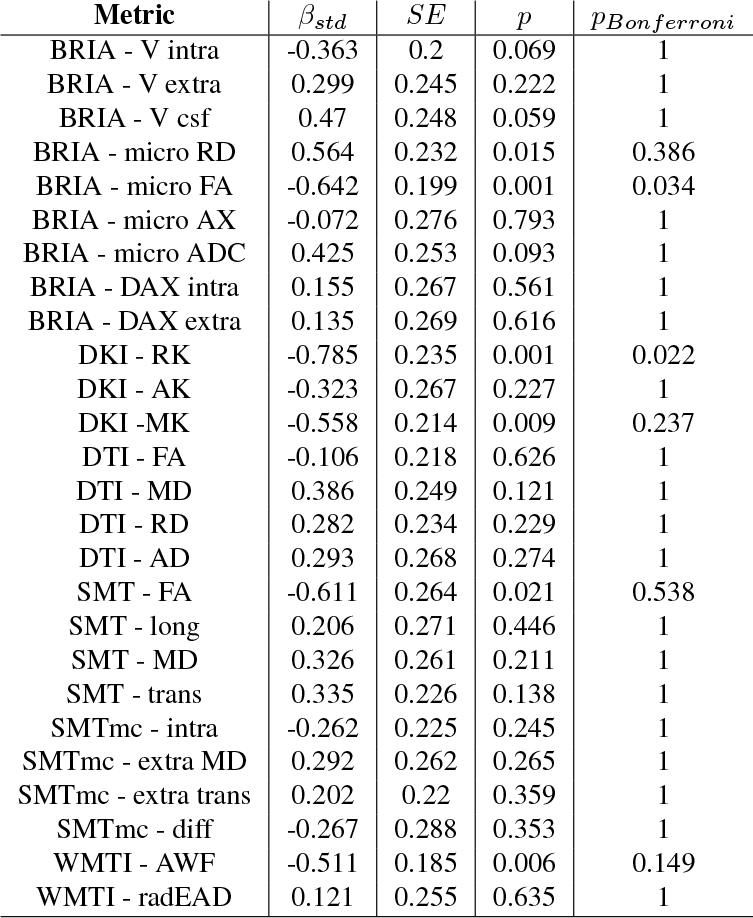

## Supplementary Note 10 The effect of the sex-age interaction on global diffusion metrics in LMER

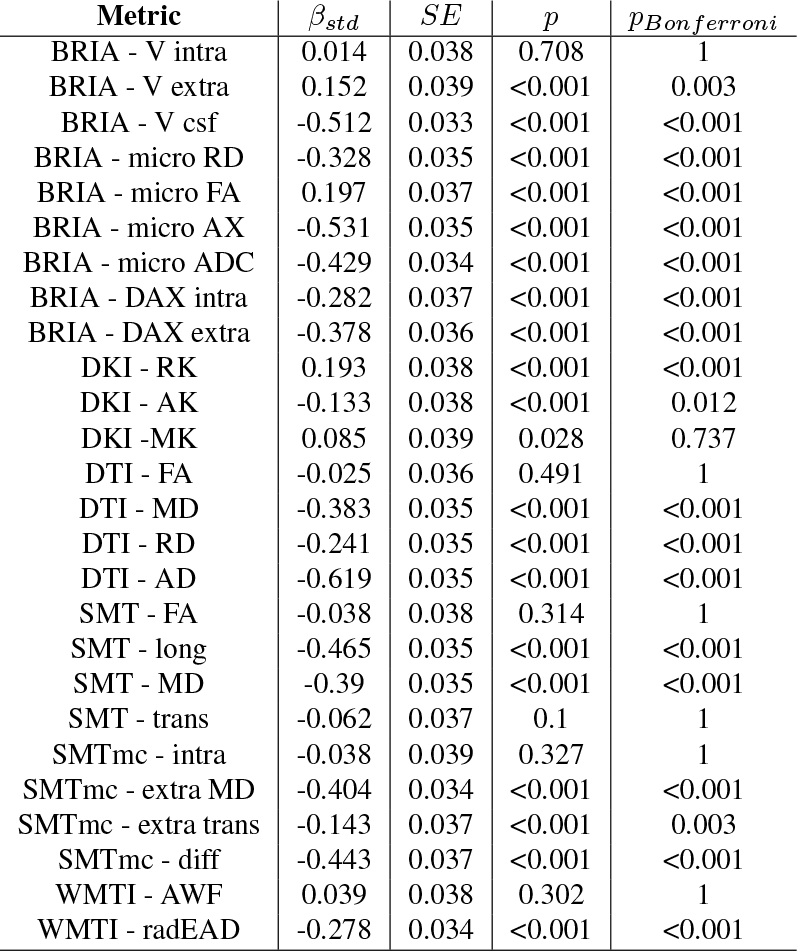

## Supplementary Note 11 The effect of the sex on regional WMM metrics

Increases refer to higher WMM values for males compared to females and decrease to lower WMM values, respectively.

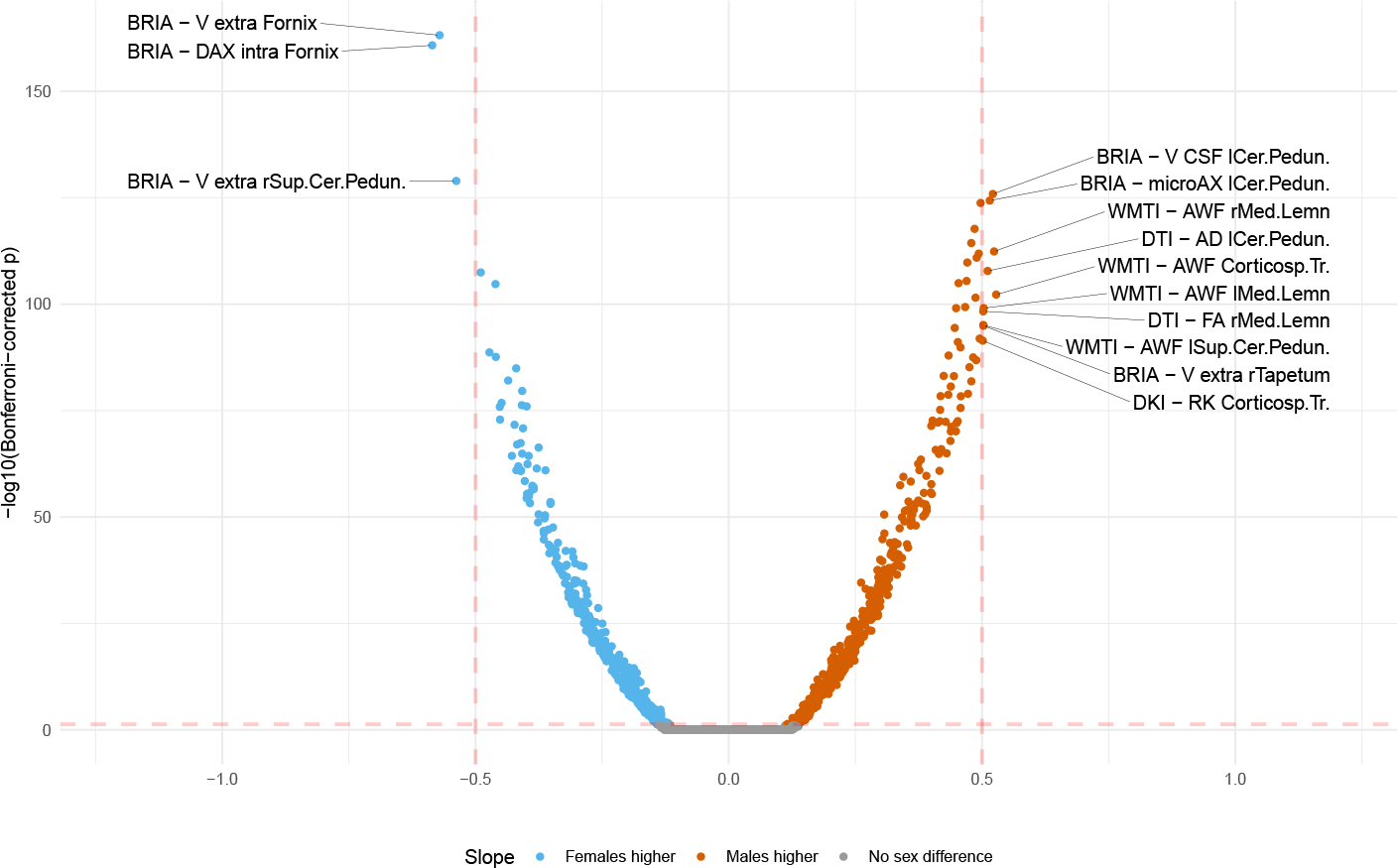

## Supplementary Note 12 The effect of the sex-age interaction on regional WMM metrics

Increases refer for higher WMM values for males at higher ages compared to females and decrease to lower WMM values, respectively.

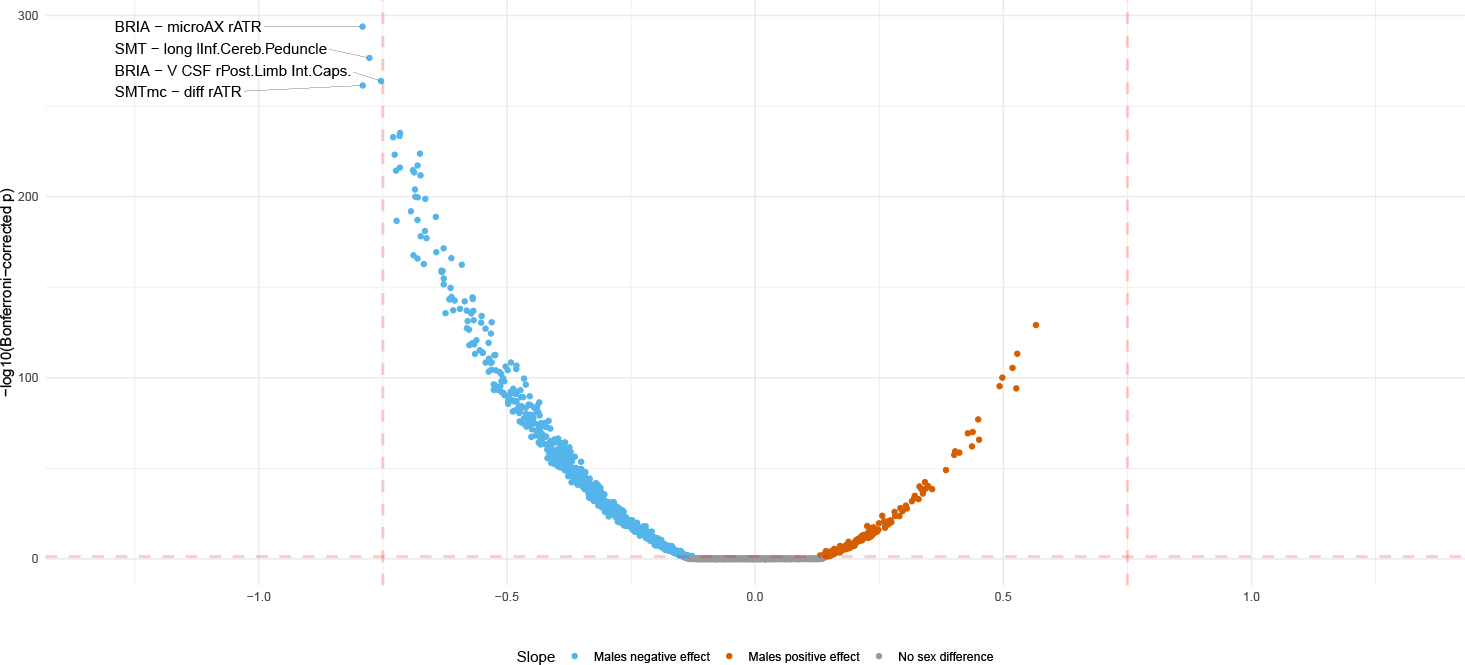

## Supplementary Note 13 Distribution of the effect of age on regional WMM metrics

Estimating average slopes from all *β*-values results in the following meta-statistics for negative *β*-values (left side of 0): Mean = -0.21±0.13, Median = -0.23± 0.12. For positive *β*-values (right side of 0) the following averages were estimated: Mean = 0.25±0.13, Median = 0.27± 0.14.

Estimating average slopes from significant effects only (right panel) results in similar meta-statistics for negative *β*-values (left side of 0): Mean = -0.21±0.13, Median = -0.24± 0.12, as well as for positive *β*-values (right side of 0): Mean = 0.25±0.13, Median = 0.27± 0.15.

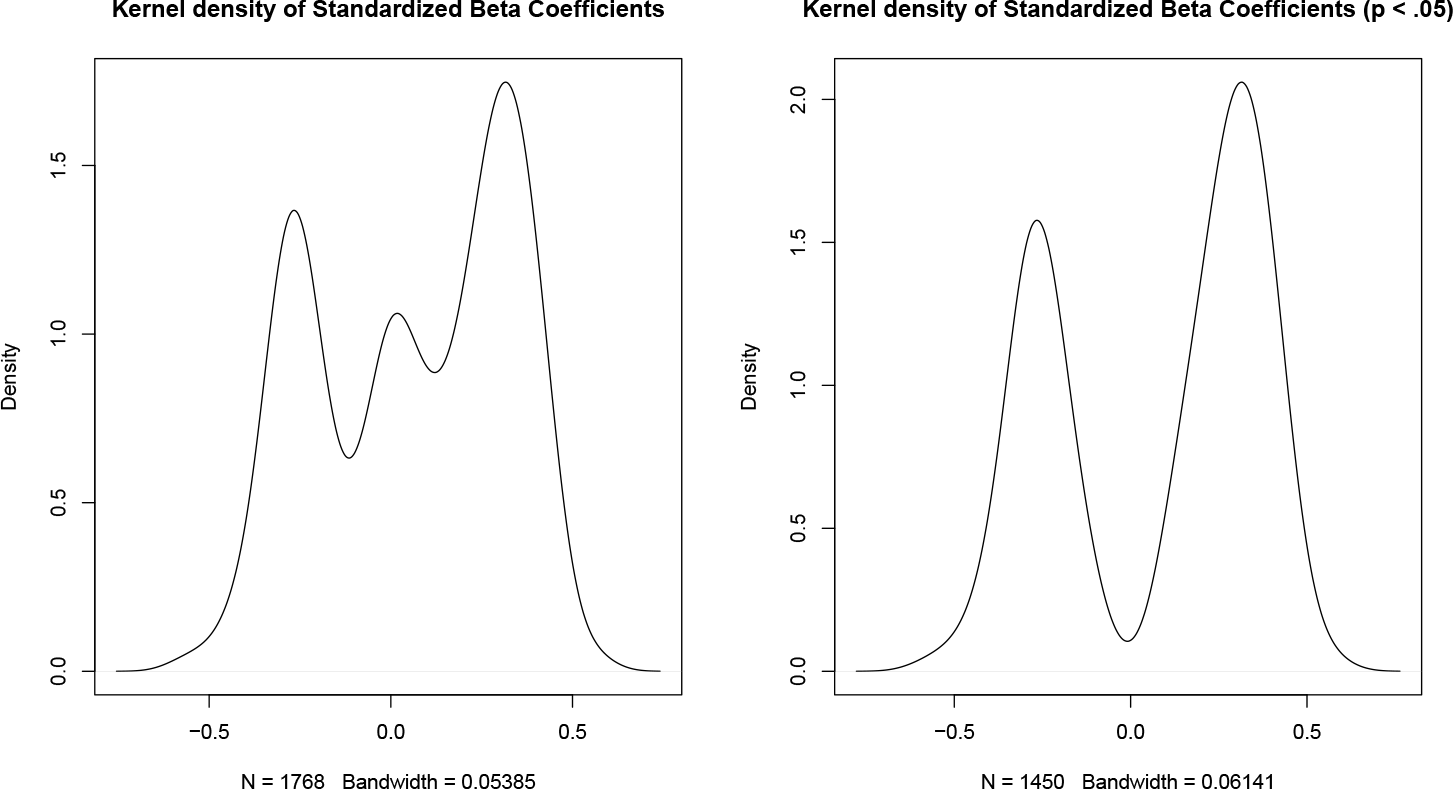

## Supplementary Note 14 Adjusted cross-sectional associations between global WMM and age at time point 1

Age-WMM relationships were adjusted for sex, the sex-age interaction, and site with the fit indicated for each time point.

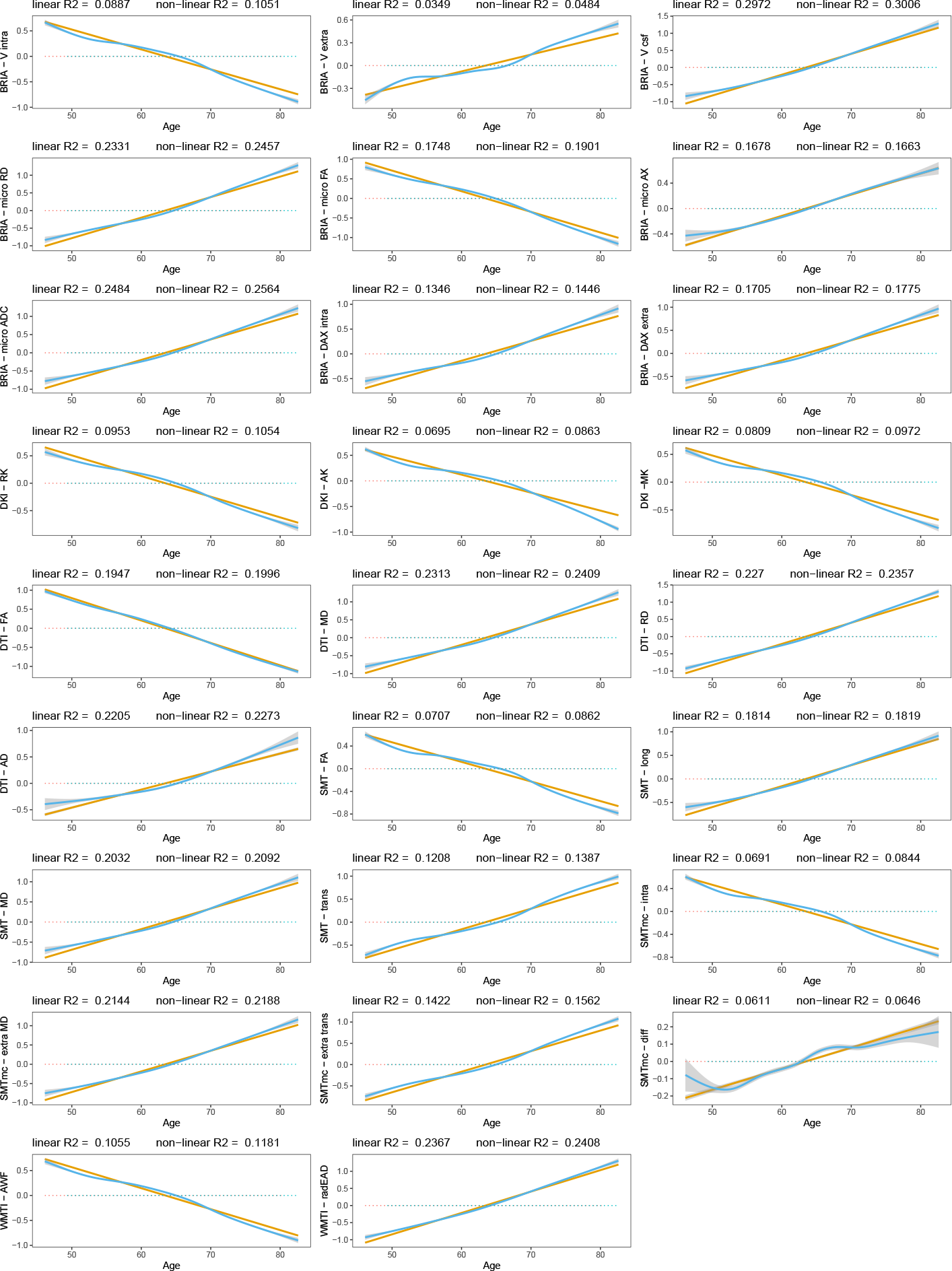

## Supplementary Note 15 Adjusted cross-sectional associations between global WMM and age at time point 1

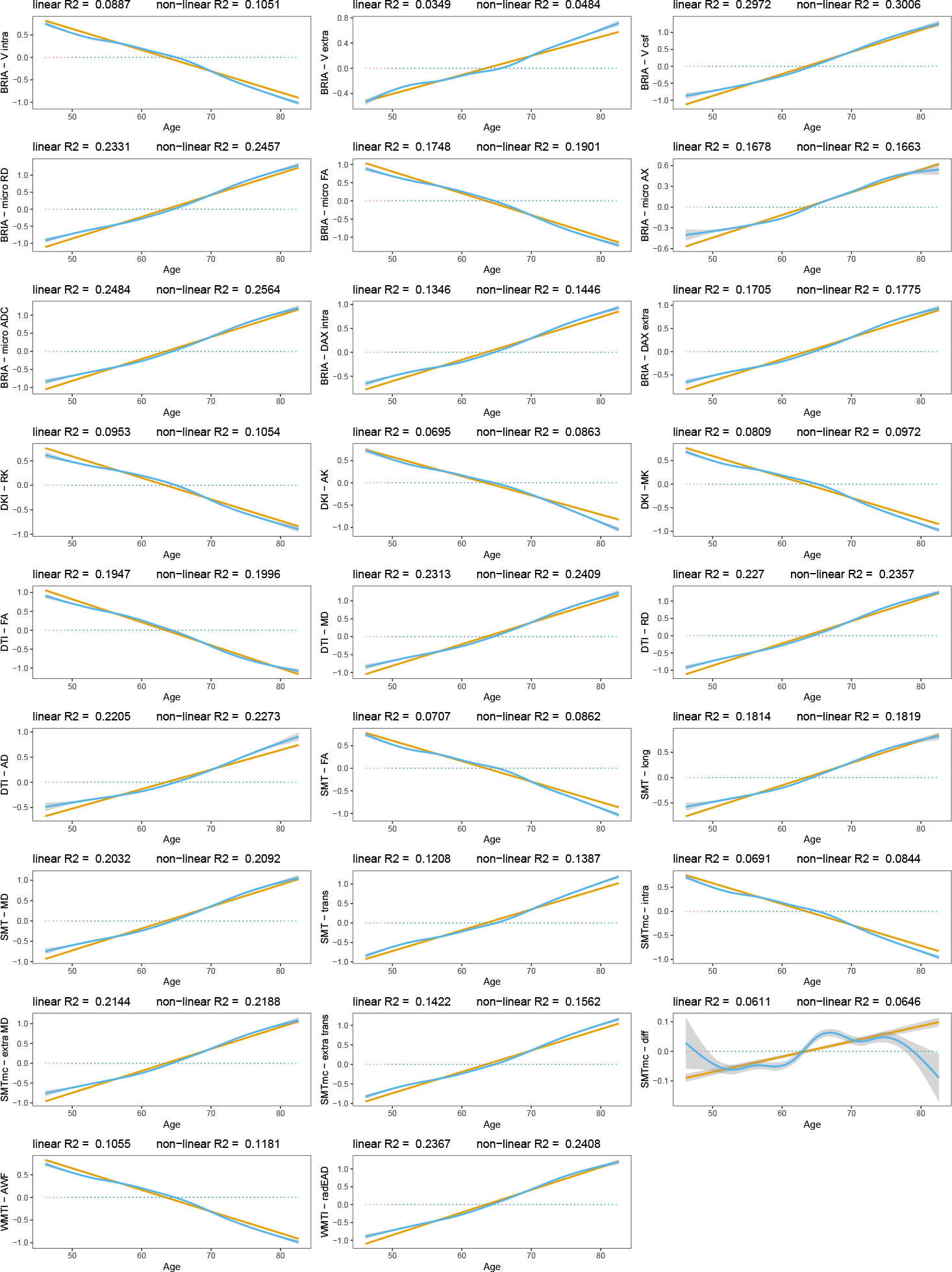

## Supplementary Note 16 Cognitive test differences between time points

Pair matches refers to incorrectly solved pair matches (executed in 3 rounds). Digits refers to the maximum number of digits remembered; intelligence to fluid intelligence; memory to prospective memory; health to a rating of the own overall health; matrix RTs refer to matrix puzzle completion times; matrix correct to the number of correctly solved matrix puzzles; matrix viewed to the number of viewed matrix puzzles; tower correct to the number of correctly solved tower puzzles; and Sym/Dig matches to the number of correct symbol-digit matches.

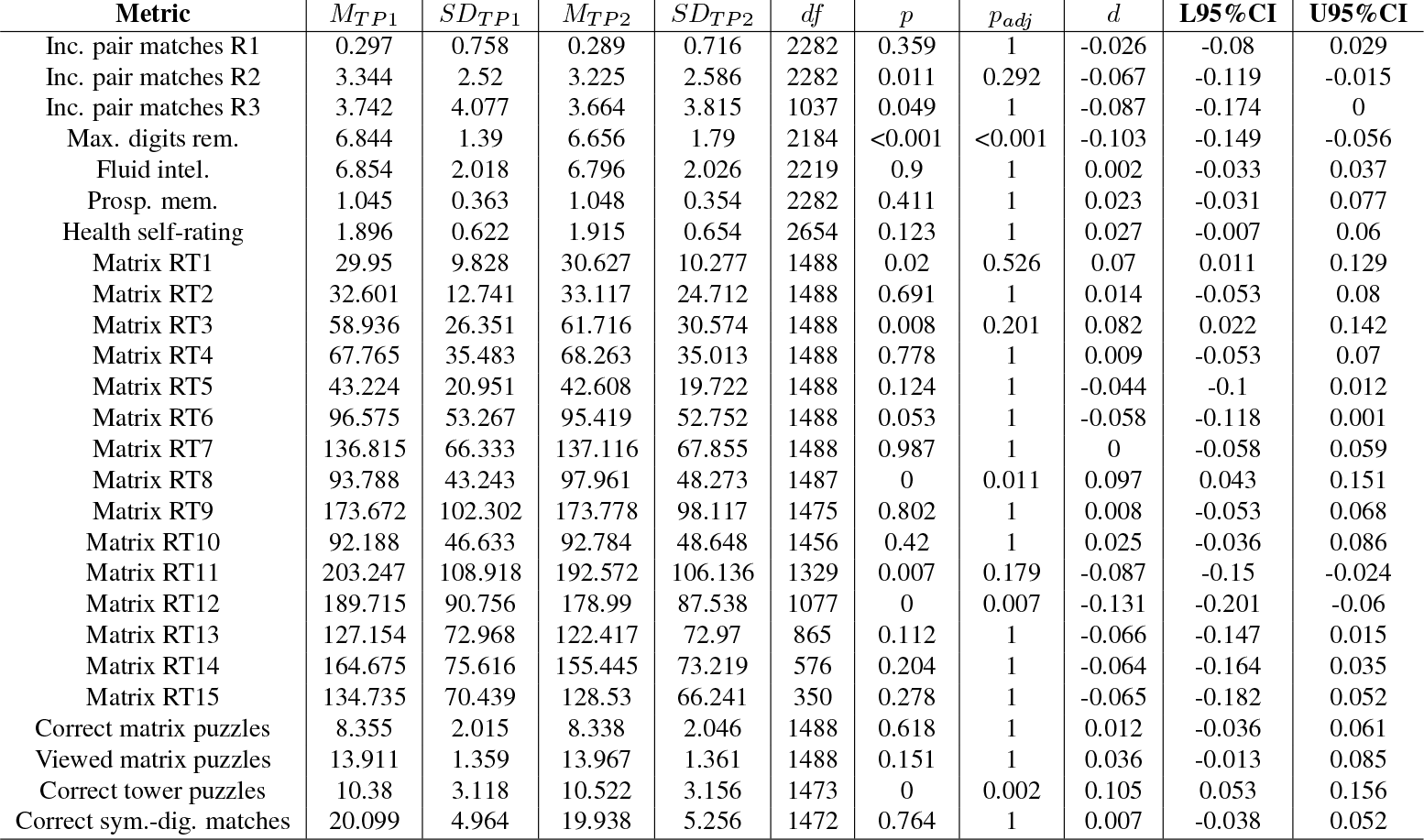

## Supplementary Note 17 Distribution of the relationship between age and the absolute annual rate of white matter microstructure change

The Figure shows the distribution of adjusted |ARoC|-age associations across brain regions 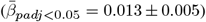. Mean associations were smaller when including non-significant associations 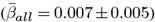.

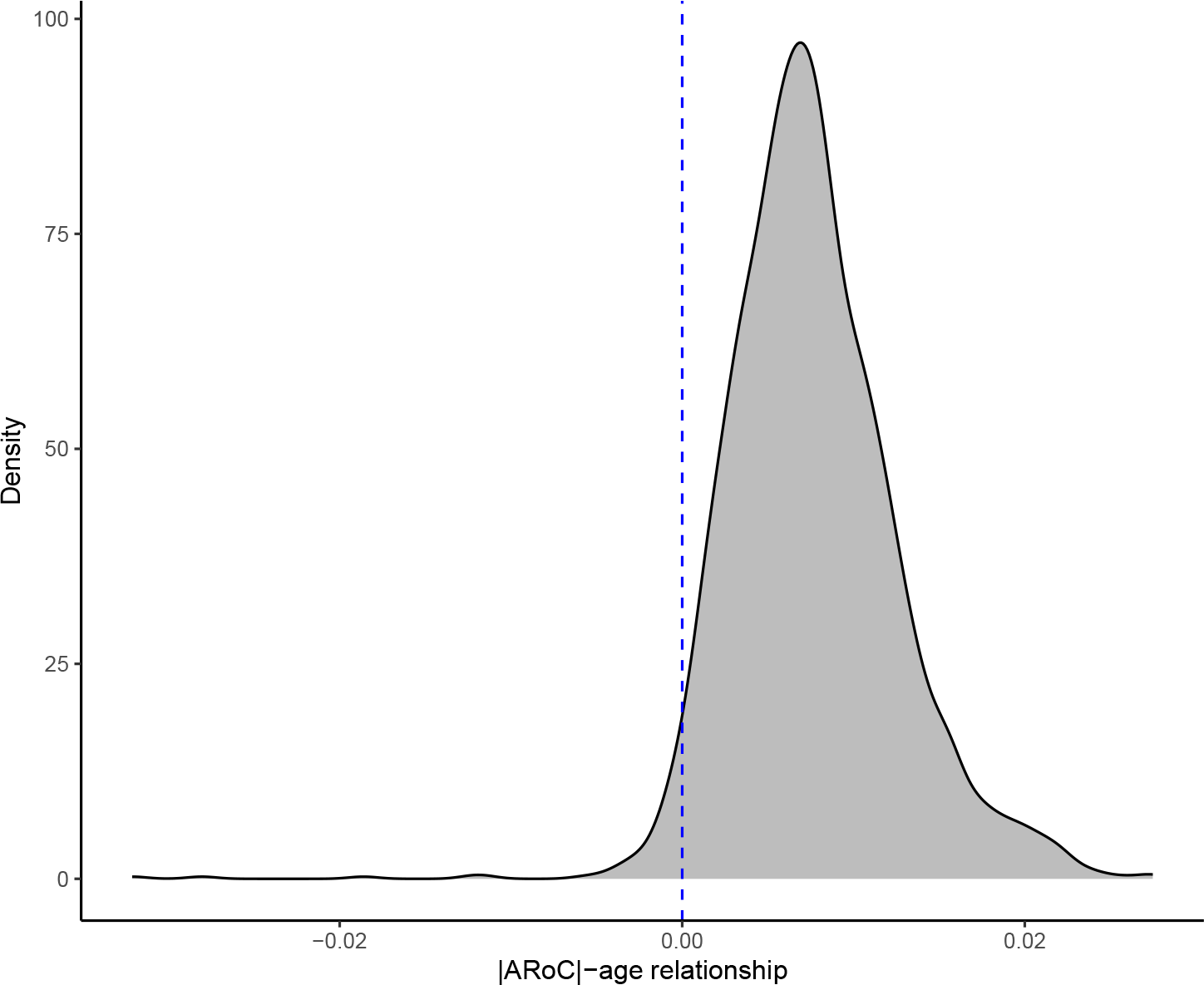

## Supplementary Note 18 Age-stratified annual rate of WMM change group-comparisons

**Table 2.** Age-stratified annual rate of WMM change group-comparisons.

| Diffusion Metric | Group 1 | Group 2 | Cohen's d | n<sub>1</sub> | n<sub>2</sub> | CI<sub>low</sub> | CI<sub>high</sub> | Magnitude | p<sub>adj</sub> |
| --- | --- | --- | --- | --- | --- | --- | --- | --- | --- |
| BRIA - v intra | 46-55 | 55-65 | 0.65 | 525 | 1157 | 0.54 | 0.78 | moderate | <.001 |
| BRIA - v intra | 46-55 | 65-75 | 1.31 | 525 | 905 | 1.18 | 1.43 | large | <.001 |
| BRIA - v intra | 46-55 | 75-81 | 1.78 | 525 | 91 | 1.54 | 2.06 | large | <.001 |
| BRIA - v intra | 55-65 | 65-75 | 0.80 | 1157 | 905 | 0.71 | 0.9 | large | <.001 |
| BRIA - v intra | 55-65 | 75-81 | 1.41 | 1157 | 91 | 1.14 | 1.73 | large | <.001 |
| BRIA - v intra | 65-75 | 75-81 | 0.78 | 905 | 91 | 0.52 | 1.05 | moderate | <.001 |
| BRIA - v extra | 46-55 | 55-65 | -0.73 | 525 | 1157 | -0.86 | -0.6 | moderate | <.001 |
| BRIA - v extra | 46-55 | 65-75 | -1.44 | 525 | 905 | -1.57 | -1.31 | large | <.001 |
| BRIA - v extra | 46-55 | 75-81 | -1.88 | 525 | 91 | -2.2 | -1.62 | large | <.001 |
| BRIA - v extra | 55-65 | 65-75 | -0.94 | 1157 | 905 | -1.04 | -0.85 | large | <.001 |
| BRIA - v extra | 55-65 | 75-81 | -1.53 | 1157 | 91 | -1.88 | -1.26 | large | <.001 |
| BRIA - v extra | 65-75 | 75-81 | -0.83 | 905 | 91 | -1.12 | -0.58 | large | <.001 |
| BRIA - v CSF | 46-55 | 55-65 | -0.30 | 525 | 1157 | -0.41 | -0.19 | small | <.001 |
| BRIA - v CSF | 46-55 | 65-75 | -0.51 | 525 | 905 | -0.62 | -0.39 | moderate | <.001 |
| BRIA - v CSF | 46-55 | 75-81 | -0.71 | 525 | 91 | -0.92 | -0.5 | moderate | <.001 |
| BRIA - v CSF | 55-65 | 65-75 | -0.20 | 1157 | 905 | -0.29 | -0.12 | small | <.001 |
| BRIA - v CSF | 55-65 | 75-81 | -0.41 | 1157 | 91 | -0.61 | -0.2 | small | 0.001 |
| BRIA - v CSF | 65-75 | 75-81 | -0.21 | 905 | 91 | -0.43 | 0.01 | small | 0.064 |
| BRIA - micro RD | 46-55 | 55-65 | -0.45 | 525 | 1157 | -0.55 | -0.35 | small | <.001 |
| BRIA - micro RD | 46-55 | 65-75 | -0.85 | 525 | 905 | -0.98 | -0.74 | large | <.001 |
| BRIA - micro RD | 46-55 | 75-81 | -1.25 | 525 | 91 | -1.5 | -1.02 | large | <.001 |
| BRIA - micro RD | 55-65 | 65-75 | -0.42 | 1157 | 905 | -0.51 | -0.33 | small | <.001 |
| BRIA - micro RD | 55-65 | 75-81 | -0.87 | 1157 | 91 | -1.13 | -0.66 | large | <.001 |
| BRIA - micro RD | 65-75 | 75-81 | -0.48 | 905 | 91 | -0.72 | -0.26 | small | <.001 |
| BRIA - micro FA | 46-55 | 55-65 | 0.56 | 525 | 1157 | 0.45 | 0.67 | moderate | <.001 |
| BRIA - micro FA | 46-55 | 65-75 | 1.11 | 525 | 905 | 0.99 | 1.24 | large | <.001 |
| BRIA - micro FA | 46-55 | 75-81 | 1.58 | 525 | 91 | 1.35 | 1.84 | large | <.001 |
| BRIA - micro FA | 55-65 | 65-75 | 0.63 | 1157 | 905 | 0.55 | 0.72 | moderate | <.001 |
| BRIA - micro FA | 55-65 | 75-81 | 1.21 | 1157 | 91 | 0.96 | 1.49 | large | <.001 |
| BRIA - micro FA | 65-75 | 75-81 | 0.68 | 905 | 91 | 0.44 | 0.92 | moderate | <.001 |
| BRIA - micro AX | 46-55 | 55-65 | -0.06 | 525 | 1157 | -0.16 | 0.04 | negligible | 0.478 |
| BRIA - micro AX | 46-55 | 65-75 | 0.04 | 525 | 905 | -0.07 | 0.14 | negligible | 0.519 |
| BRIA - micro AX | 46-55 | 75-81 | 0.20 | 525 | 91 | -0.05 | 0.43 | small | 0.332 |
| BRIA - micro AX | 55-65 | 65-75 | 0.09 | 1157 | 905 | 0.01 | 0.19 | negligible | 0.170 |
| BRIA - micro AX | 55-65 | 75-81 | 0.26 | 1157 | 91 | 0.04 | 0.48 | small | 0.121 |
| BRIA - micro AX | 65-75 | 75-81 | 0.16 | 905 | 91 | -0.04 | 0.39 | negligible | 0.426 |
| BRIA - micro ADC | 46-55 | 55-65 | -0.32 | 525 | 1157 | -0.44 | -0.22 | small | <.001 |

Table 2: Age-stratified annual rate of WMM change group-comparisons (Continued)
|  |  |  |  |  |  |  |  |  |  |
| --- | --- | --- | --- | --- | --- | --- | --- | --- | --- |
| BRIA – micro ADC | 46-55 | 65-75 | -0.55 | 525 | 905 | -0.66 | -0.43 | moderate | <.001 |
| BRIA – micro ADC | 46-55 | 75-81 | -0.80 | 525 | 91 | -1.03 | -0.58 | moderate | <.001 |
| BRIA – micro ADC | 55-65 | 65-75 | -0.23 | 1157 | 905 | -0.32 | -0.14 | small | <.001 |
| BRIA – micro ADC | 55-65 | 75-81 | -0.48 | 1157 | 91 | -0.7 | -0.28 | small | <.001 |
| BRIA – micro ADC | 65-75 | 75-81 | -0.26 | 905 | 91 | -0.48 | -0.04 | small | 0.023 |
| BRIA – DAX intra | 46-55 | 55-65 | -0.30 | 525 | 1157 | -0.41 | -0.2 | small | <.001 |
| BRIA – DAX intra | 46-55 | 65-75 | -0.51 | 525 | 905 | -0.62 | -0.4 | moderate | <.001 |
| BRIA – DAX intra | 46-55 | 75-81 | -0.74 | 525 | 91 | -0.96 | -0.52 | moderate | <.001 |
| BRIA – DAX intra | 55-65 | 65-75 | -0.20 | 1157 | 905 | -0.29 | -0.11 | small | <.001 |
| BRIA – DAX intra | 55-65 | 75-81 | -0.44 | 1157 | 91 | -0.67 | -0.23 | small | <.001 |
| BRIA – DAX intra | 65-75 | 75-81 | -0.24 | 905 | 91 | -0.47 | -0.02 | small | 0.033 |
| BRIA – DAX extra | 46-55 | 55-65 | -0.26 | 525 | 1157 | -0.36 | -0.15 | small | <.001 |
| BRIA – DAX extra | 46-55 | 65-75 | -0.40 | 525 | 905 | -0.51 | -0.29 | small | <.001 |
| BRIA – DAX extra | 46-55 | 75-81 | -0.56 | 525 | 91 | -0.78 | -0.33 | moderate | <.001 |
| BRIA – DAX extra | 55-65 | 65-75 | -0.15 | 1157 | 905 | -0.24 | -0.05 | negligible | 0.003 |
| BRIA – DAX extra | 55-65 | 75-81 | -0.31 | 1157 | 91 | -0.53 | -0.09 | small | 0.012 |
| BRIA – DAX extra | 65-75 | 75-81 | -0.16 | 905 | 91 | -0.39 | 0.06 | negligible | 0.145 |
| DKI – RK | 46-55 | 55-65 | 0.59 | 525 | 1157 | 0.48 | 0.7 | moderate | <.001 |
| DKI – RK | 46-55 | 65-75 | 1.34 | 525 | 905 | 1.24 | 1.45 | large | <.001 |
| DKI – RK | 46-55 | 75-81 | 1.90 | 525 | 91 | 1.69 | 2.17 | large | <.001 |
| DKI – RK | 55-65 | 65-75 | 1.05 | 1157 | 905 | 0.98 | 1.14 | large | <.001 |
| DKI – RK | 55-65 | 75-81 | 1.73 | 1157 | 91 | 1.44 | 2.05 | large | <.001 |
| DKI – RK | 65-75 | 75-81 | 0.98 | 905 | 91 | 0.71 | 1.29 | large | <.001 |
| DKI – AK | 46-55 | 55-65 | 0.41 | 525 | 1157 | 0.31 | 0.51 | small | <.001 |
| DKI – AK | 46-55 | 65-75 | 0.75 | 525 | 905 | 0.64 | 0.87 | moderate | <.001 |
| DKI – AK | 46-55 | 75-81 | 1.12 | 525 | 91 | 0.88 | 1.36 | large | <.001 |
| DKI – AK | 55-65 | 65-75 | 0.35 | 1157 | 905 | 0.27 | 0.44 | small | <.001 |
| DKI – AK | 55-65 | 75-81 | 0.76 | 1157 | 91 | 0.54 | 0.99 | moderate | <.001 |
| DKI – AK | 65-75 | 75-81 | 0.44 | 905 | 91 | 0.23 | 0.67 | small | <.001 |
| DKI – MK | 46-55 | 55-65 | 0.61 | 525 | 1157 | 0.5 | 0.72 | moderate | <.001 |
| DKI – MK | 46-55 | 65-75 | 1.22 | 525 | 905 | 1.09 | 1.34 | large | <.001 |
| DKI – MK | 46-55 | 75-81 | 1.69 | 525 | 91 | 1.45 | 1.97 | large | <.001 |
| DKI – MK | 55-65 | 65-75 | 0.72 | 1157 | 905 | 0.64 | 0.81 | moderate | <.001 |
| DKI – MK | 55-65 | 75-81 | 1.32 | 1157 | 91 | 1.07 | 1.6 | large | <.001 |
| DKI – MK | 65-75 | 75-81 | 0.74 | 905 | 91 | 0.51 | 0.99 | moderate | <.001 |
| DTI – FA | 46-55 | 55-65 | 0.23 | 525 | 1157 | 0.11 | 0.34 | small | <.001 |
| DTI – FA | 46-55 | 65-75 | 0.66 | 525 | 905 | 0.56 | 0.77 | moderate | <.001 |
| DTI – FA | 46-55 | 75-81 | 1.00 | 525 | 91 | 0.74 | 1.29 | large | <.001 |
| DTI – FA | 55-65 | 65-75 | 0.46 | 1157 | 905 | 0.36 | 0.55 | small | <.001 |
| DTI – FA | 55-65 | 75-81 | 0.82 | 1157 | 91 | 0.58 | 1.1 | large | <.001 |
| DTI – FA | 65-75 | 75-81 | 0.40 | 905 | 91 | 0.17 | 0.66 | small | 0.001 |

Table 2: Age-stratified annual rate of WMM change group-comparisons (Continued)
|  |  |  |  |  |  |  |  |  |  |
| --- | --- | --- | --- | --- | --- | --- | --- | --- | --- |
| DTI – MD | 46-55 | 55-65 | -0.43 | 525 | 1157 | -0.54 | -0.33 | small | <.001 |
| DTI – MD | 46-55 | 65-75 | -0.79 | 525 | 905 | -0.91 | -0.67 | moderate | <.001 |
| DTI – MD | 46-55 | 75-81 | -1.15 | 525 | 91 | -1.41 | -0.94 | large | <.001 |
| DTI – MD | 55-65 | 65-75 | -0.38 | 1157 | 905 | -0.46 | -0.29 | small | <.001 |
| DTI – MD | 55-65 | 75-81 | -0.77 | 1157 | 91 | -1 | -0.56 | moderate | <.001 |
| DTI – MD | 65-75 | 75-81 | -0.40 | 905 | 91 | -0.62 | -0.17 | small | 0.001 |
| DTI – RD | 46-55 | 55-65 | -0.74 | 525 | 1157 | -0.85 | -0.63 | moderate | <.001 |
| DTI – RD | 46-55 | 65-75 | -1.47 | 525 | 905 | -1.59 | -1.36 | large | <.001 |
| DTI – RD | 46-55 | 75-81 | -1.92 | 525 | 91 | -2.27 | -1.63 | large | <.001 |
| DTI – RD | 55-65 | 65-75 | -0.93 | 1157 | 905 | -1.04 | -0.85 | large | <.001 |
| DTI – RD | 55-65 | 75-81 | -1.53 | 1157 | 91 | -1.88 | -1.24 | large | <.001 |
| DTI – RD | 65-75 | 75-81 | -0.76 | 905 | 91 | -1.04 | -0.52 | moderate | <.001 |
| DTI – AD | 46-55 | 55-65 | -0.22 | 525 | 1157 | -0.33 | -0.11 | small | <.001 |
| DTI – AD | 46-55 | 65-75 | -0.32 | 525 | 905 | -0.43 | -0.2 | small | <.001 |
| DTI – AD | 46-55 | 75-81 | -0.43 | 525 | 91 | -0.67 | -0.2 | small | 0.001 |
| DTI – AD | 55-65 | 65-75 | -0.09 | 1157 | 905 | -0.19 | -0.0066 | negligible | 0.109 |
| DTI – AD | 55-65 | 75-81 | -0.21 | 1157 | 91 | -0.42 | 0.007 | small | 0.114 |
| DTI – AD | 65-75 | 75-81 | -0.12 | 905 | 91 | -0.35 | 0.08 | negligible | 0.289 |
| SMT – FA | 46-55 | 55-65 | 0.53 | 525 | 1157 | 0.43 | 0.64 | moderate | <.001 |
| SMT – FA | 46-55 | 65-75 | 1.06 | 525 | 905 | 0.94 | 1.19 | large | <.001 |
| SMT – FA | 46-55 | 75-81 | 1.51 | 525 | 91 | 1.28 | 1.77 | large | <.001 |
| SMT – FA | 55-65 | 65-75 | 0.59 | 1157 | 905 | 0.5 | 0.68 | moderate | <.001 |
| SMT – FA | 55-65 | 75-81 | 1.15 | 1157 | 91 | 0.93 | 1.41 | large | <.001 |
| SMT – FA | 65-75 | 75-81 | 0.66 | 905 | 91 | 0.43 | 0.91 | moderate | <.001 |
| SMT – long | 46-55 | 55-65 | -0.15 | 525 | 1157 | -0.26 | -0.04 | negligible | 0.021 |
| SMT – long | 46-55 | 65-75 | -0.16 | 525 | 905 | -0.27 | -0.05 | negligible | 0.018 |
| SMT – long | 46-55 | 75-81 | -0.11 | 525 | 91 | -0.35 | 0.1 | negligible | 1.000 |
| SMT – long | 55-65 | 65-75 | -0.01 | 1157 | 905 | -0.11 | 0.07 | negligible | 1.000 |
| SMT – long | 55-65 | 75-81 | 0.04 | 1157 | 91 | -0.19 | 0.25 | negligible | 1.000 |
| SMT – long | 65-75 | 75-81 | 0.05 | 905 | 91 | -0.18 | 0.3 | negligible | 1.000 |
| SMT – MD | 46-55 | 55-65 | -0.28 | 525 | 1157 | -0.39 | -0.18 | small | <.001 |
| SMT – MD | 46-55 | 65-75 | -0.46 | 525 | 905 | -0.57 | -0.35 | small | <.001 |
| SMT – MD | 46-55 | 75-81 | -0.64 | 525 | 91 | -0.87 | -0.42 | moderate | <.001 |
| SMT – MD | 55-65 | 65-75 | -0.18 | 1157 | 905 | -0.26 | -0.09 | negligible | <.001 |
| SMT – MD | 55-65 | 75-81 | -0.36 | 1157 | 91 | -0.58 | -0.15 | small | 0.003 |
| SMT – MD | 65-75 | 75-81 | -0.19 | 905 | 91 | -0.42 | 0.02 | negligible | 0.096 |
| SMT – trans | 46-55 | 55-65 | -0.49 | 525 | 1157 | -0.6 | -0.38 | small | <.001 |
| SMT – trans | 46-55 | 65-75 | -0.93 | 525 | 905 | -1.06 | -0.82 | large | <.001 |
| SMT – trans | 46-55 | 75-81 | -1.36 | 525 | 91 | -1.61 | -1.14 | large | <.001 |
| SMT – trans | 55-65 | 65-75 | -0.47 | 1157 | 905 | -0.56 | -0.39 | small | <.001 |
| SMT – trans | 55-65 | 75-81 | -0.97 | 1157 | 91 | -1.23 | -0.74 | large | <.001 |

Table 2: Age-stratified annual rate of WMM change group-comparisons (Continued)
|  |  |  |  |  |  |  |  |  |  |
| --- | --- | --- | --- | --- | --- | --- | --- | --- | --- |
| SMT – trans | 65-75 | 75-81 | -0.55 | 905 | 91 | -0.8 | -0.32 | moderate | <.001 |
| SMTmc – intra | 46-55 | 55-65 | 0.70 | 525 | 1157 | 0.6 | 0.81 | moderate | <.001 |
| SMTmc – intra | 46-55 | 65-75 | 1.36 | 525 | 905 | 1.23 | 1.49 | large | <.001 |
| SMTmc – intra | 46-55 | 75-81 | 1.82 | 525 | 91 | 1.56 | 2.14 | large | <.001 |
| SMTmc – intra | 55-65 | 65-75 | 0.82 | 1157 | 905 | 0.73 | 0.9 | large | <.001 |
| SMTmc – intra | 55-65 | 75-81 | 1.42 | 1157 | 91 | 1.15 | 1.75 | large | <.001 |
| SMTmc – intra | 65-75 | 75-81 | 0.76 | 905 | 91 | 0.5 | 1.03 | moderate | <.001 |
| SMTmc – extra MD | 46-55 | 55-65 | -0.32 | 525 | 1157 | -0.43 | -0.21 | small | <.001 |
| SMTmc – extra MD | 46-55 | 65-75 | -0.55 | 525 | 905 | -0.67 | -0.44 | moderate | <.001 |
| SMTmc – extra MD | 46-55 | 75-81 | -0.77 | 525 | 91 | -1 | -0.55 | moderate | <.001 |
| SMTmc – extra MD | 55-65 | 65-75 | -0.23 | 1157 | 905 | -0.32 | -0.14 | small | <.001 |
| SMTmc – extra MD | 55-65 | 75-81 | -0.46 | 1157 | 91 | -0.67 | -0.24 | small | <.001 |
| SMTmc – extra MD | 65-75 | 75-81 | -0.23 | 905 | 91 | -0.45 | -0.01 | small | 0.041 |
| SMTmc – extra trans | 46-55 | 55-65 | -0.61 | 525 | 1157 | -0.72 | -0.49 | moderate | <.001 |
| SMTmc – extra trans | 46-55 | 65-75 | -1.16 | 525 | 905 | -1.28 | -1.05 | large | <.001 |
| SMTmc – extra trans | 46-55 | 75-81 | -1.63 | 525 | 91 | -1.92 | -1.36 | large | <.001 |
| SMTmc – extra trans | 55-65 | 65-75 | -0.64 | 1157 | 905 | -0.73 | -0.55 | moderate | <.001 |
| SMTmc – extra trans | 55-65 | 75-81 | -1.20 | 1157 | 91 | -1.49 | -0.95 | large | <.001 |
| SMTmc – extra trans | 65-75 | 75-81 | -0.64 | 905 | 91 | -0.9 | -0.4 | moderate | <.001 |
| SMTmc – diff | 46-55 | 55-65 | 0.61 | 525 | 1157 | 0.51 | 0.72 | moderate | <.001 |
| SMTmc – diff | 46-55 | 65-75 | 1.23 | 525 | 905 | 1.12 | 1.34 | large | <.001 |
| SMTmc – diff | 46-55 | 75-81 | 1.73 | 525 | 91 | 1.44 | 2.06 | large | <.001 |
| SMTmc – diff | 55-65 | 65-75 | 0.78 | 1157 | 905 | 0.68 | 0.87 | moderate | <.001 |
| SMTmc – diff | 55-65 | 75-81 | 1.40 | 1157 | 91 | 1.1 | 1.79 | large | <.001 |
| SMTmc – diff | 65-75 | 75-81 | 0.73 | 905 | 91 | 0.47 | 1.01 | moderate | <.001 |
| WMTI – AWF | 46-55 | 55-65 | 0.55 | 525 | 1157 | 0.43 | 0.66 | moderate | <.001 |
| WMTI – AWF | 46-55 | 65-75 | 1.11 | 525 | 905 | 0.99 | 1.24 | large | <.001 |
| WMTI – AWF | 46-55 | 75-81 | 1.58 | 525 | 91 | 1.38 | 1.84 | large | <.001 |
| WMTI – AWF | 55-65 | 65-75 | 0.65 | 1157 | 905 | 0.56 | 0.74 | moderate | <.001 |
| WMTI – AWF | 55-65 | 75-81 | 1.24 | 1157 | 91 | 1.02 | 1.49 | large | <.001 |
| WMTI – AWF | 65-75 | 75-81 | 0.72 | 905 | 91 | 0.51 | 0.97 | moderate | <.001 |
| WMTI – radEAD | 46-55 | 55-65 | -0.58 | 525 | 1157 | -0.7 | -0.47 | moderate | <.001 |
| WMTI – radEAD | 46-55 | 65-75 | -1.16 | 525 | 905 | -1.28 | -1.04 | large | <.001 |
| WMTI – radEAD | 46-55 | 75-81 | -1.52 | 525 | 91 | -1.79 | -1.27 | large | <.001 |
| WMTI – radEAD | 55-65 | 65-75 | -0.64 | 1157 | 905 | -0.73 | -0.56 | moderate | <.001 |
| WMTI – radEAD | 55-65 | 75-81 | -1.09 | 1157 | 91 | -1.38 | -0.87 | large | <.001 |
| WMTI – radEAD | 65-75 | 75-81 | -0.52 | 905 | 91 | -0.77 | -0.28 | moderate | <.001 |

## Supplementary Note 19 Adjusted annual rate of WMM change throughout ages

WMM are standardised for comparability without mean centering for a better understanding of onset values. Non-linear curves were fitted, using splines of generalized additive models, and Pearson’s correlation coefficients were estimated (top left in each plot). We present uncorrected *p*-values, which were significant at the Bonferroni-corrected *α* < 0.05/26 = 0.0019. Additionally, we estimated associations between |ARoC| and age (to meaningfully estimate meta-statistics), showing a higher |ARoC| at higher ages 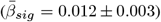, with a slightly lower average slopes when considering non-significant age-|ARoC| associations 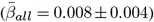

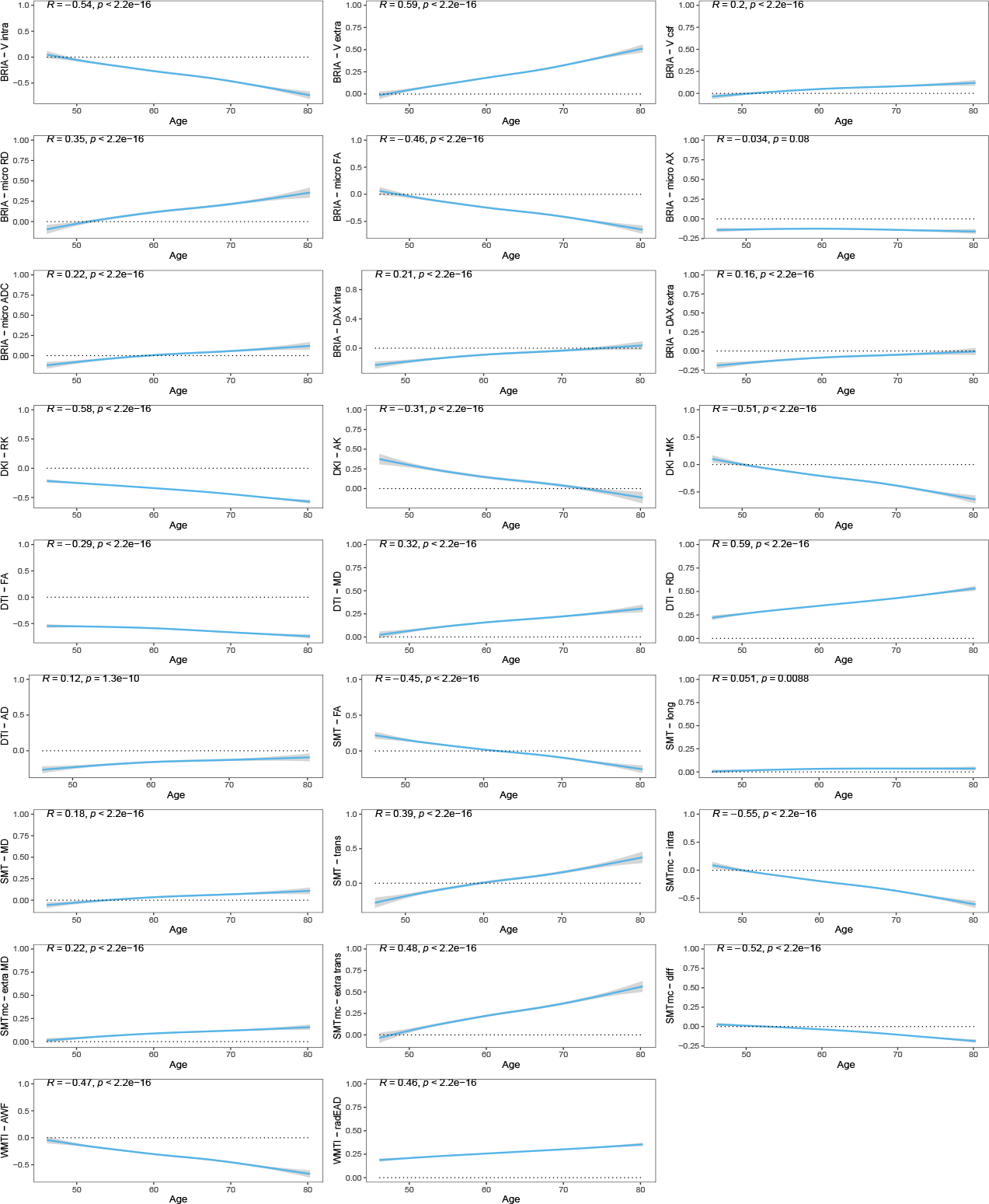

## Supplementary Note 20 Unadjusted annual rate of WMM change throughout ages

WMM are standardised for comparability without mean centering for a better understanding of onset values. Red lines were implemented as visual aid.

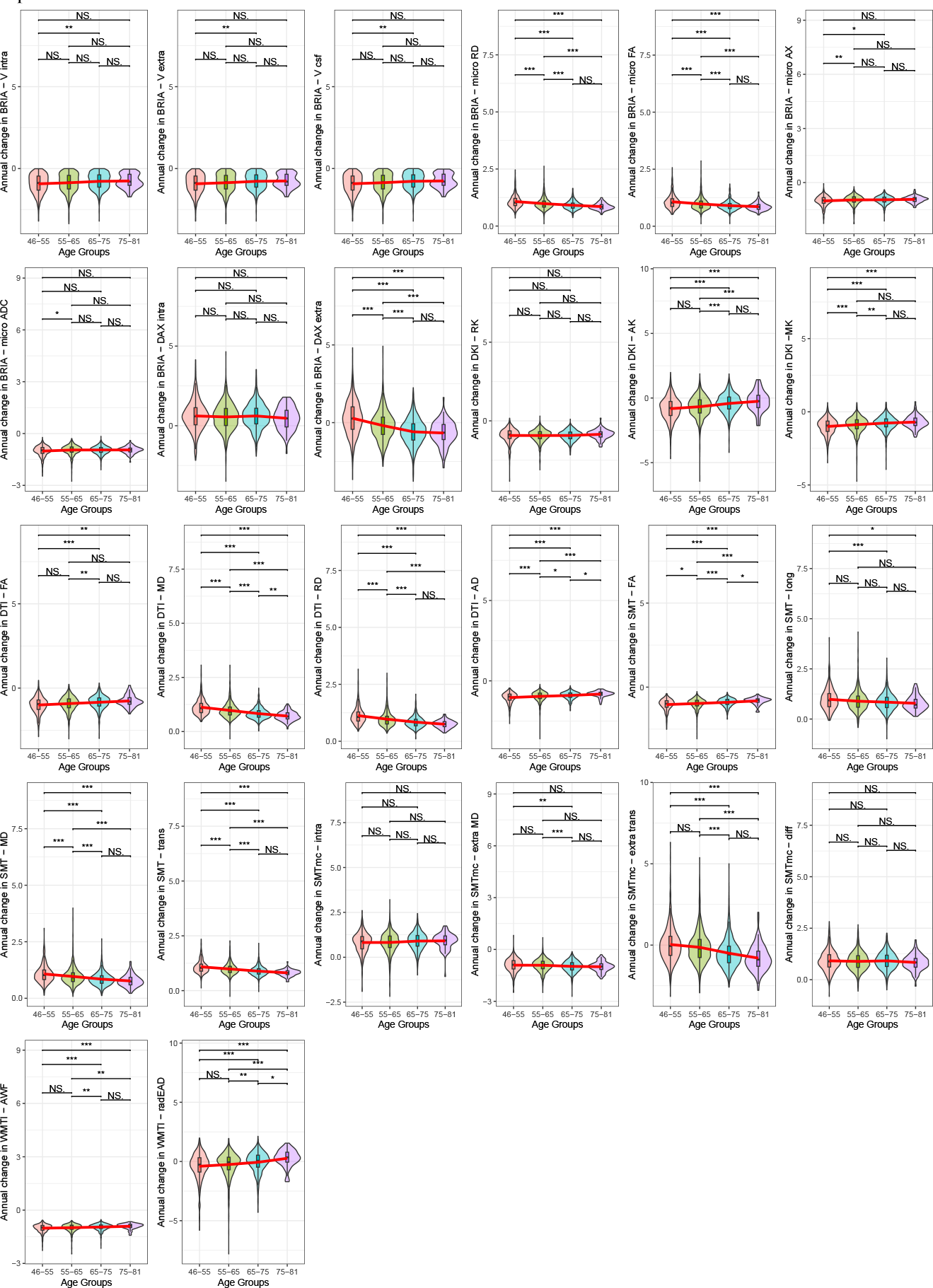

## Supplementary Note 21 Corrected sex- and age-stratified annual rate of WMM change

WMM are standardised for comparability (including mean centering). Red colours indicate females and blue colours males.

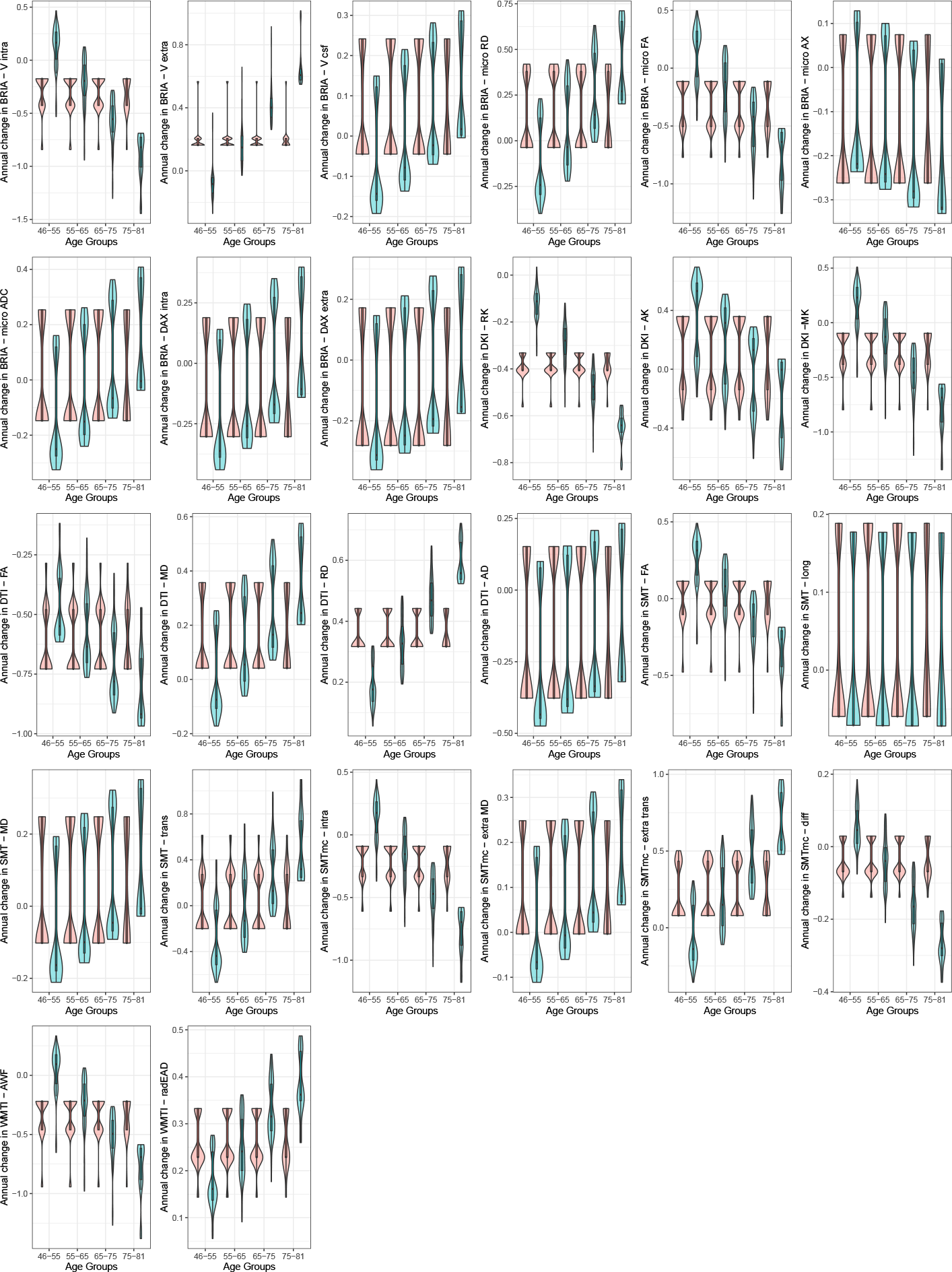

## Supplementary Note 22 Regional associations between WMM change in the Cerebral Peduncle and PGRS

Panel (a) presents the associations between PGRS and WMM change. Panel (b) presents the regional associations between PGRS and cross-sectional regional WMM at time point one. Panel (c) presents the cross-sectional regional WMM associations with PGRS at time point two. Panel (d) presents cross-sectional regional associations between WMM and PGRS for the validation sample. Boxes the statistical significance at an uncorrected *α <* 0.05. Colours indicate the association strength (standardised *β*-coefficients). *Note:* All associations were adjusted for age, sex, the *age × sex* interaction, and site. None of the presented associations survived the adjustment of the *α*-level for multiple comparisons.

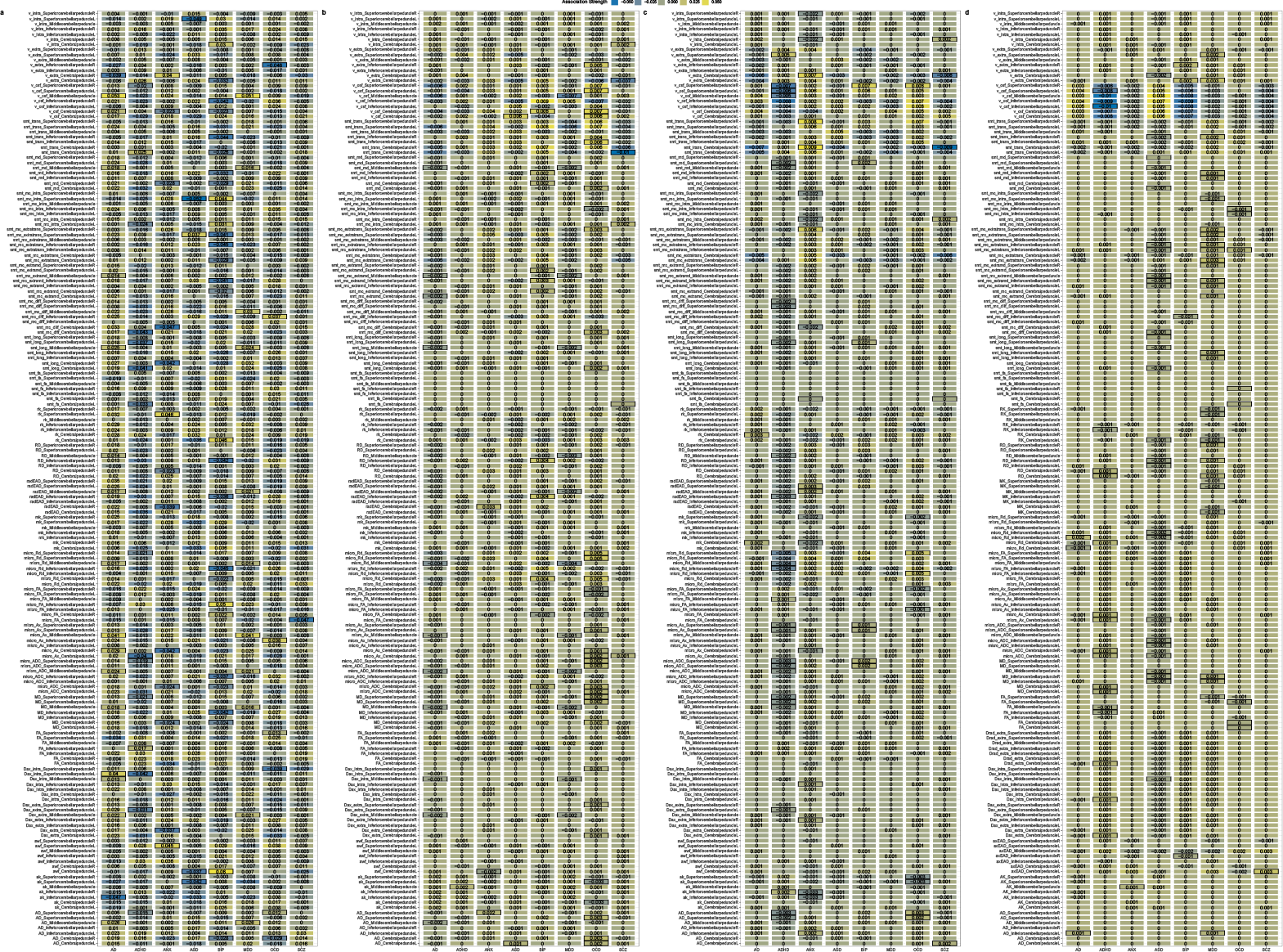

## Supplementary Note 23 Regional associations between WMM change in the Fornix and PGRS

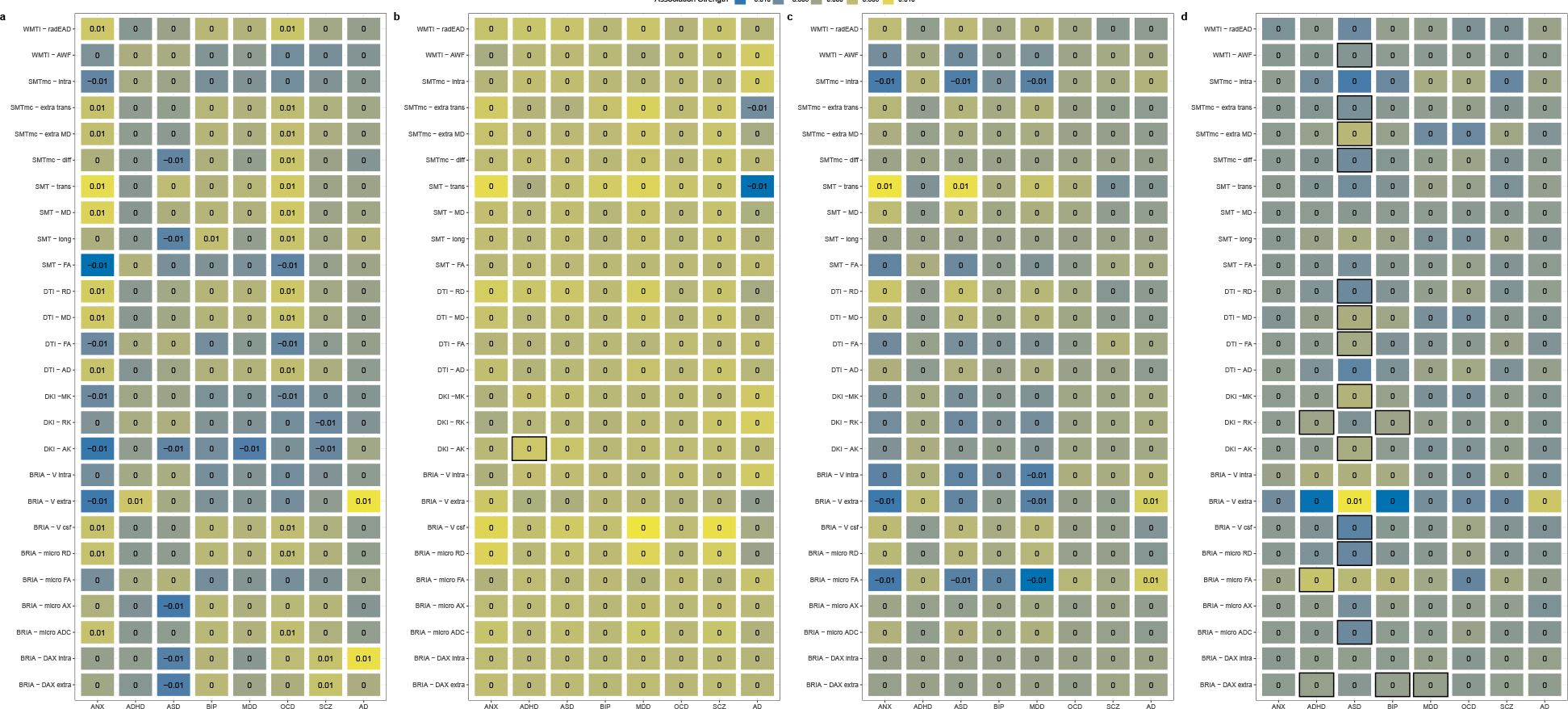

## Supplementary Note 24 Distribution of the relationship between PGRS and both the annual rate of WMM change as well as cross-sectional WMM

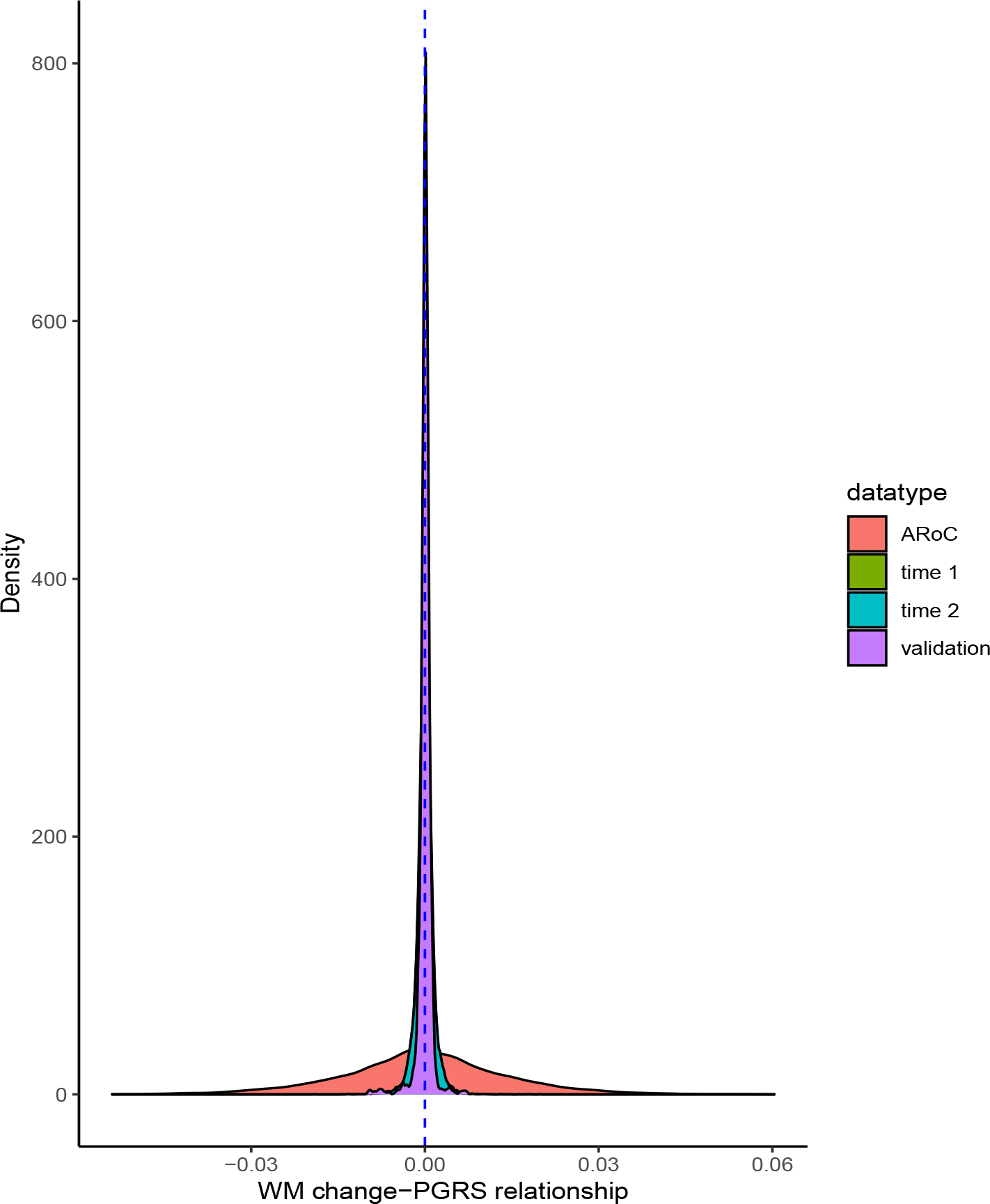

## Supplementary Note 25 Voxel-level changes in BRIA - DAX extra

Red to yellow colour indicate increases, dark to light blue colours decreases in, respectively, where the scale is determined by the *α*-level ranging from 0.05 towards 0. MNI152 X:0, Y:-18, Z: 18 are indicated by the cursor with slices spacing of *±* 4 steps along the z-axis in the coordinate system.

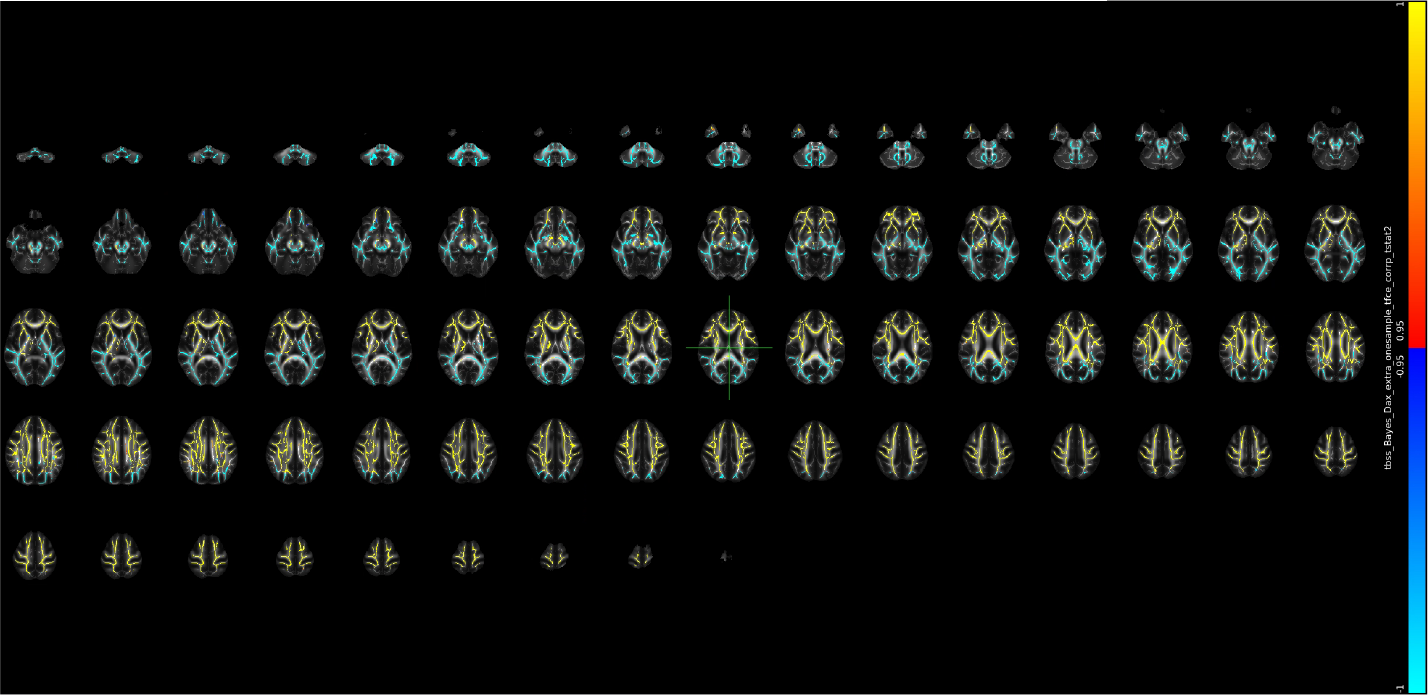

## Supplementary Note 26 Voxel-level changes in BRIA - DAX intra

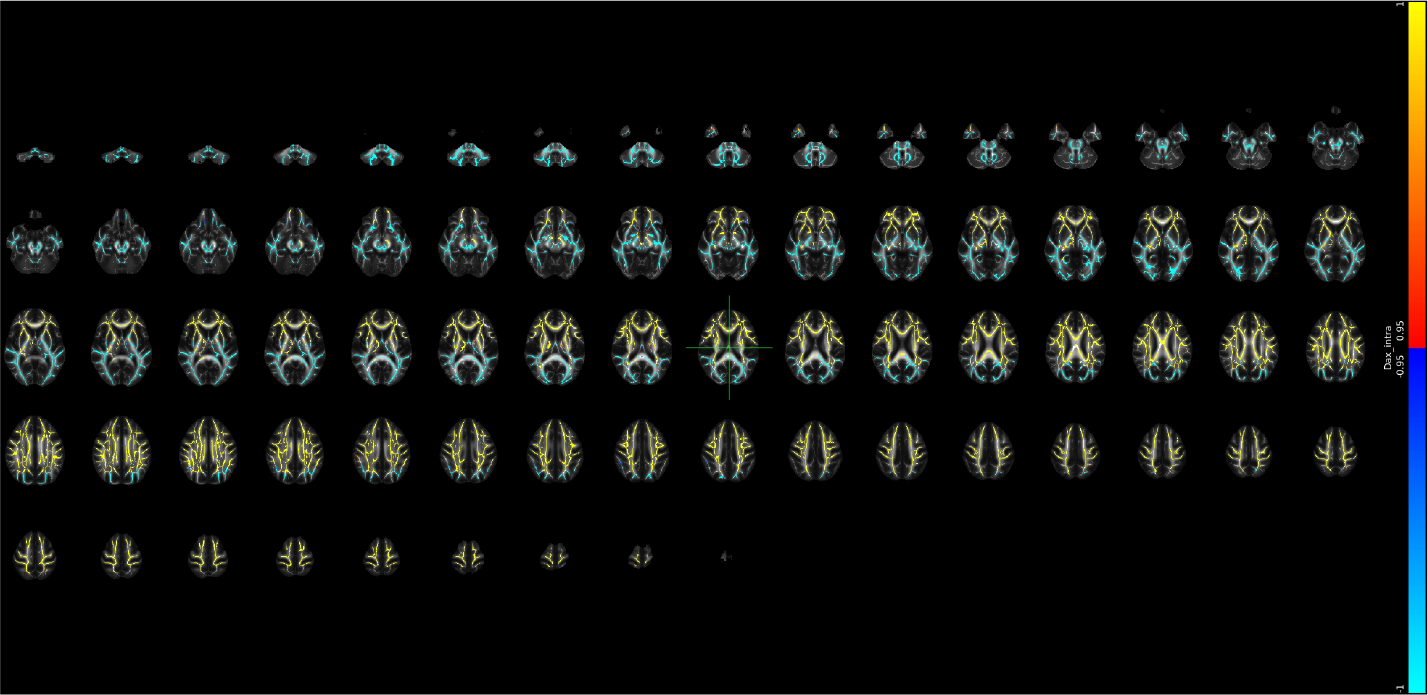

## Supplementary Note 27 Voxel-level changes in BRIA - DAX intra

Red to yellow colour indicate increases, dark to light blue colours decreases in, respectively, where the scale is determined by the *α*-level ranging from 0.05 towards 0. MNI152 X:0, Y:-18, Z: 18 are indicated by the cursor with slices spacing of *±*4 steps along the z-axis in the coordinate system.

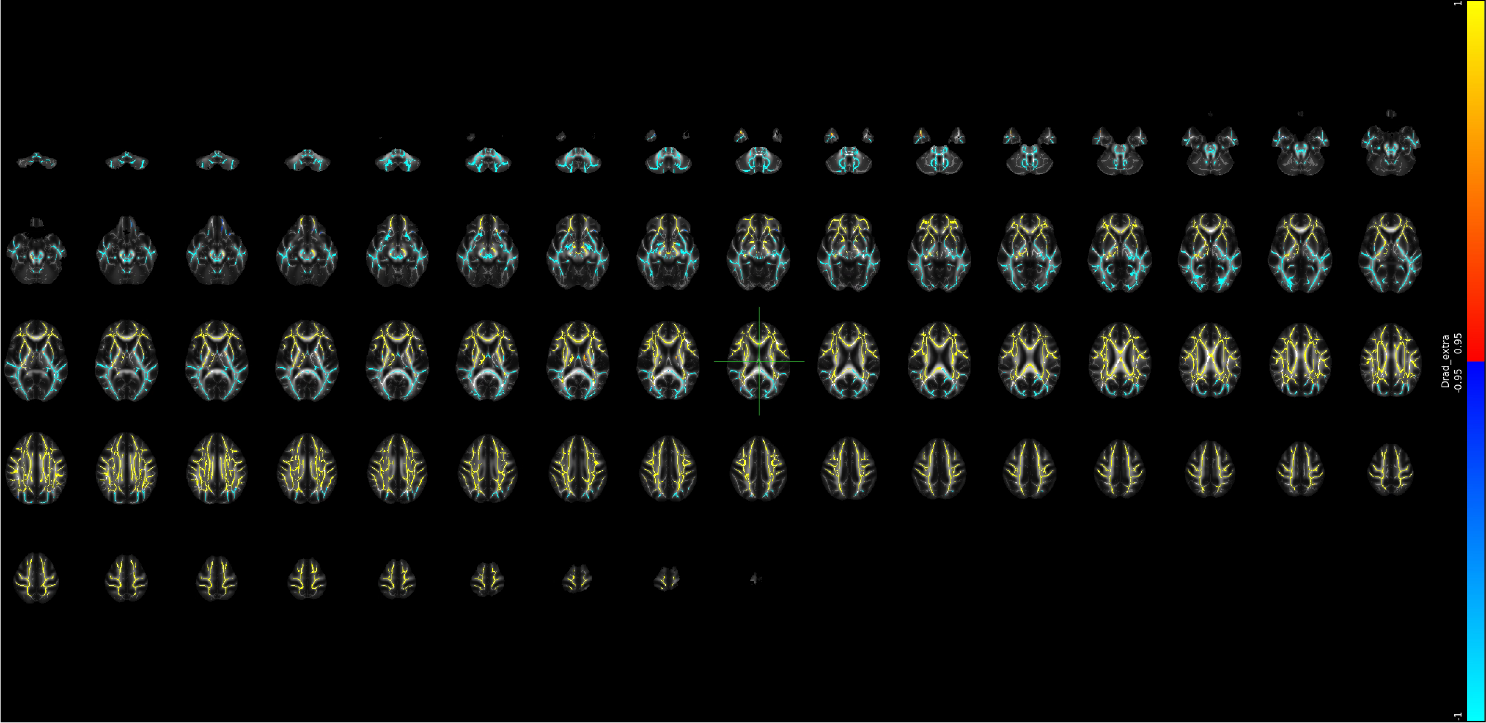

## Supplementary Note 28 Voxel-level changes in BRIA - microADC

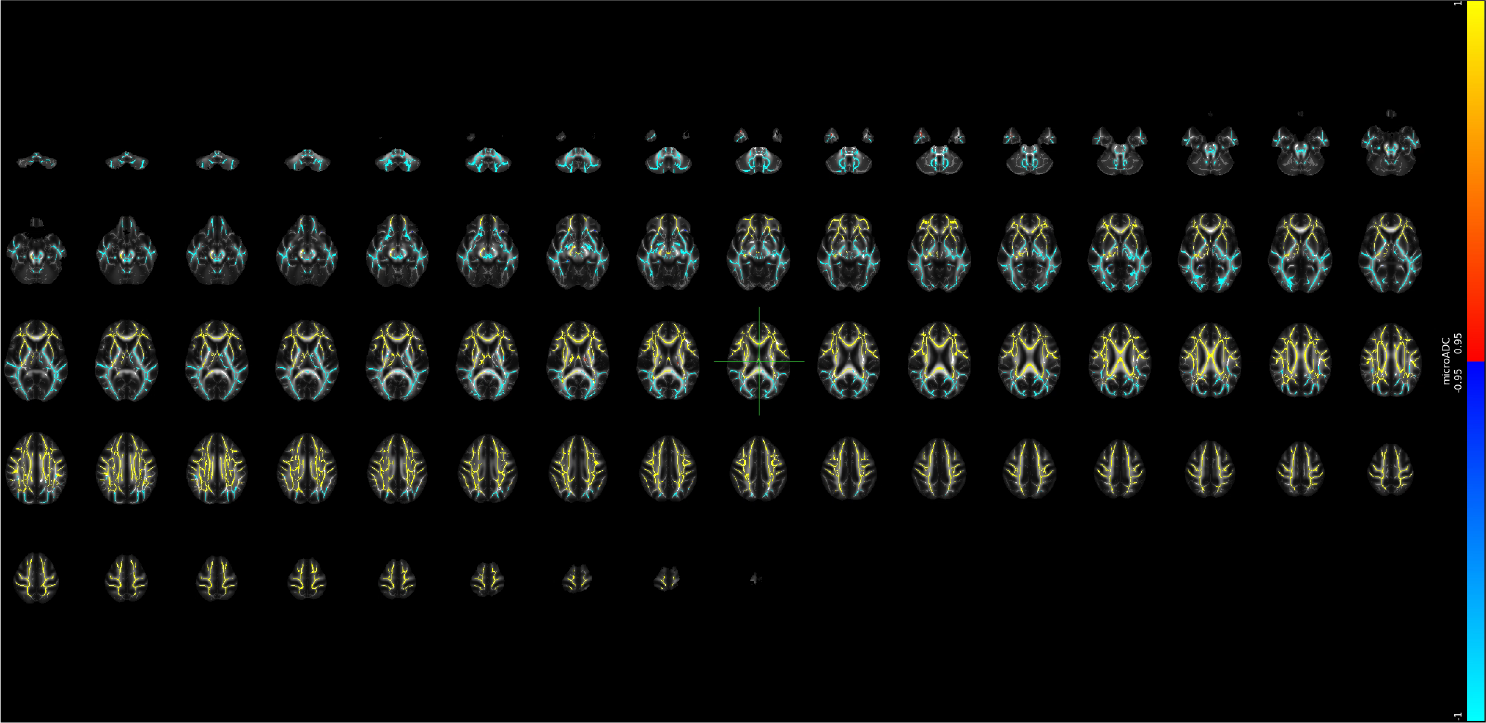

## Supplementary Note 29 Voxel-level changes in BRIA - microAX

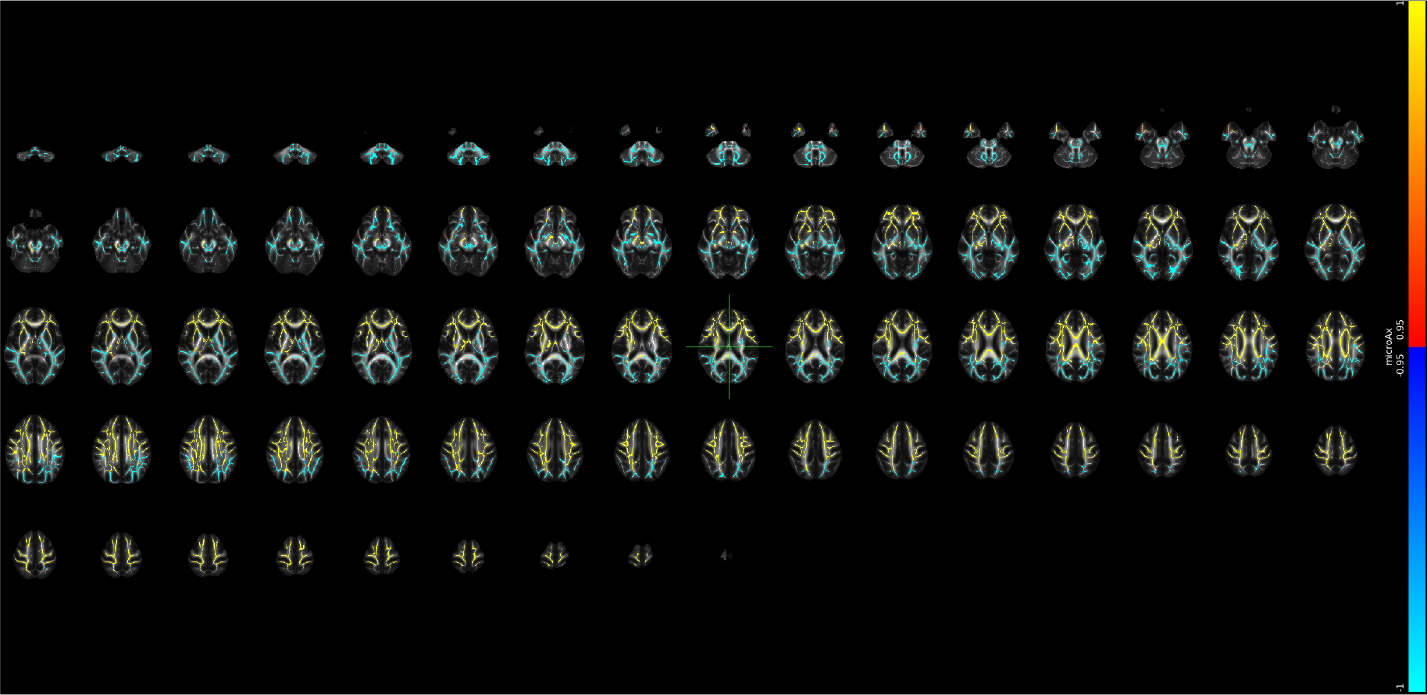

## Supplementary Note 30 Voxel-level changes in BRIA - microFA

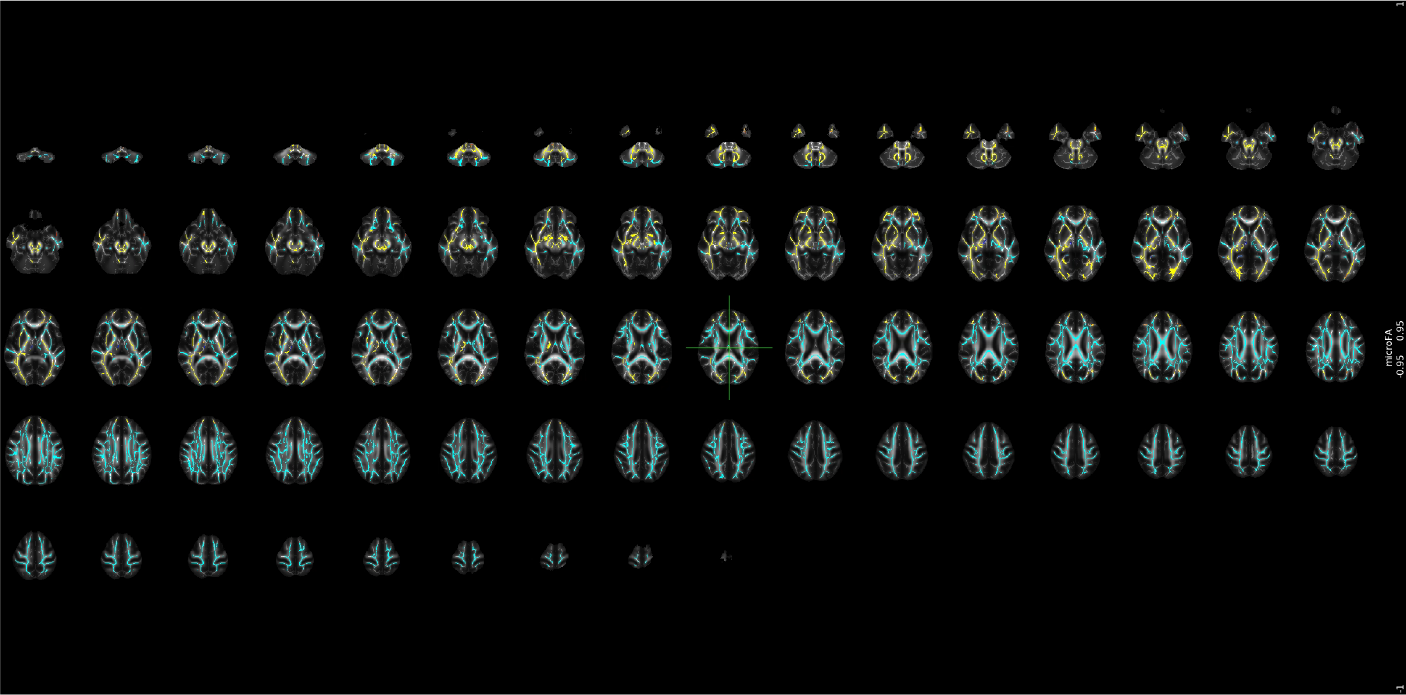

## Supplementary Note 31 Voxel-level changes in BRIA - microRD

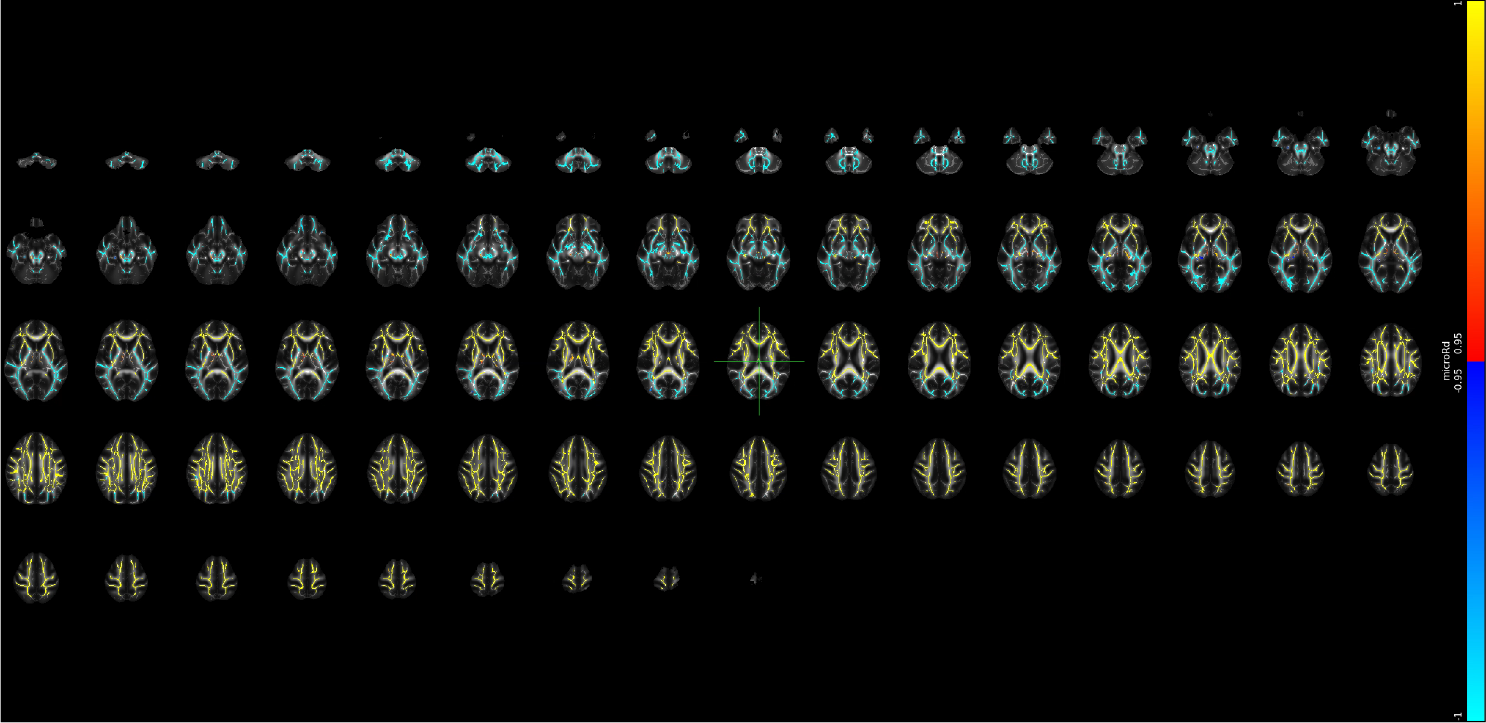

## Supplementary Note 32 Voxel-level changes in BRIA - Vcsf

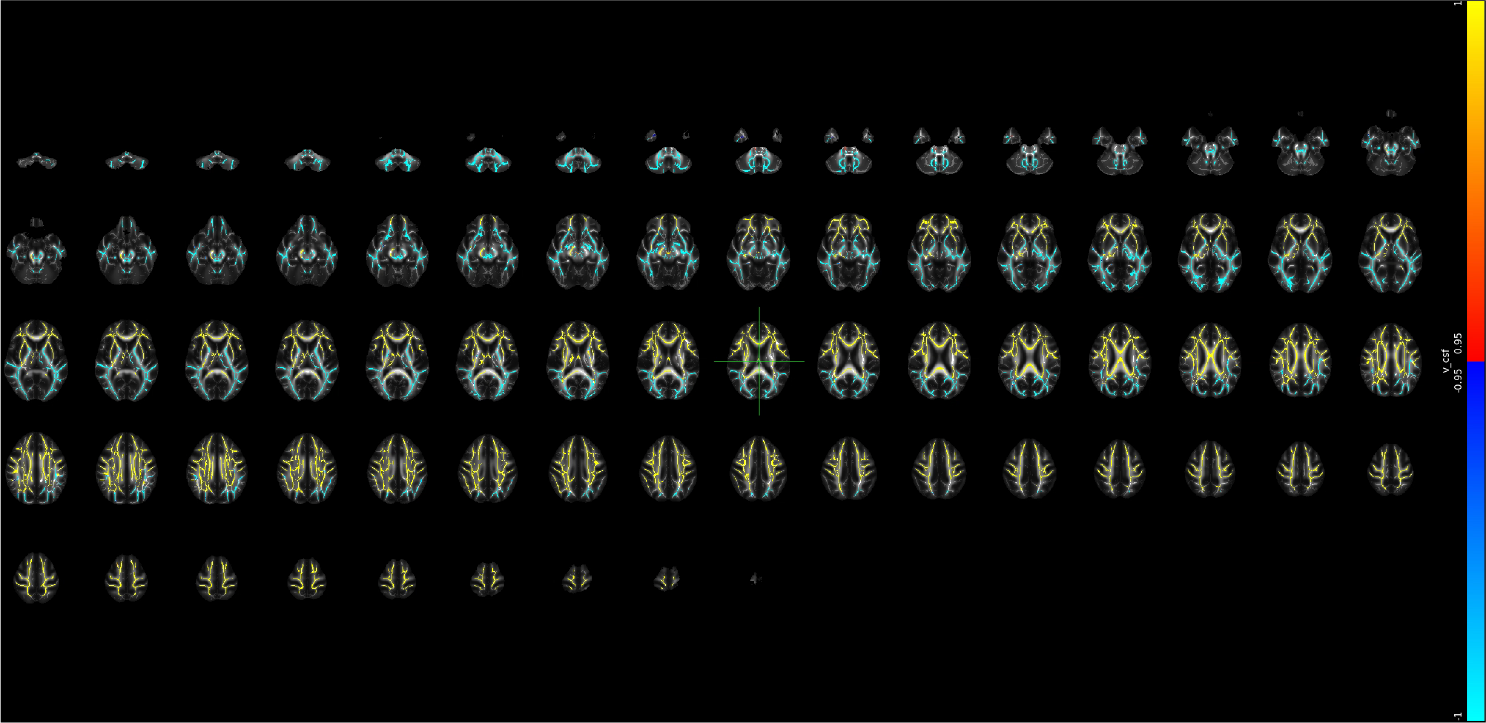

## Supplementary Note 33 Voxel-level changes in BRIA - Vextra

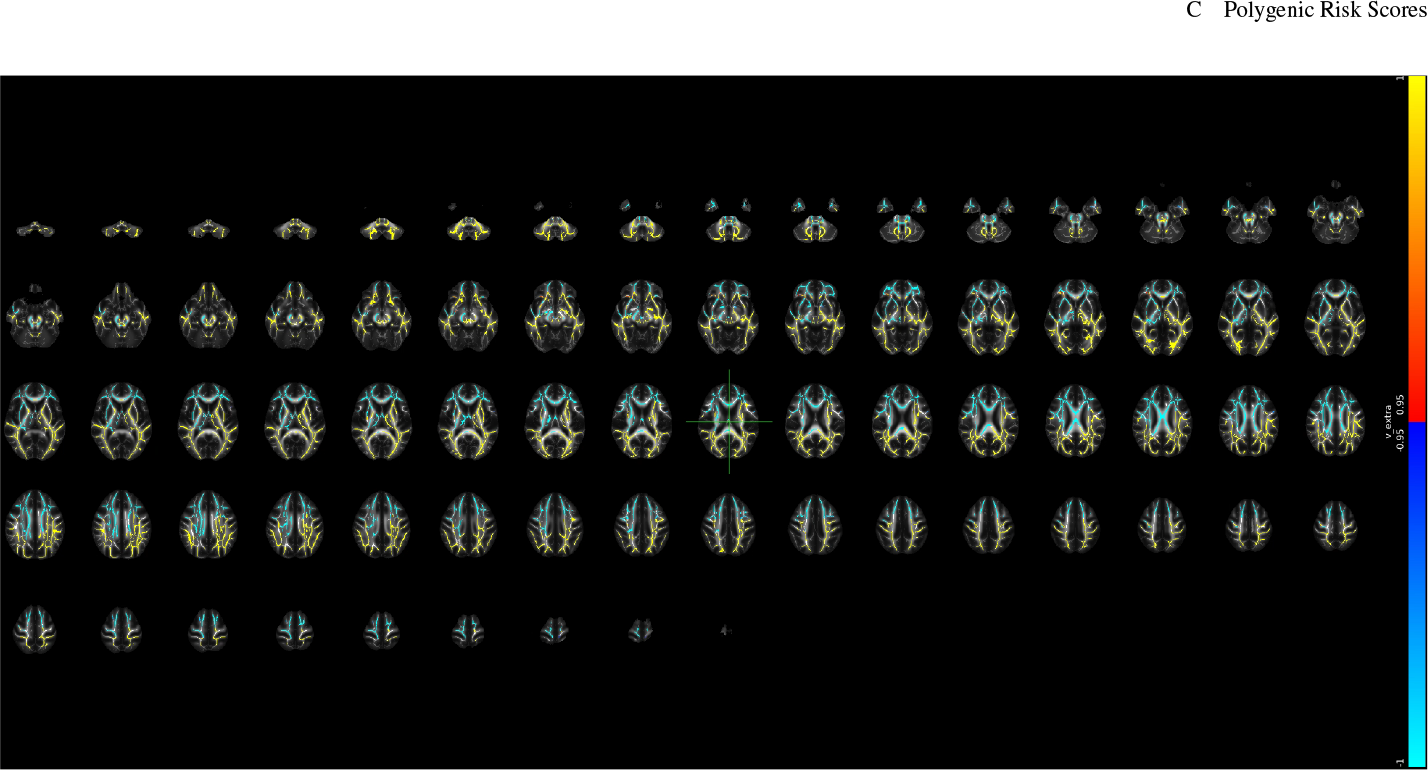

## Supplementary Note 34 Voxel-level changes in BRIA - Vintra

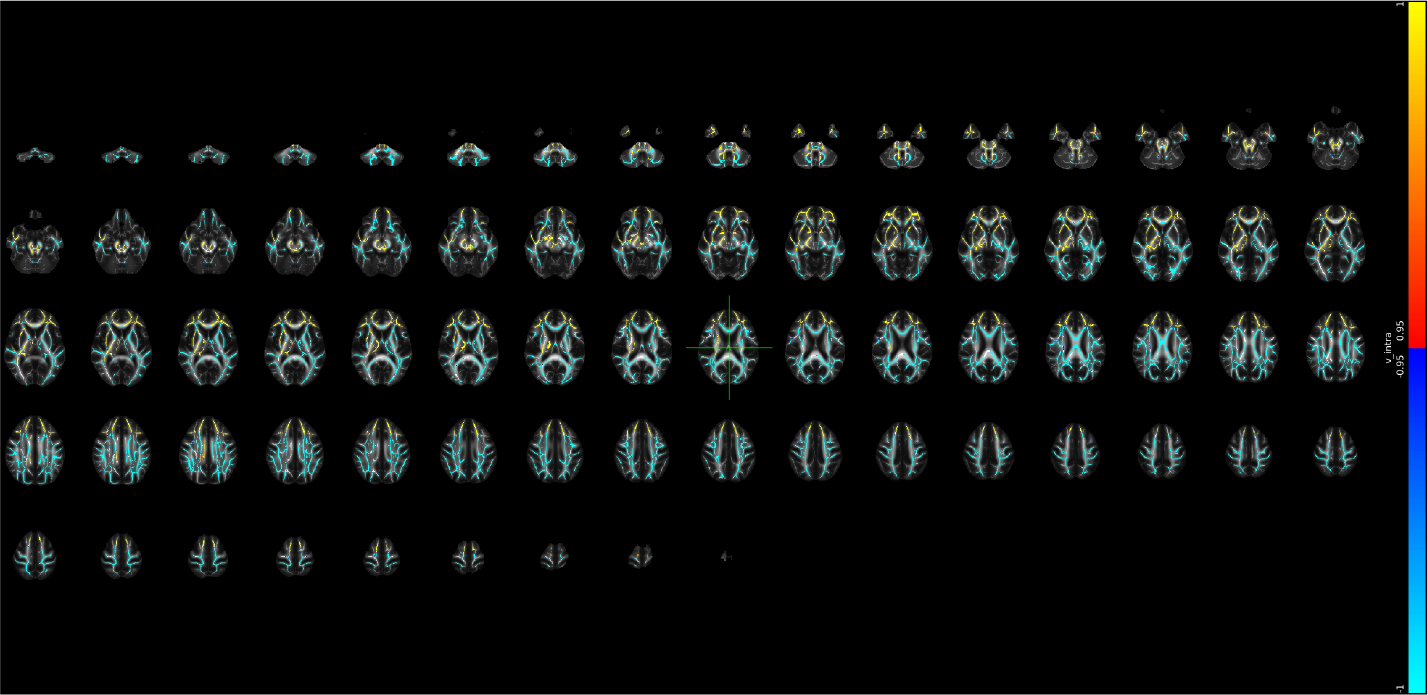

## Supplementary Note 35 Voxel-level changes in DKI - AK

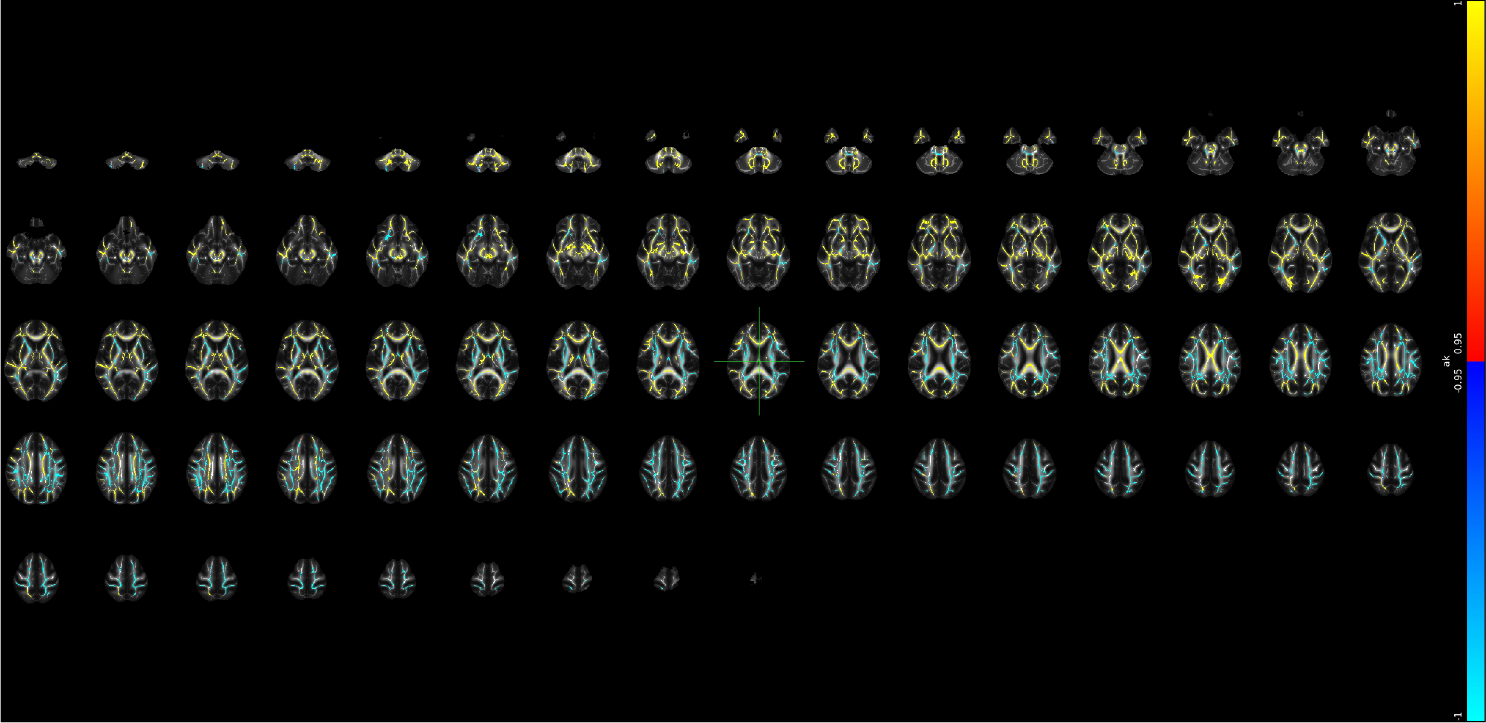

## Supplementary Note 36 Voxel-level changes in DKI - MK

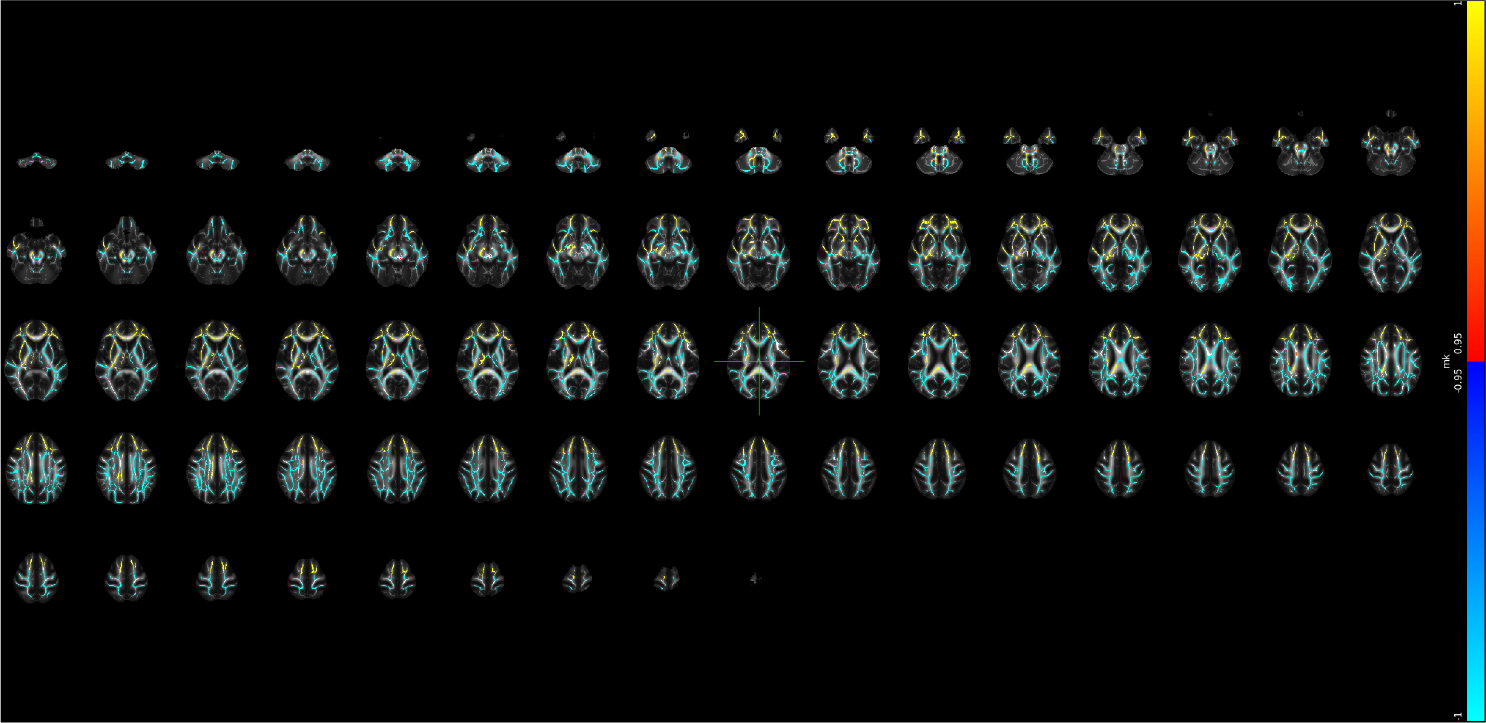

## Supplementary Note 37 Voxel-level changes in DKI - RK

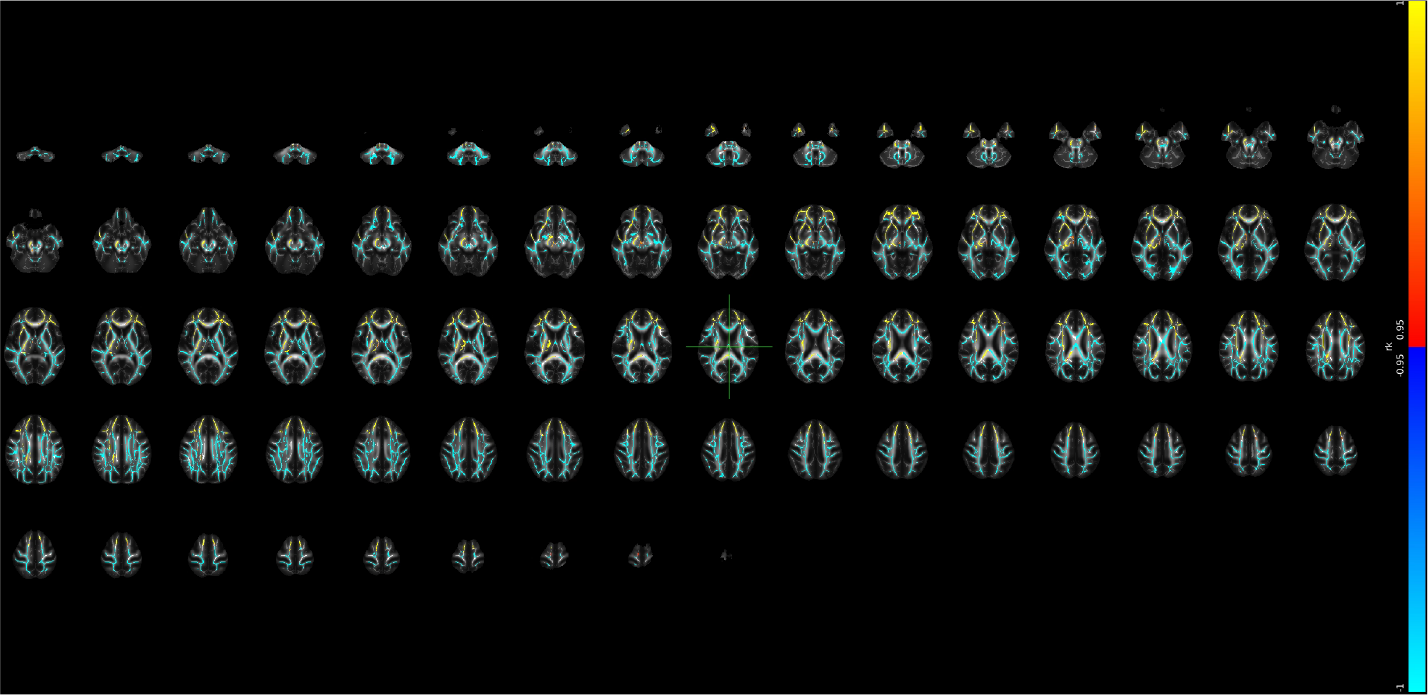

## Supplementary Note 38 Voxel-level changes in DTI - AD

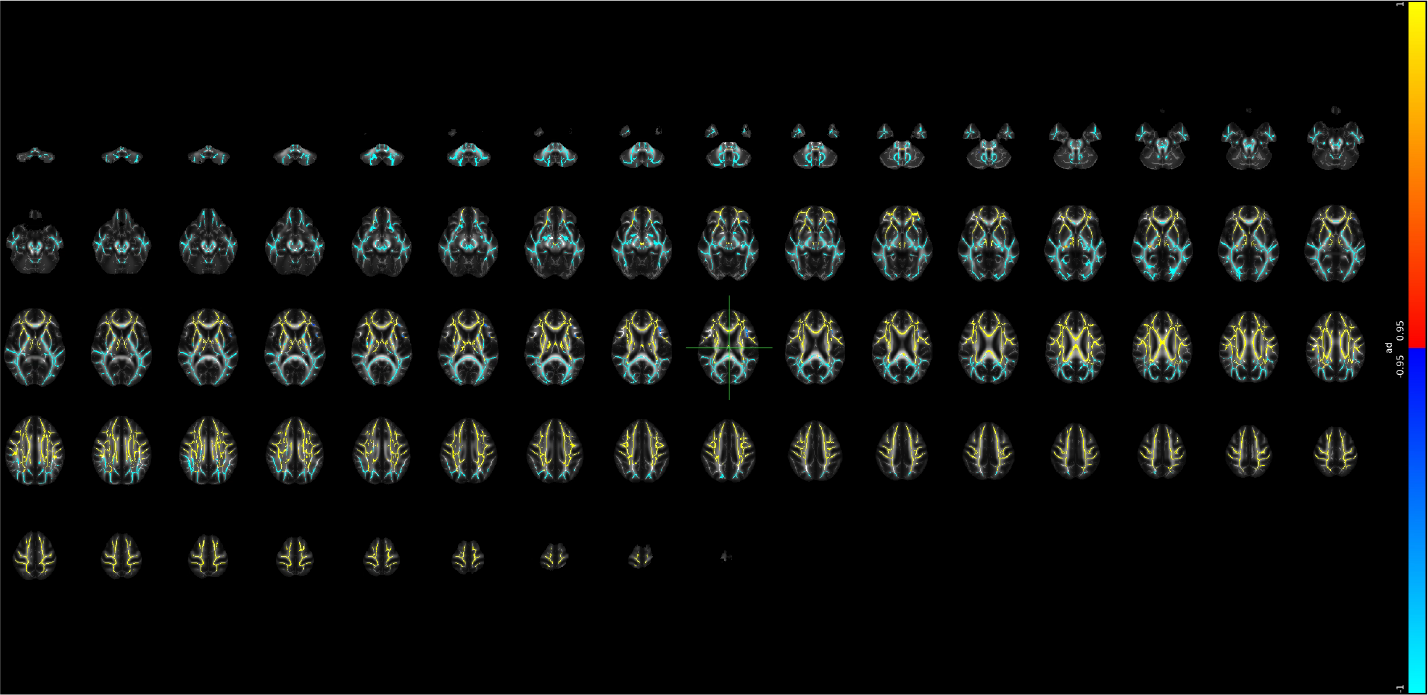

## Supplementary Note 39 Voxel-level changes in DTI - FA

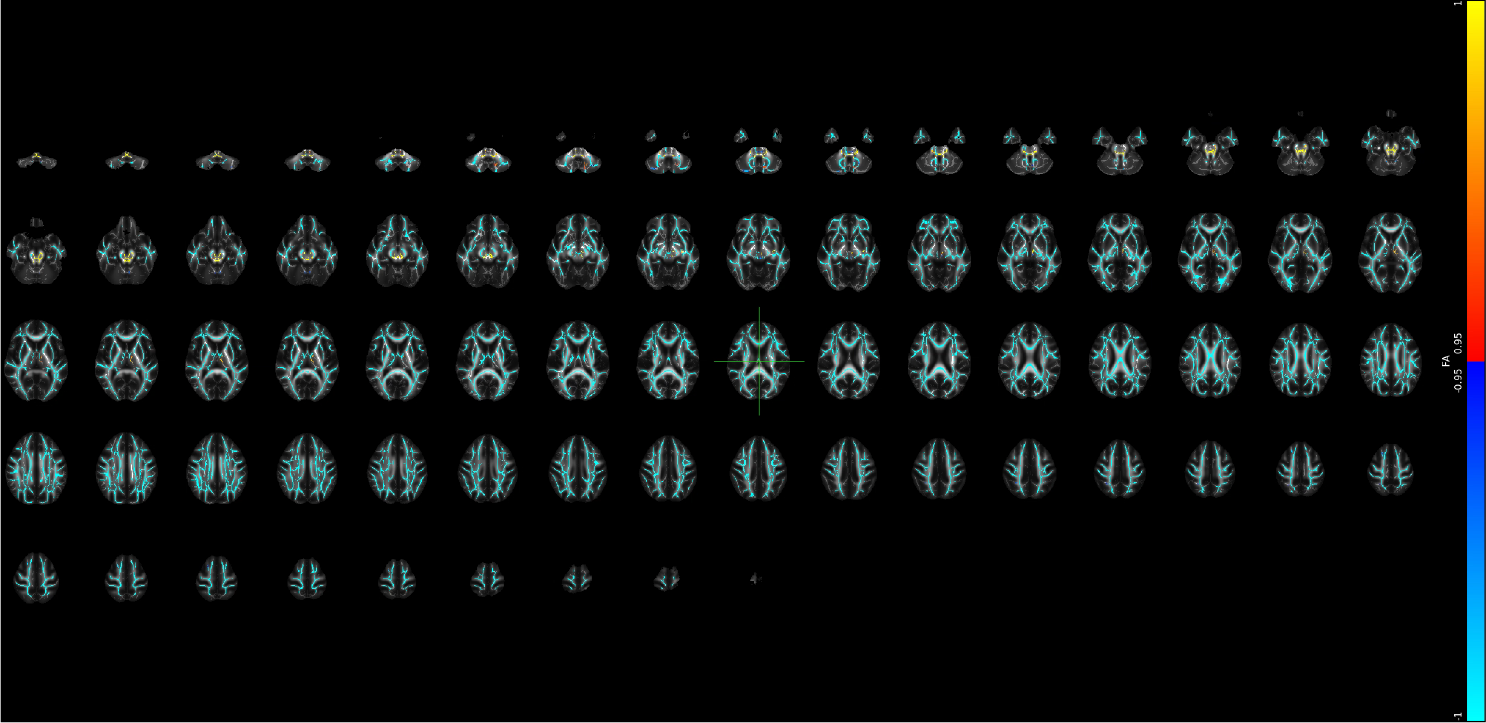

## Supplementary Note 40 Voxel-level changes in DTI - MD

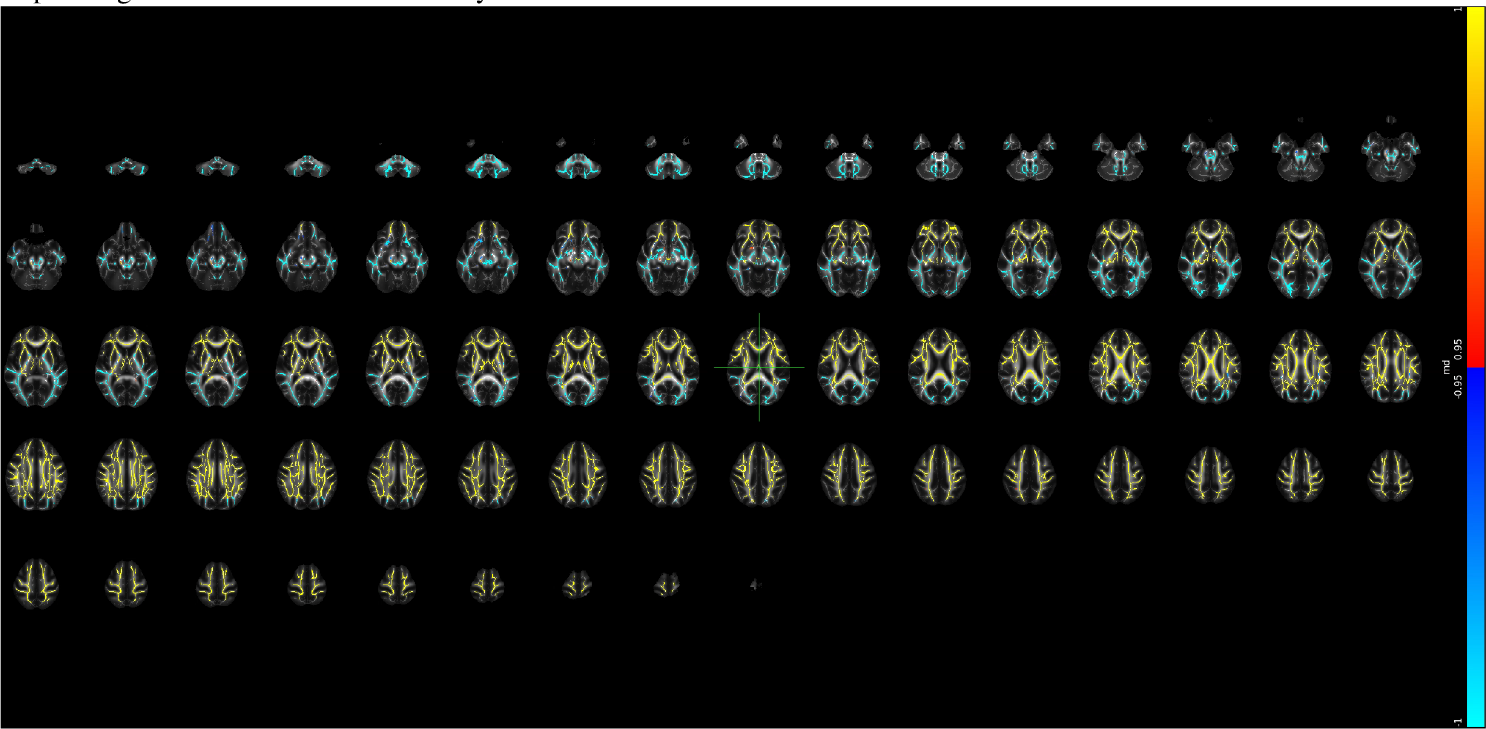

## Supplementary Note 41 Voxel-level changes in DTI - RD

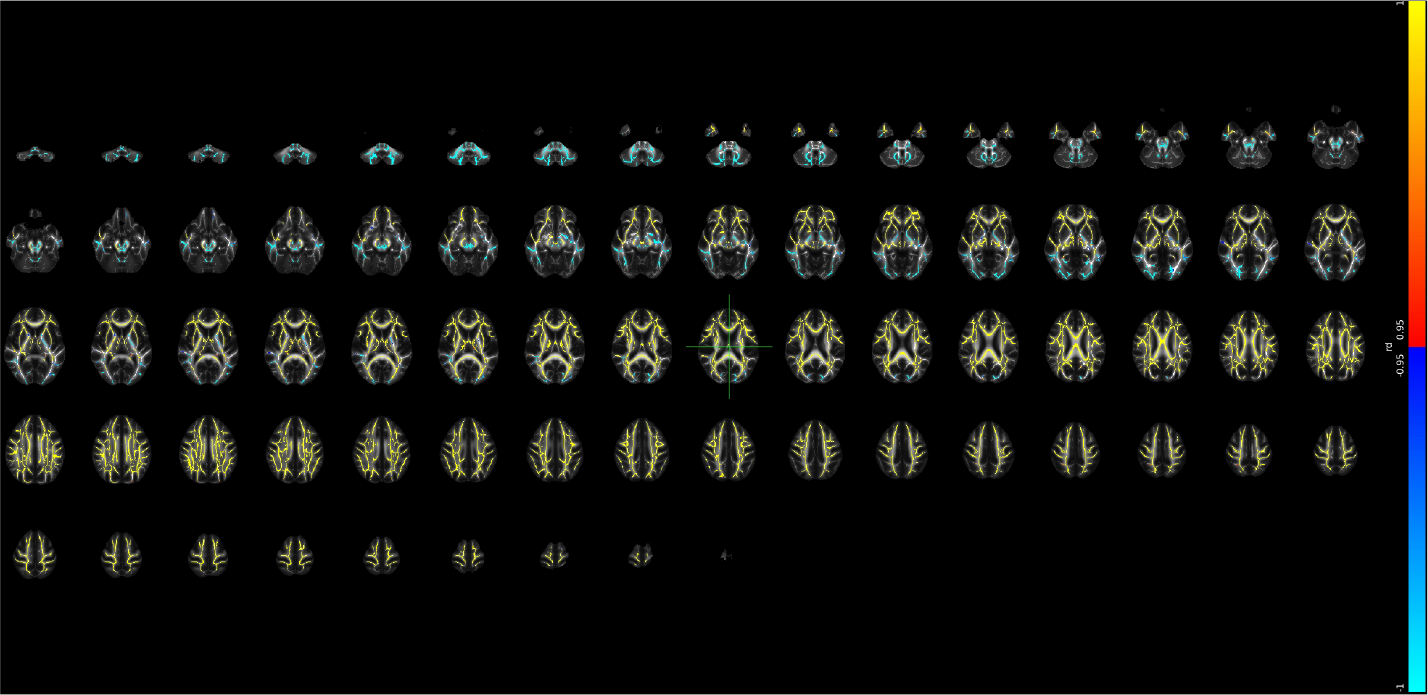

## Supplementary Note 42 Voxel-level changes in SMTmc - diffusion coefficient

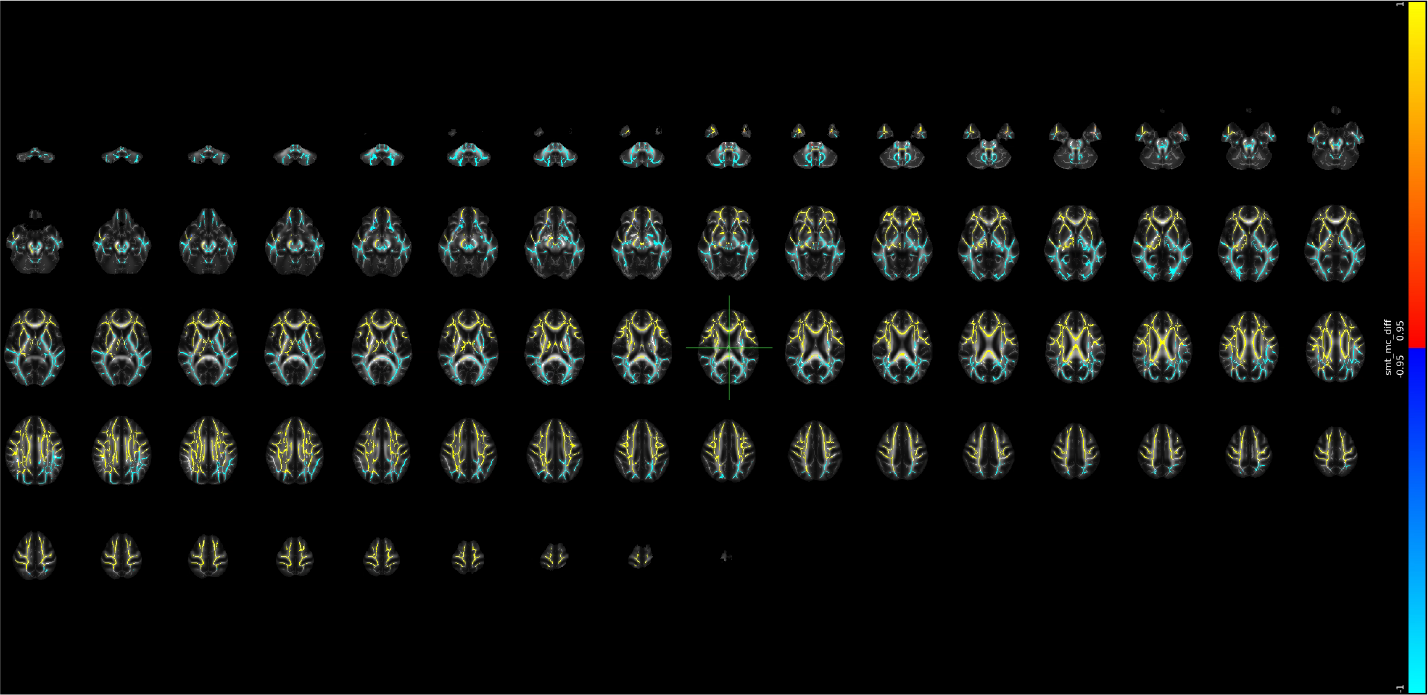

## Supplementary Note 43 Voxel-level changes in SMTmc - extra MD

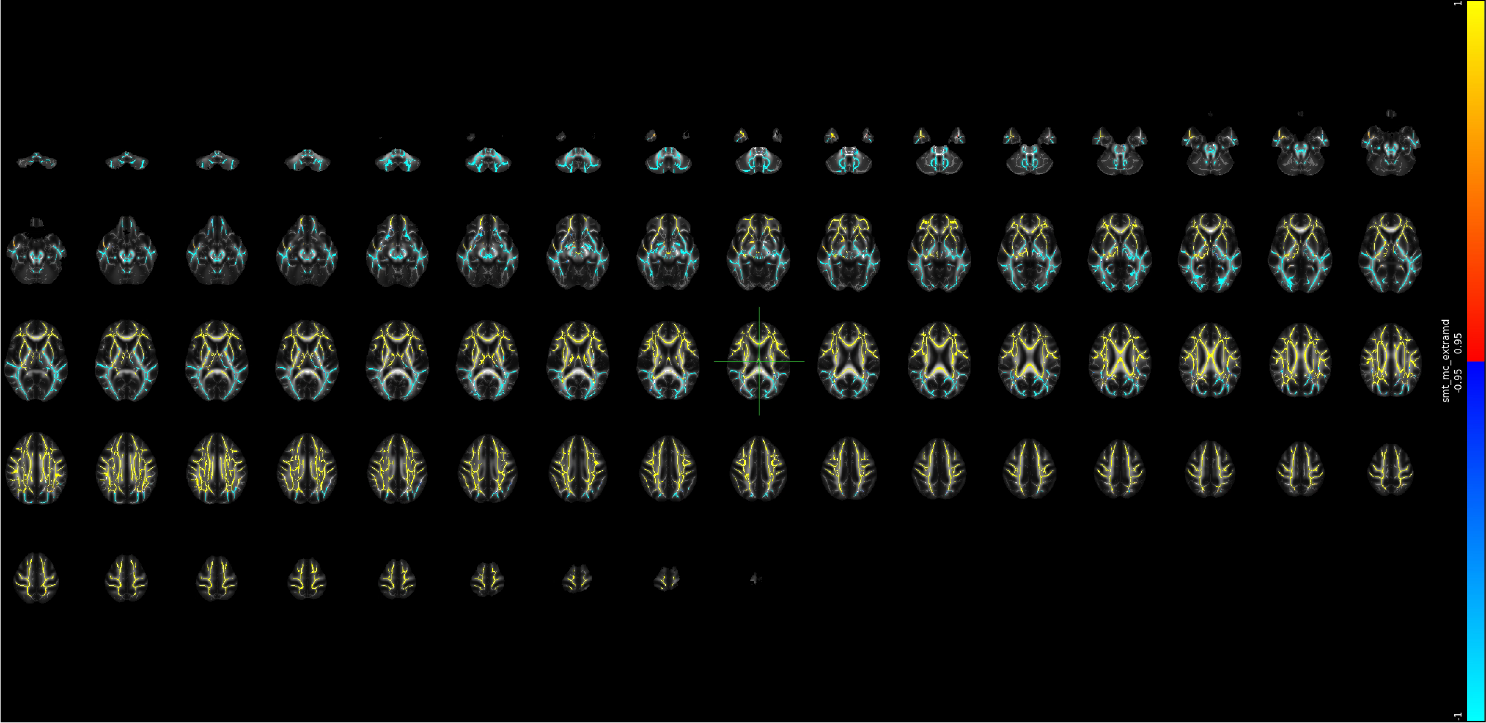

## Supplementary Note 44 Voxel-level changes in SMTmc - extra trans

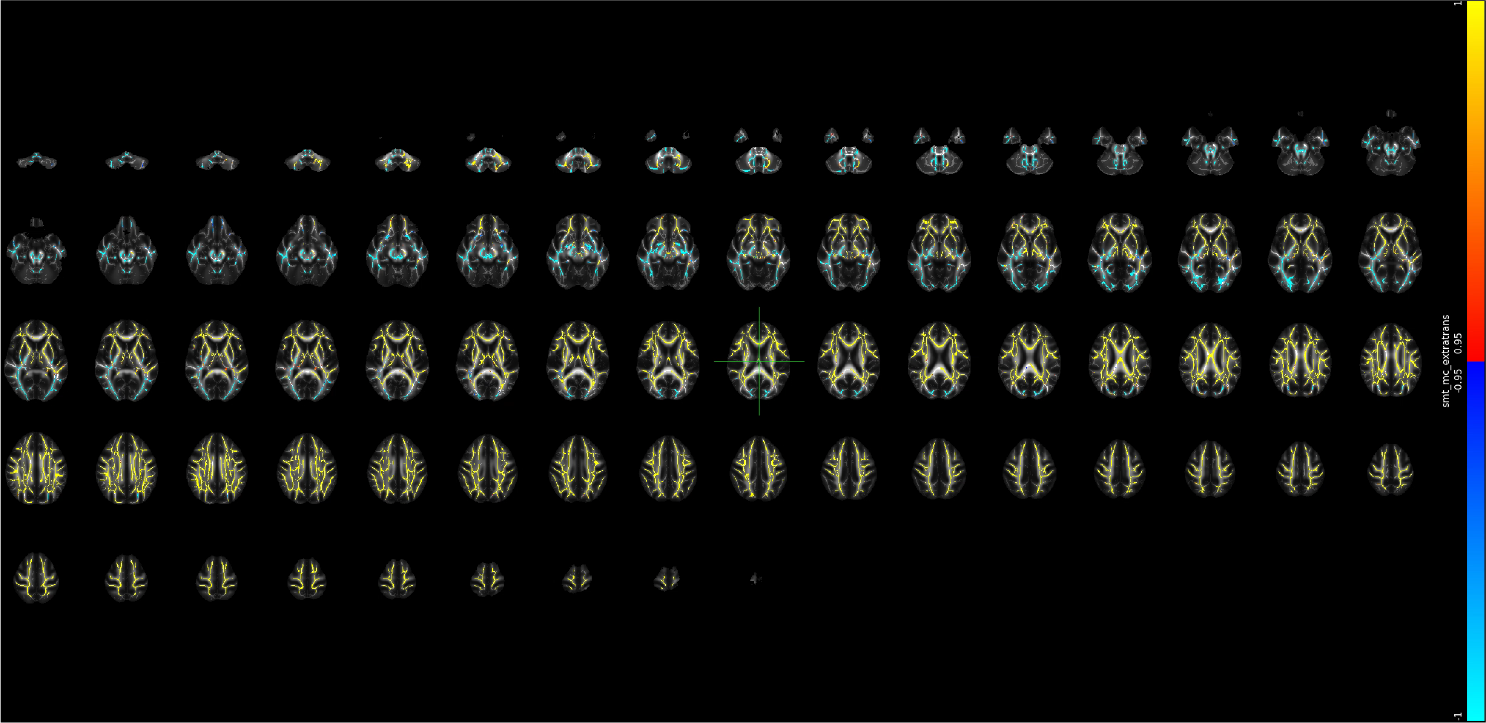

## Supplementary Note 45 Voxel-level changes in SMTmc - intra

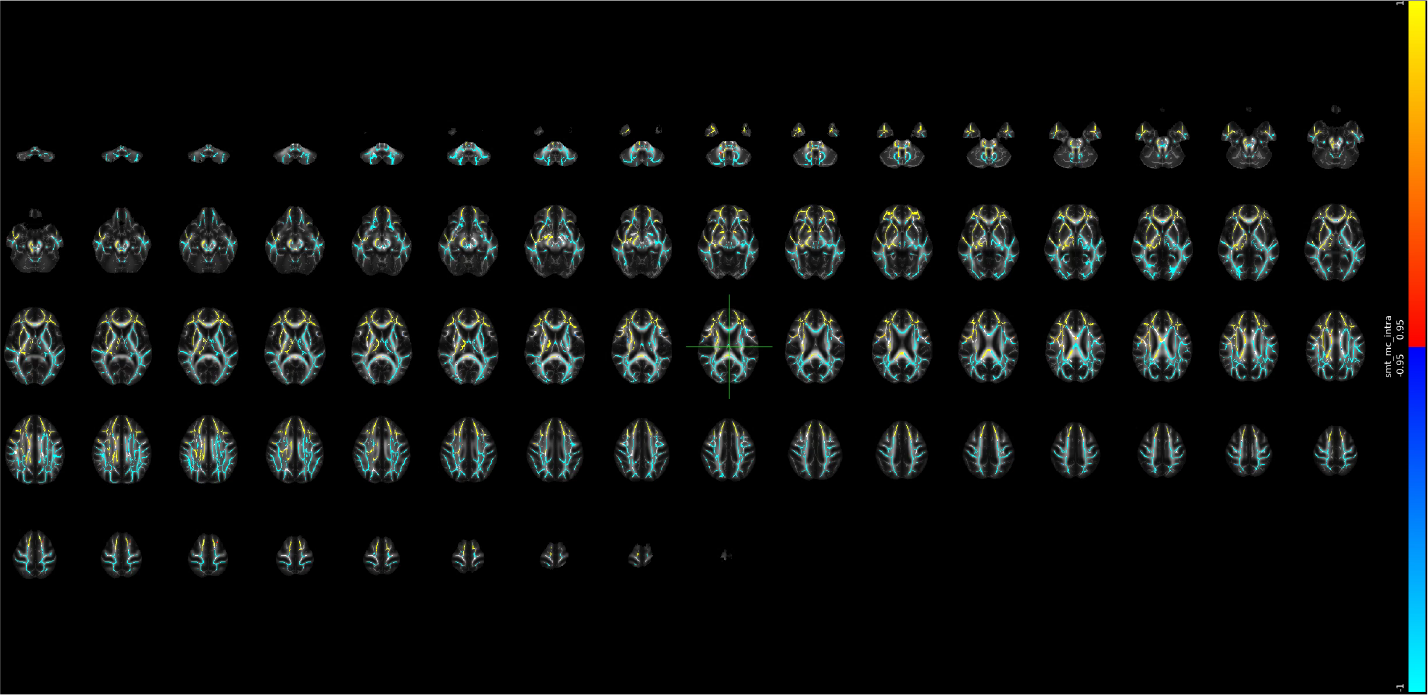

## Supplementary Note 46 Voxel-level changes in WMT - AWF

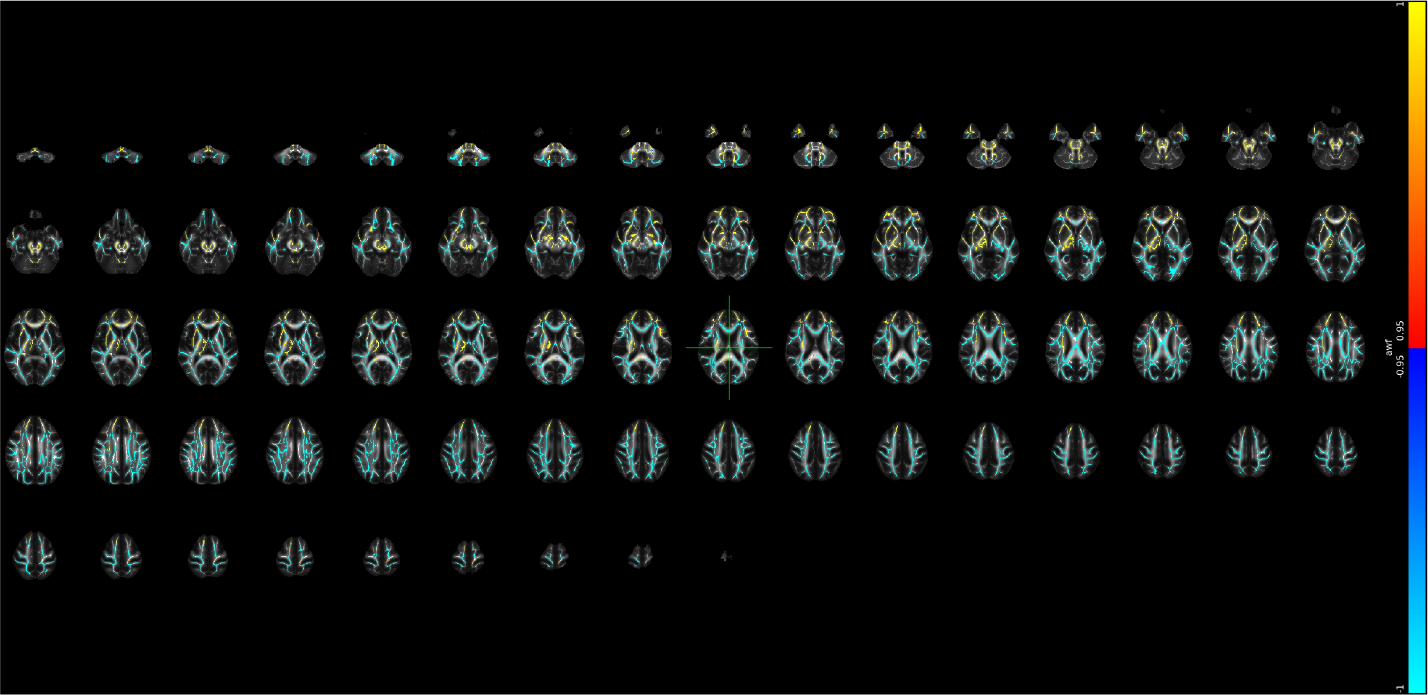

## Supplementary Note 47 Voxel-level changes in WMTI - axEAD

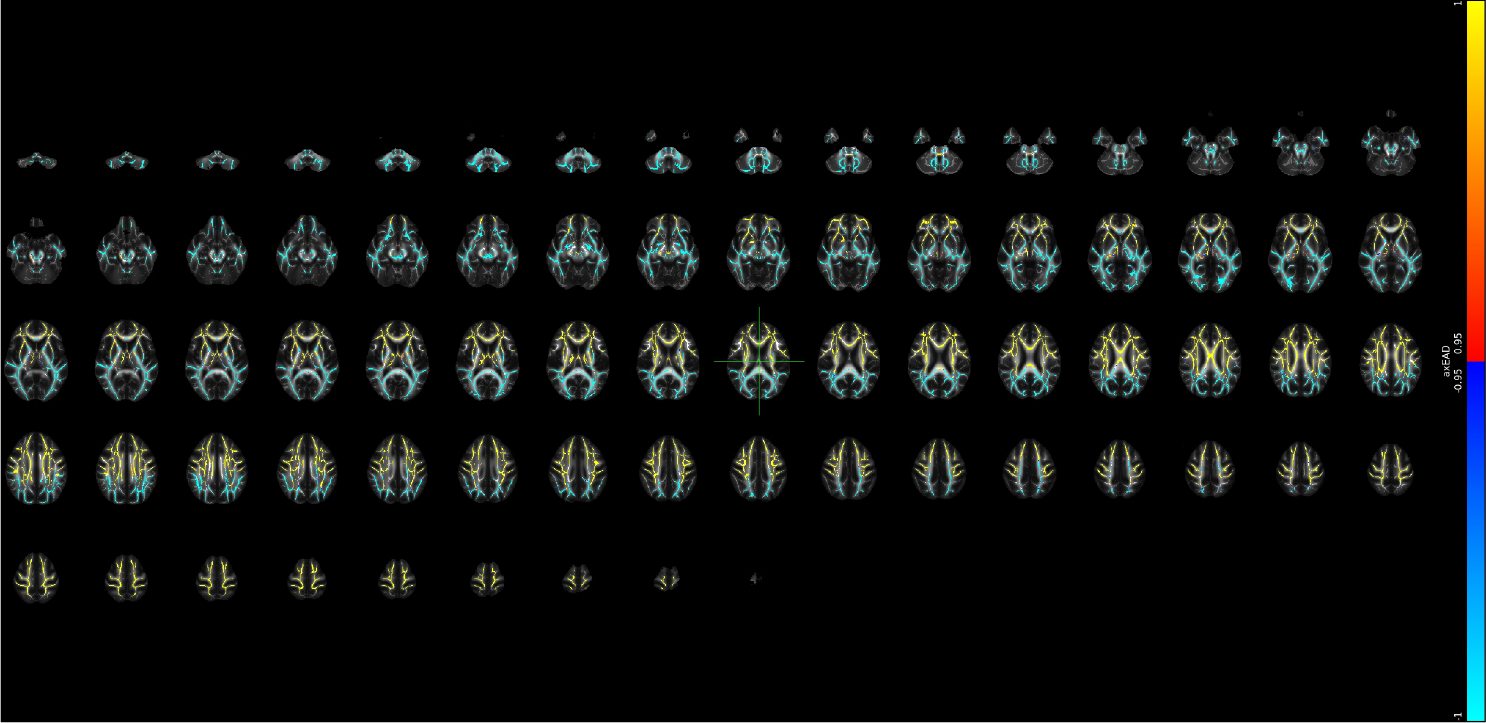

## Supplementary Note 48 Voxel-level changes in WMTI - radEAD

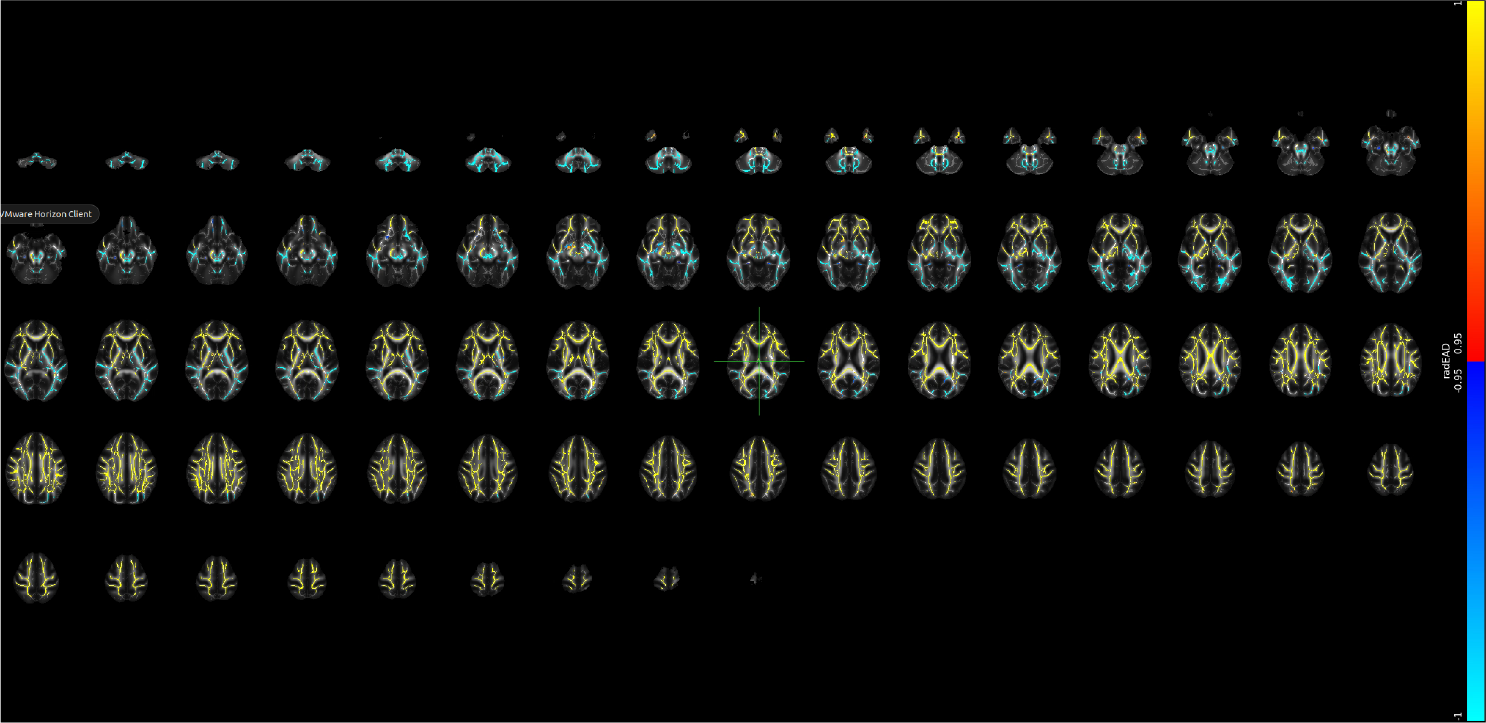

## Supplementary Note 49 Voxel-level changes in SMT - FA

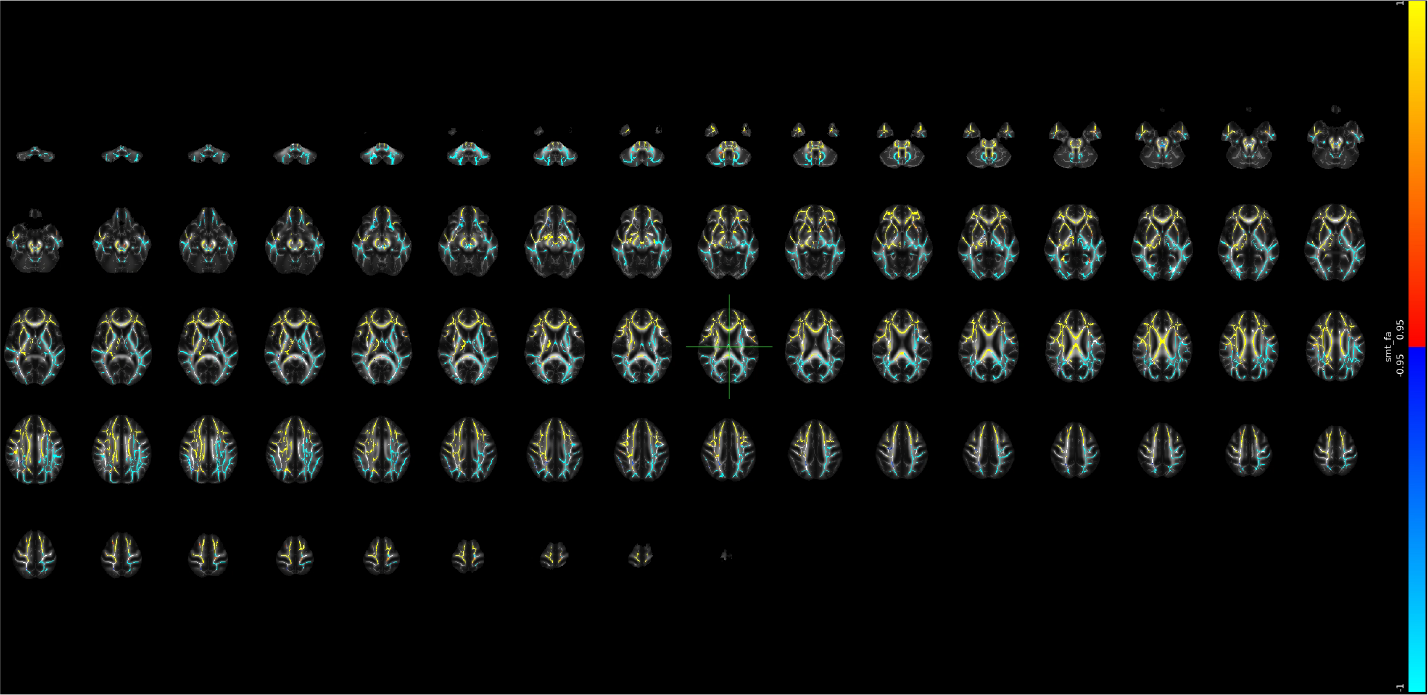

## Supplementary Note 50 Voxel-level changes in SMT - MD

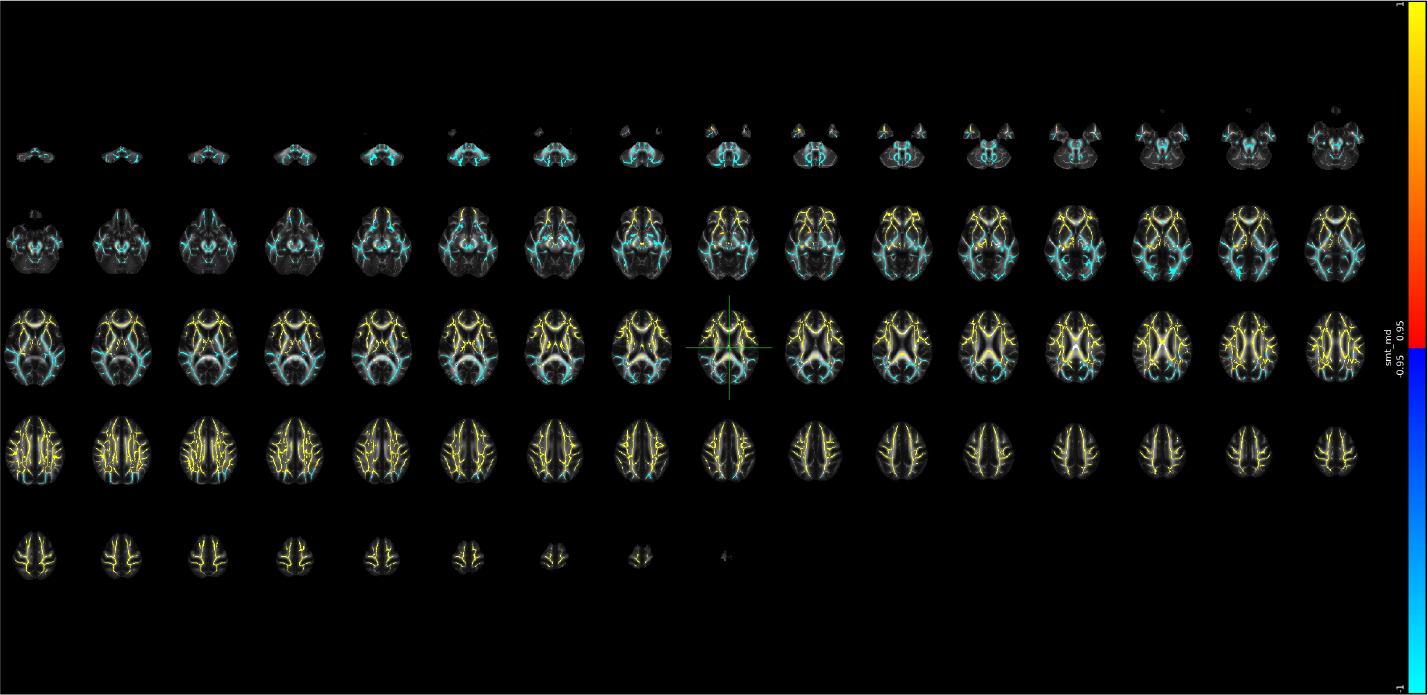

## Supplementary Note 51 Voxel-level changes in SMT - longitudinal diffusion coefficient

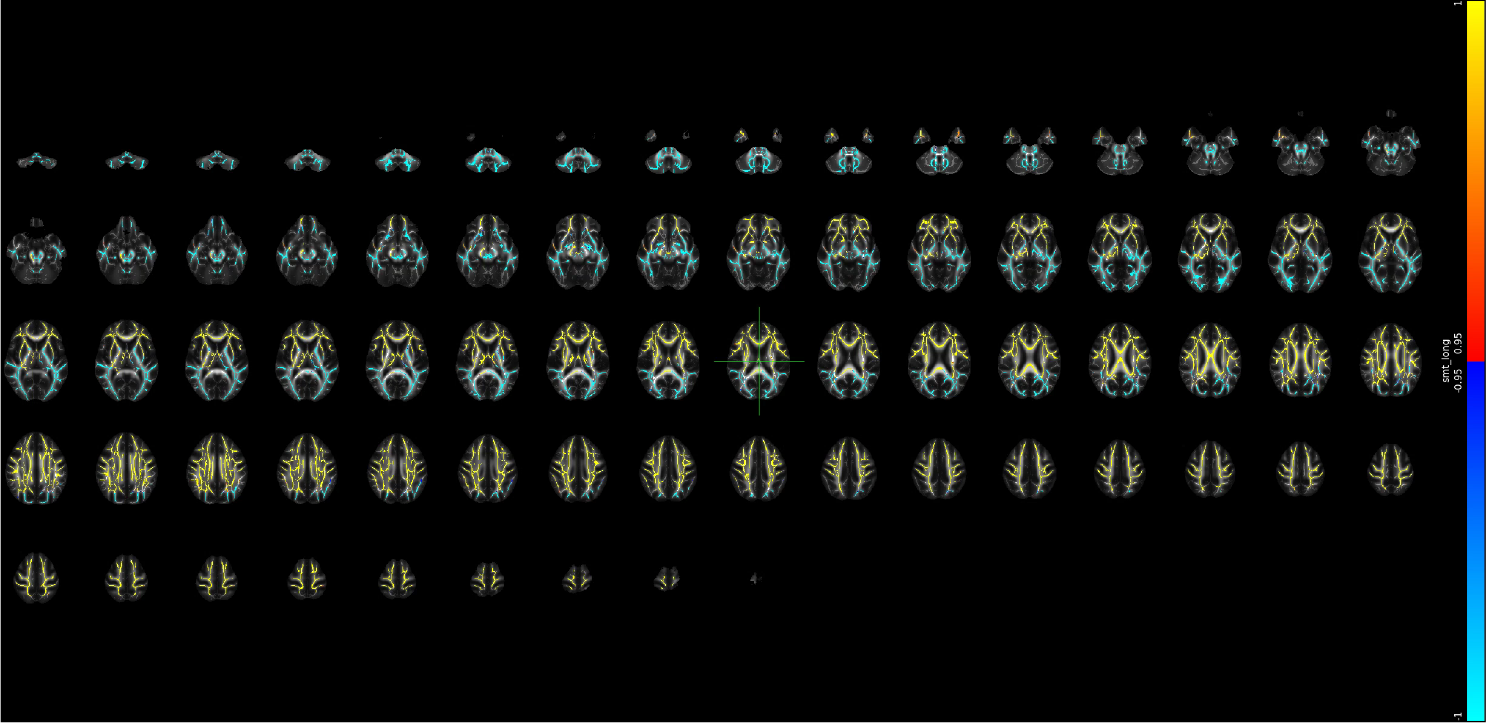

## Supplementary Note 52 Voxel-level changes in SMT - transverse diffusion coefficient

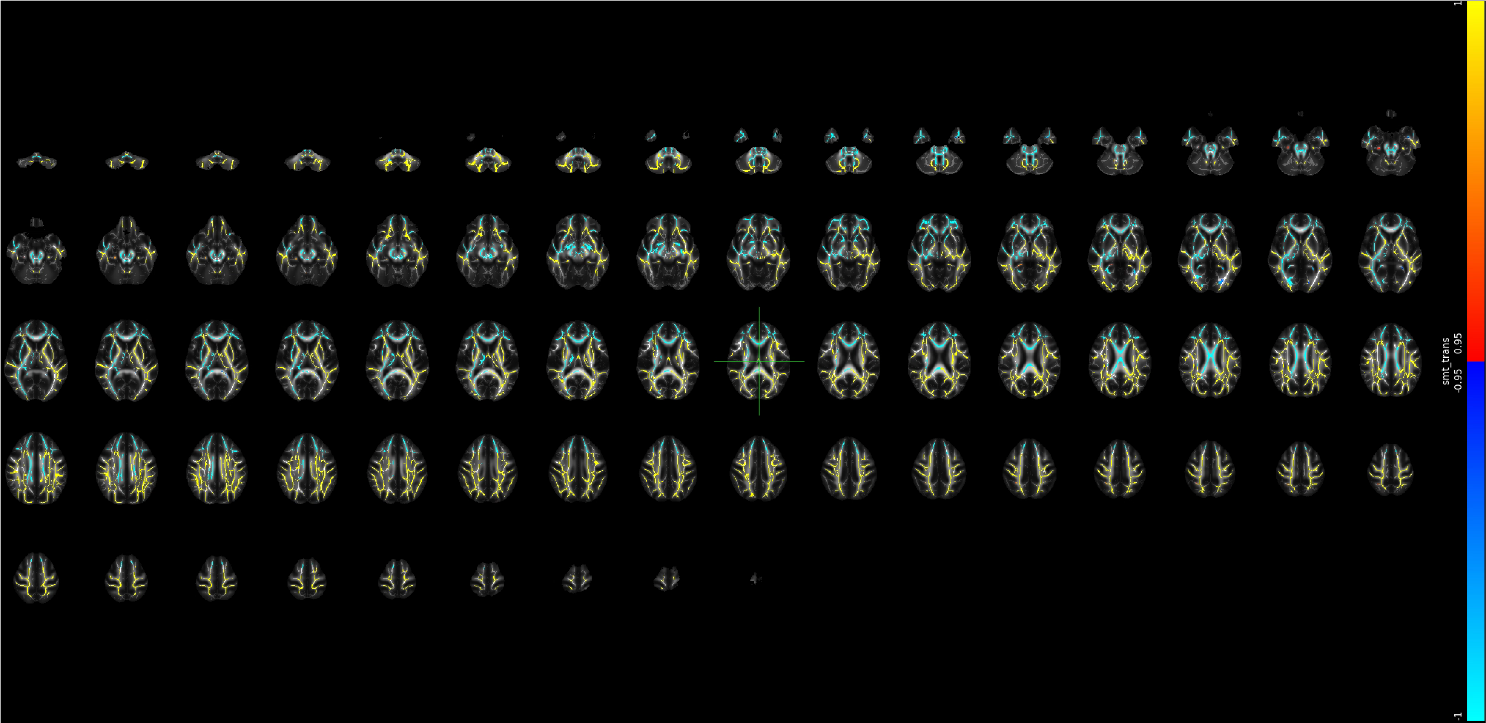

## Notes

### Author Declarations

This study has been conducted using UKB data under Application 27412.

